# Genetic and phenotypic architecture of human myocardial trabeculation

**DOI:** 10.1101/2024.03.26.24304726

**Authors:** Kathryn A McGurk, Mengyun Qiao, Sean L Zheng, Arunashis Sau, Albert Henry, Antonio Luiz P Ribeiro, Antônio H Ribeiro, Fu Siong Ng, R Thomas Lumbers, Wenjia Bai, James S Ware, Declan P O’Regan

## Abstract

Cardiac trabeculae form a network of muscular strands that line the inner surfaces of the heart. Their development depends on multiscale morphogenetic processes and, while highly conserved across vertebrate evolution, their role in the pathophysiology of the mature heart is not fully understood. We report variant associations across the allele frequency spectrum for trabecular morphology in 47,803 participants of the UK Biobank using fractal dimension analysis of cardiac imaging. We identified an association between trabeculation and rare variants in 56 genes that regulate myocardial contractility and ventricular development. Genome-wide association studies identified 68 novel loci in pathways that regulate sarcomeric function, differentiation of the conduction system, and cell fate determination. We found that trabeculation-associated variants were modifiers of cardiomyopathy phenotypes with opposing effects in hypertrophic and dilated cardiomyopathy. Together, these data provide insights into mechanisms that regulate trabecular development and plasticity, and identify a potential role in modifying monogenic disease expression.

## Introduction

The endocardial surfaces of the adult human heart are lined by a fenestrated network of muscular trabeculae that form an interface between the compact myocardium and intra-cardiac blood flow. Trabeculae enable nutrient and oxygen diffusion from the blood to the myocardium in early development^1^. The ventricular trabecular myocardium develops as a sponge-like network of cardiomyocytes that is critical for the development of the conduction system and ventricular chamber maturation^2,3^. Factors controlling their coordinated development are still emerging but depend on gene expression regulating cardiomyocyte polarity, cell adhesion, and actin cytoskeleton dynamics, where tension heterogeneity directs the patterning of the myocardial wall during organogenesis^2,4^. The function of trabeculae in the adult heart is less well understood, but they are thought to play a role in achieving efficient cardiac performance through force transmission and modifying flow dynamics^5^.

Increased trabeculation develops in healthy individuals as a physiological and reversible phenotypic adaptation to altered loading conditions, for instance in pregnancy and athletic training^6,7^. Hypertrabeculation is also observed in the context of heart failure, cardiomyopathy, and other circulatory and muscular disorders^3,8–10^. Trabecular remodelling may share mechanisms with the underlying myocardial disease^10^. While trabeculae are thought to play a pathophysiological role in the natural history of ventricular remodeling^11,12^, it is plausible that they are regulated by genetic modifiers of cardiomyopathy^3^. Exploration of the pleiotropy between the presence of hypertrabeculation and cardiomyopathies, identification of modifiers of the natural history of trabeculation, and discovery of the full spectrum of genome-wide common and rare genetic factors with influence over the variation in trabecular morphology, would aid our understanding of this complex and highly conserved phenotype.

Here, we use a deep learning approach for image segmentation and apply fractal dimension analysis as a quantitative measure of trabecular complexity^11,13–16^. We analysed imaging in 47,803 participants of UK Biobank with genotyping and whole exome sequencing to discover common and rare genetic variants related to adult cardiac trabeculation in the left ventricle (**Figure 1**) and test potential causal relationships using genetic instruments.

**Figure 1.**
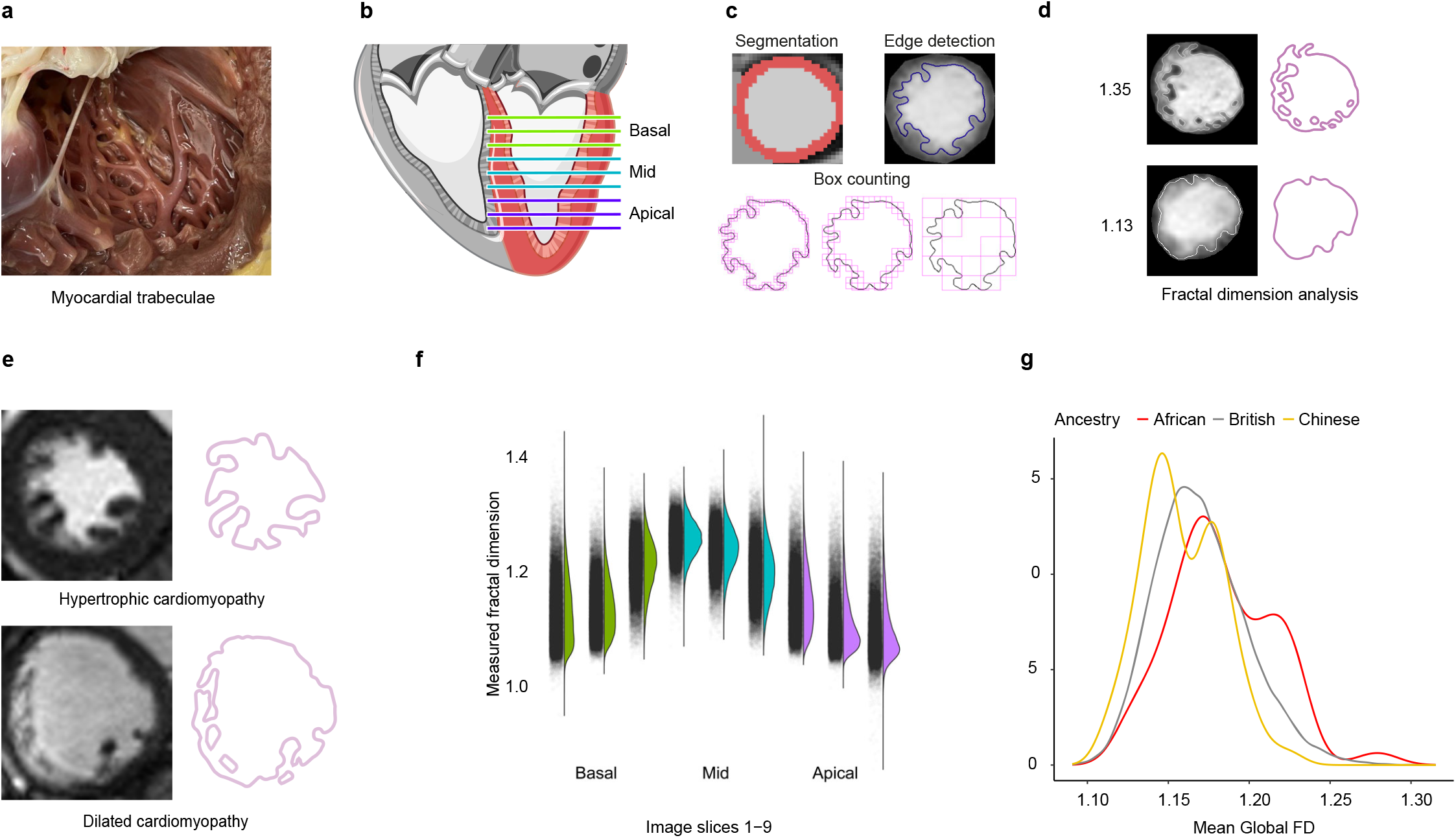
Summary of the analysis of trabeculation. **a**, Myocardial trabeculae on the endocardial surface of the human left ventricle. Image credit: Lukas Mach/Imperial College London. **b**, Cardiac magnetic resonance imaging of the left ventricle was acquired in the short-axis plane from base to apex. Image credit: Arpatsara/Shutterstock.com **c, d**, The myocardium was segmented using deep learning algorithms and edge detection used to define the boundary between the trabeculae and the blood pool. Trabecular complexity was defined by measuring the fractal dimension (FD) of this boundary using a box-counting methodology. **e**, Examples of trabecular morphology and edge detection are given for participants with hypertrophic and dilated cardiomyopathy cardiomyopathies. The images have been reproduced with permission from the UK Biobank. **f**, The distribution of FD at each level of the left ventricle. **g**, The distributions of FD by ancestry. African ancestry had increased mean global FD and Chinese ancestry had decreased mean global FD.

## Results

### Study overview

The UK Biobank study recruited 500,000 participants aged 40 to 69 years in the United Kingdom between 2006 and 2010^17^. Genotyping array data and exome sequencing data are available for over 450,000 participants. A sub-study recalled participants for cardiac magnetic resonance imaging (CMR)^18^ and volumetric traits were measured using quality-controlled deep learning algorithms^19–21^. Imaging was made available in two releases which formed a discovery group of 38,245 and a validation group of 9,558 participants.

Trabecular morphology was quantified using edge detection of the endocardium, to derive a scale-invariant fractal dimension ratio for each slice, where a higher value indicates a greater degree of surface complexity (**Figure 1**)^12,14^. The traits were adjusted using multiple linear regression for age at scan, age^2^, sex (not significant), age:sex interaction (not significant), imaging centre, body surface area, systolic blood pressure, vigorous exercise, and 10 genetic principal components of ancestry (significant associations with PCs 1, 2, 3, and 8; **Figure S1, Table S1**). Comparisons were also undertaken with and without adjustment for left ventricular end-diastolic volume (LVEDV). Additional outcome and phenotypic data included hospital episode statistics, self-reported questionnaire data, and activity data.

Trabecular traits were assessed for association with genetic, phenotypic, and clinical outcome data (**Figure 2**). We analyzed the cohort as separate discovery and validation cohorts as well as in combination, for rare variant burden analyses, for genome-wide association studies, and for association with curated cardiomyopathy-associated variants. Polygenic risk scores derived from published case/control cardiomyopathy GWAS and genetic correlation analyses were analyzed. Two-sample Mendelian randomization assessed for causality of trabeculation, cardiomyopathy, and heart failure. Curated and phenome-wide association study clinical outcomes were analyzed. Ancestry, alcohol, measures of physical activity, electrocardiogram diagnoses, and relationships with CMR-derived measures were assessed.

**Figure 2.**
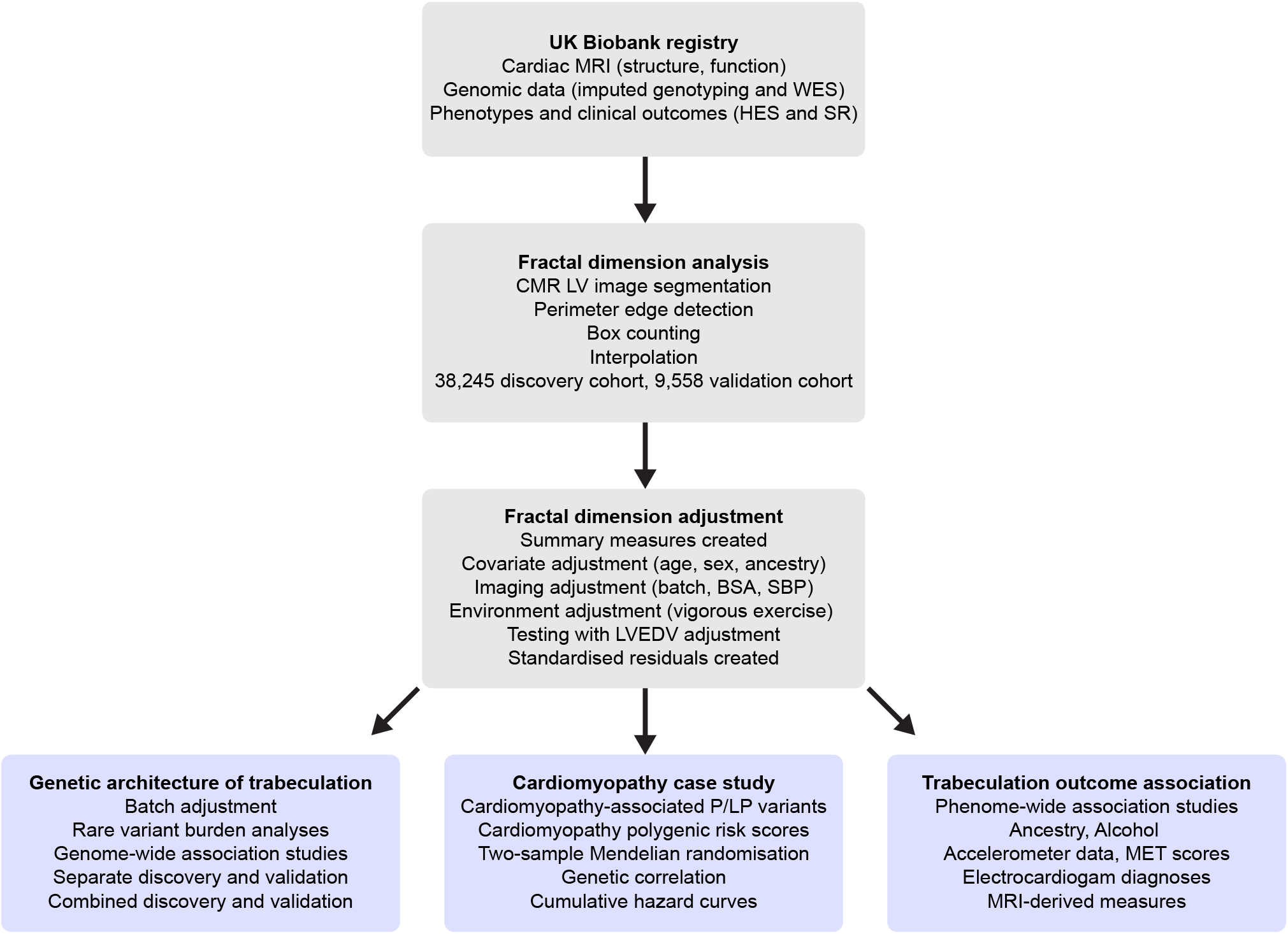
Study flowchart. A summary of the main steps in our analysis of fractal dimension and the genetic and outcome associations. MRI, magnetic resonance imaging; CMR, cardiac MRI; WES, exome sequencing; HES, hospital episode statistics; SR, self reported; LV, left ventricle; BSA, body surface area; SBP, systolic blood pressure; LVEDV, left ventricular end-diastolic volume; P/LP, pathogenic/likely pathogenic; MET, metabolic equivalent of activity.

### Modifiers of trabecular morphology

African ancestry had increased mean global fractal dimension compared to White British ancestry that dominates the UK Biobank demographics (*β*=0.28, SE=0.07, *P*=0.0003751; **Figure S28**). Indian, Chinese, and Bangladeshi ancestry had the lowest mean global fractal dimension (*β*=0.19, SE=0.08, *P*=0.008049 compared to White British ancestry). The association with African ancestry was independent of LVEDV, LV mass, and body mass index.

There was no significant difference in mean trabeculation (regardless of LVEDV) with obstetric history, although the hypertrabeculation observed during pregnancy is expected to be reversible^7^. Moderate or high alcohol intake (comparing the top 10% to the bottom 10% of alcohol intake (g/day^22^)) was associated with a high fractal dimension (*β*=0.09, SE=0.01, n=2,847 each, *P*=0.0006396; **Figure S2**), independent of LVEDV (P=0.0169). Physical activity increased fractal dimension in part due to LVEDV (**Figure S2; Supplemental methods**). Mean global fractal dimension had the strongest relationship with CMR measures of left ventricular volume (end-diastolic (LVEDV R=0.35), systolic (LVESV R=0.32), stroke volume (LVSV R=0.30)) and strain (global peak radial strain (Err R=**-**0.19), global peak systolic circumferential strain (Ecc R=0.28); **Figures S28-S29**). Increased mean global fractal dimension was associated with decreased ejection fraction (R=**-**0.13), an inverse relationship that has been described for trabecular morphology previously^23^.

### Phenome-wide association studies

Phenome-wide association studies (PheWASs) identified significant associations between measures of trabecular morphology and cardiovascular disease-related clinical outcomes (**Figure 3, Figure S18, Figure S32, Table S8**). These included cardiomyopathies, heart failure, conduction disorders, and valve diseases. Of note, no significant association was observed with thromboembolic events or stroke. To assess for pleiotropy with current cardiomyopathy diagnostic imaging measures, we analysed PheWAS of maximum wall thickness (max WT), systolic blood pressure (SBP), end-diastolic volume (LVEDV), and ejection fraction (LVEF) alongside the PheWASs of trabecular morphology (**Figure 3**). Conduction disorders, cardiomyopathies, heart failure, valve diseases, and myocardial infarction were identified by all traits. Hypertension was associated with max WT, fractal dimension, and SBP. Associations with fractal dimension were identified for UK Biobank baseline electrocardiogram diagnoses^24^ of first-degree AV block, left bundle branch block, sinus bradycardia, and atrial fibrillation, of which left bundle branch block, sinus bradycardia, and atrial fibrillation were independent of LVEDV.

**Figure 3.**
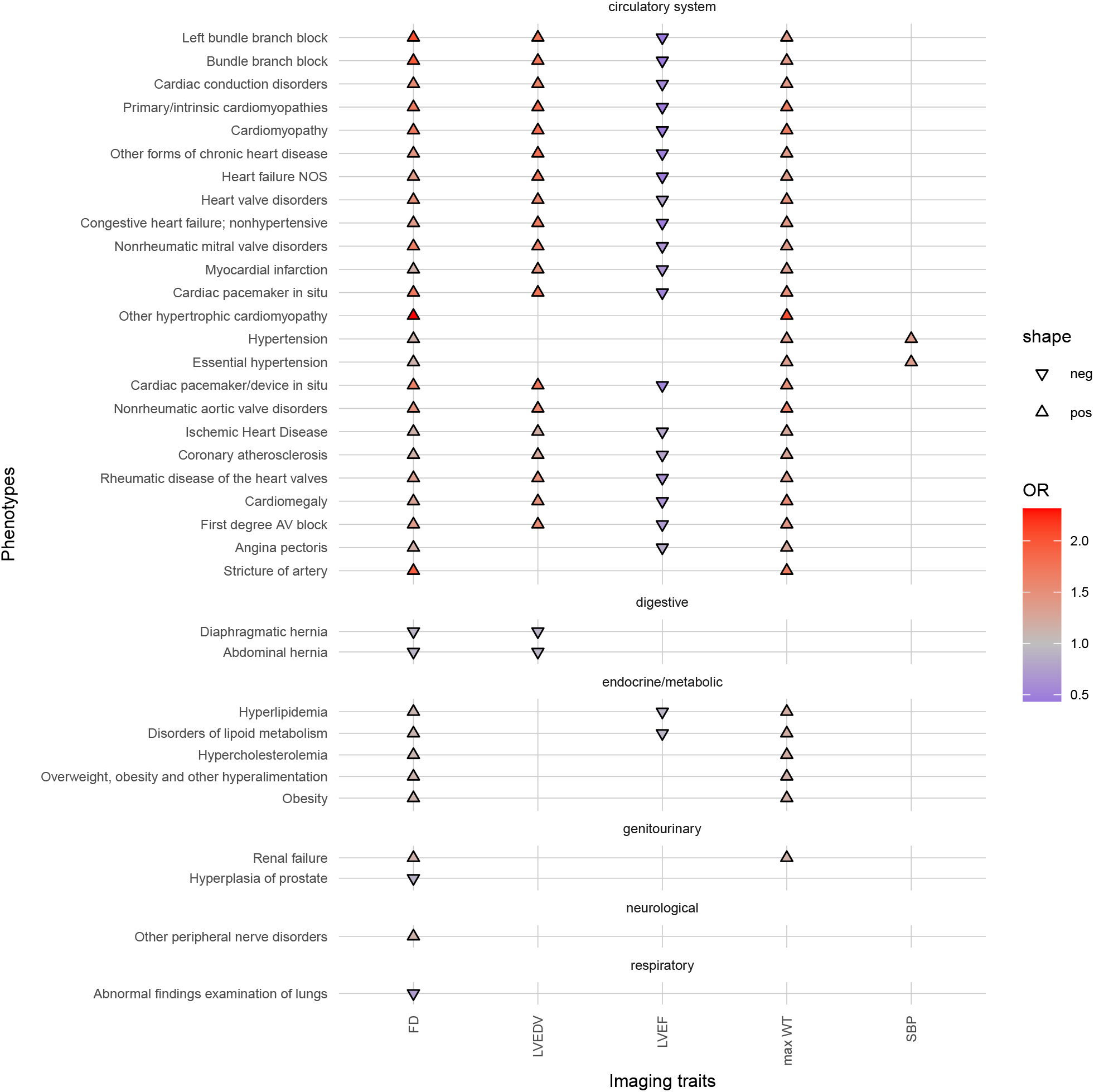
Phenome-wide association study of trabecular complexity and other CMR-derived traits. Maximum wall thickness (max WT) for HCM, end-diastolic volume (LVEDV) for DCM, systolic blood pressure (SBP), and ejection fraction (LVEF), were analyzed to assess pleiotropy between the trabecular complexity (FD) and remodeling traits altered in cardiomyopathies. Fractal dimension was measured at different spatial locations in the left ventricle and the aggregate of all results is shown. The analyses were completed on 38,245 participants of the UK Biobank population. Phenotypes as phecodes are described on the y-axis with the phecode category separating the groups and the imaging traits are on the x-axis. Each point denotes a significant PheWAS association with a Bonferroni correction for 1,163 analyzed phecodes. The shape and colour denote the direction of effect and odds ratio. Categories and phenotypes other than the circulatory system category that did not associate with measures of trabecular morphology but were associated with the imaging measures were removed from the plot for clarity, see the supplementary for the full PheWAS results.

### Rare variant association studies

We identified and validated 30 genes in which rare protein-altering variants were associated with trabecular morphology (**Figure 4, Table S7**). This included genes involved in inherited cardiac conditions, calcium signaling, and implicated in neurodevelopment. Analysis of the full dataset (discovery and validation cohorts together) identified 26 further genes (**Figure 4, Table S10**) including genes known to regulate trabeculation and associated with cardiomyopathy.

**Figure 4.**
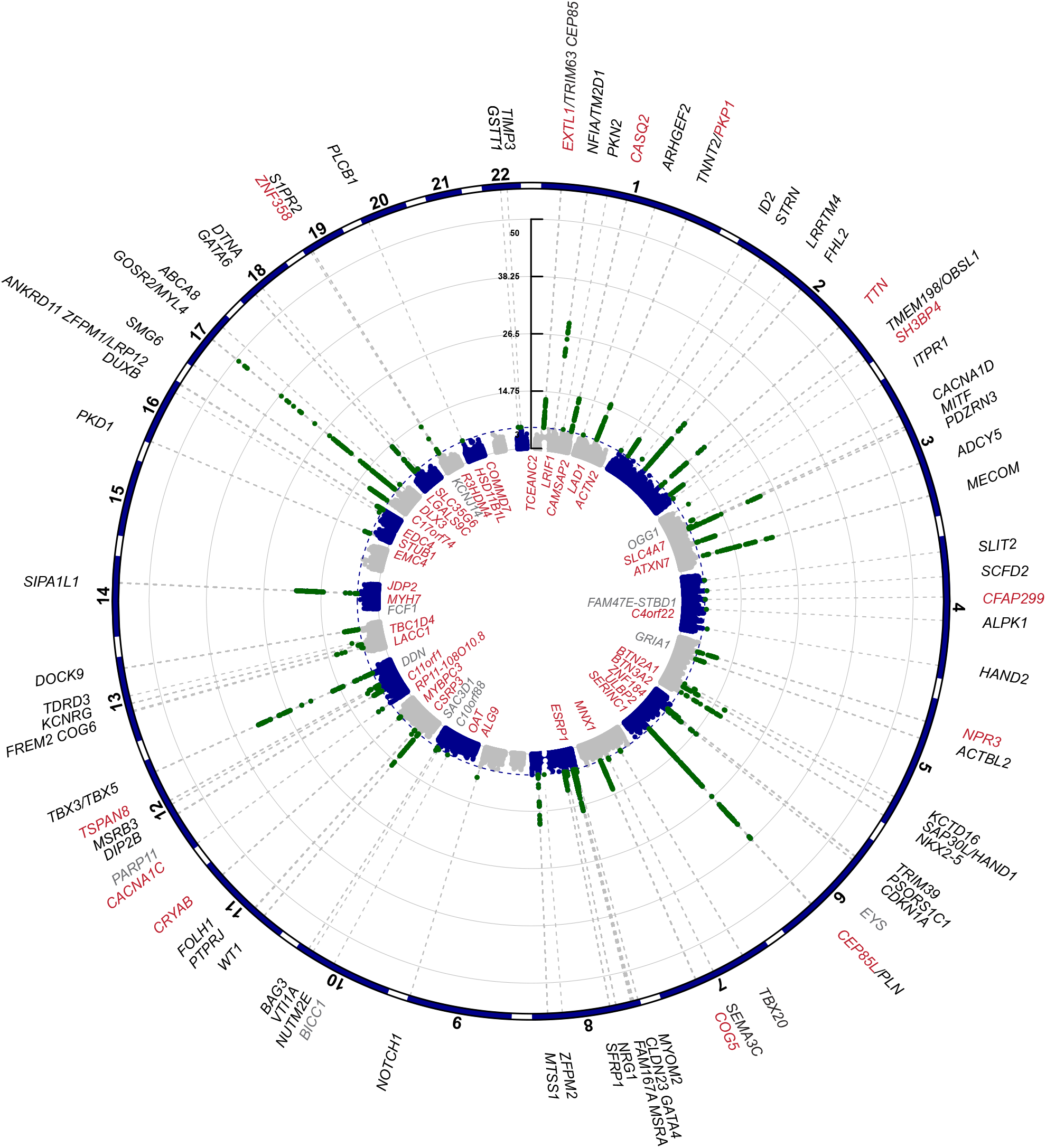
Manhattan plot for genetic loci associated with trabeculation. Common and rare variant genetic analyses for trabecular complexity assessed with fractal dimension analysis. The prioritized gene is noted for the significant loci identified through combined discovery and validation of common variant GWAS on the outside of a meta-Manhattan plot of the minimum *P* value for each SNP across measures of trabeculation. The rare variant association study (RVAS) identified genes with a burden of protein-altering variants that are associated with trabecular morphology (depicted inside the circle). Genes in red on the outside circle denote a gene identified through both GWAS and RVAS. Genes in grey were only identified in the discovery analysis and not in the full analysis of combined discovery and validation cohorts.

Trabeculation was influenced by genes with roles in cardiac function and disease (*TTN, MYBPC3, MYH7, CRYAB, CACNA1C, CASQ2, PLN*). *TTN* encodes a large, abundant protein of striated muscle, which has a key role in dilated cardiomyopathy. Loss-of-function variants increased trabecular morphology with the strongest effects towards the apex. *MYBPC3* (Myosin Binding Protein C3) encodes the cardiac isoform of myosin-binding protein C. Protein-altering variants in *MYBPC3* are the most common genetic cause of hypertrophic cardiomyopathy^25^. Missense variants in *MYH7* (cardiac myosin heavy chain beta) have definitive evidence of causation of HCM or DCM (via distinct mechanisms) and increased trabeculation. *CRYAB* (Crystallin Alpha B) is an aggregating chaperone that is mainly expressed in the heart and has definitive evidence for causing Alpha-B crystallinopathy^25^, where left ventricular hypertrophy is observed with overt syndromic features. Missense variants increased apical trabeculation. *CACNA1C* (Calcium Voltage-Gated Channel Subunit Alpha 1C) is involved in cellular calcium influx after depolarisation. The gene has definitive evidence for Timothy syndrome which is a multiorgan disorder that includes cardiovascular malformation and fatal arrhythmias. Missense variants decreased trabeculation. *CASQ2* (Calsequestrin 2) is a calcium-binding protein that stores calcium for muscle function. It is highly expressed in the heart and arteries and has definitive evidence for causing catecholaminergic polymorphic ventricular tachycardia. Missense variants in the gene associated with decreased apical trabeculation. *CEP85L* (Centrosomal Protein 85 Like) has broad tissue expression and has been linked to cancer and lissencephaly. The single exon gene *PLN* (Phospholamban) sits within the *CEP85L* locus. *PLN* has the highest expression in the heart and arteries and has definitive evidence for hypertrophic cardiomyopathy. Missense variants in the *CEP85L* region decreased trabeculation.

### Genome-wide association studies

The SNP-based heritability was estimated for each of the trabecular morphology measures (**Table S3**). Mean global fractal dimension had the highest estimated heritability (h^2^=0.43). The most basal and apical slices had heritability estimates of ∼0.2 while slices in the mid-region of the left ventricle (slices 5-7) had the highest estimated heritability of the slices (h^2^∼0.4).

We identified 32 loci from GWASs of 30,419 participants with validation in 7,593 participants and replicated previous studies^12^ (**Figure 4, Figures S4-S16, Table S4-S5**). We used LocusZoom, eQTLs, and transcriptome-wide association study to prioritise genes at each locus. The identified associations include loci of definitive evidence cardiomyopathy-associated genes (*TNNT2, TTN, PLN*) and colocalise with recent HCM (e.g., *TBX3, STRN, MTSS1*^26^), DCM (e.g., *PKD1, STRN, MITF, CDKN1A*^27^), and heart failure GWAS^28^. For sensitivity analyses, we removed any individuals in the UK Biobank who had a diagnosis of any cardiomyopathy or heart failure. All variants replicated except for the loci at *SFRP1, NUTM2E, SCFD2*, and *EYS*. All validated loci remained with adjustment for LVEDV, only loci at *MITF, PSORS1C1, EYS*, and *BICC1* were less significant (*P*=>5x10^−8^; **Figure S3**).

Analysis of the full GWAS (38,012 European participants; both discovery and validation cohorts) identified 36 further significant loci of (**Table S9**). This includes genes that regulate the development of trabeculation (*NKX2-5, TBX20, HAND2, NRG1, NOTCH1, DTNA*) and those with evidence of involvement in cardiomyopathies (*BAG3, CRYAB, RIT1 (ARHGEF2*)). We validated 13 loci through rare variant association studies (RVAS; *EXTL1, CASQ2, CEP85L, COG5, TSPAN8, TTN, SH3BP4, CFAP299, CRYAB, CACNA1C, PKP1 (TNNT2), NPR3, ZNF358*). Significant gene ontology (GO) resource enrichment analysis showed the strongest relationship with cardiac myofibril and sarcomeric cellular components, electrical conduction His cell differentiation, ventricle formation, and muscle cell fate commitment (**Table S13**).

We found an overlap with loci identified here and those identified from GWAS of QRS duration^29^; calcium handling genes (*CASQ2, PLN, CACNA1D*), transcription factors (*TBX3, HAND1, NFIA*), cyclin-dependent kinase inhibitors (*CDKN1A*), and others (*SIPA1L1, GOSR2, STRN*), but not cardiac sodium channel genes (*SCN5A* and *SCN10A*). The identification of *TTN, NKX2-5, MTSS1, TNNT2*, and *CACNA1C*, suggest a relationship with trabeculation in ventricular depolarisation and repolarisation^30^. Many genes identified here have been found to influence ECG traits (*TRIM63, DES, NPR3, NKX2-5, CDKN1A, SFRP1, WT1, ZFPM1, GATA6, NFIA, TNNT2, TTN, PDZRN3, HAND1, MTSS1, DEFB136, TBX3, GOSR2/MYH6, ZNF358, FLRT2, HAND2*)^30^. *CACNA1D* has moderate evidence for causing autosomal recessive sinoatrial node dysfunction and deafness.

The association with *NKX2-5, NOTCH1, GATA4, GATA6*, and *HAND1*, links trabeculation to congenital heart disease (CHD). *NKX2-5, NOTCH1*, and *GATA4*, have high confidence for non-syndromic CHD, atrial septal defect (*NKX2-5, GATA4*), aortic valve stenosis and tetralogy of fallot (*NOTCH1*). *NKX2-5* when suppressed causes excessive trabeculation in models^10^ and it is identified in carriers with LVNC alongside DCM or conduction disease^31^. GATA6 is a transcription factor with a role during heart development and has been linked to several congenital heart disease phenotypes previously^32^ and *HAND1* has moderate evidence for congenital heart disease^33^. *PKD1* is associated with polycystic kidney disease, a syndromic congenital heart disease^33^.

### Genetic modifiers of trabeculation in cardiomyopathy

Hypertrabeculation occurs in patients with HCM and DCM^12,20,34^, although the morphology of trabeculae in these cardiomyopathies may reflect the underlying disease and pattern of remodeling (**Figure 1**). In the UK Biobank, fractal dimension was increased in participants diagnosed with hypertrophic and dilated cardiomyopathies, atrial fibrillation and flutter, and congenital heart disease (**Supplemental methods**). The relationship between trabeculation and HCM was independent of LVEDV and wall thickness but was related to the degree of ventricular dilatation in participants with DCM (**Figures S28-S29; Supplemental methods**).

About 30% of cardiomyopathy cases are attributable to a rare variant in a CM-associated gene, many of which encode components of the sarcomere^35^. After excluding individuals with a diagnosis of cardiomyopathy, UK biobank participants carrying either a HCM or DCM-associated pathogenic or likely pathogenic variant had increased fractal dimension (compared with participants without such a variant; **Figure S17, Figure S31**). Participants with a diagnosis of cardiomyopathy also had increased fractal dimension, irrespective of the presence/absence of a potentially causative sarcomere variant. The presence of hypertrabeculation on imaging was a risk factor for a subsequent diagnosis of heart failure (HR=1.3, 95% CI=1.08-1.6), mitral valve disorders (HR=1.4, 95% CI=1.12-1.70), and bundle branch block (HR=1.2, 95% CI=1.01-1.50; **Figures S30-S31**). Of 81 people with cardiomyopathy reported after imaging, 25% (n=20) had hypertrabeculation on imaging and none had hypotrabeculation. A less trabeculated ventricle was a risk factor for heart failure when adjusted for LVEDV (**Figure S31**).

Polygenic risk scores (PRS) for HCM derived from case-control common variant GWAS, showed an inverse relationship with fractal dimension in UK Biobank (comparing the top 10% of the PRS to the bottom 10%: *β*=-0.12, SE=0.01, n=3,039 each, *P*=3.577x10^−6^), while the opposite relationship was identified with the DCM PRS (top 10% of the PRS to the bottom 10%: *β*=0.11, SE=0.01, n=3,042 each, *P*=3.789x10^−5^). The strongest genetic covariance (C(G)=0.15-0.16) and correlation (rG=0.40-0.43) was between trabecular morphology and LVEDV (**Table S2**). A negative correlation was observed with maximum wall thickness (rG=**-**0.10-**-**0.05; **Table S2**).

To assess causality we used two-sample Mendelian randomization (MR) for association testing of genetically-predicted levels of an exposure on an outcome^36^ (see **Supplemental methods; Figure 5, Figures S19-S27, Table S6**). We did not find that trabeculation (mean global FD) as an exposure had a causal association with cardiomyopathies or heart failure outcomes that was independent of LV remodelling (see **Table S6** for multivariable MR analyses). However, we identified opposing directions of effects for many trabeculation-associated variants when applied to HCM and DCM, suggesting they modify the phenotype differently in each disease (**Figure 5**). We identified evidence supporting causal effects of genetic liability to DCM with increased trabeculation and heart failure. Conversely, genetic liability to HCM was negatively associated with increased trabeculation, DCM and heart failure. These findings suggest that shared biological processes contribute to trabeculation and heart failure in both cardiomyopathies but through opposing mechanisms that are remodelling-dependent.

**Figure 5.**
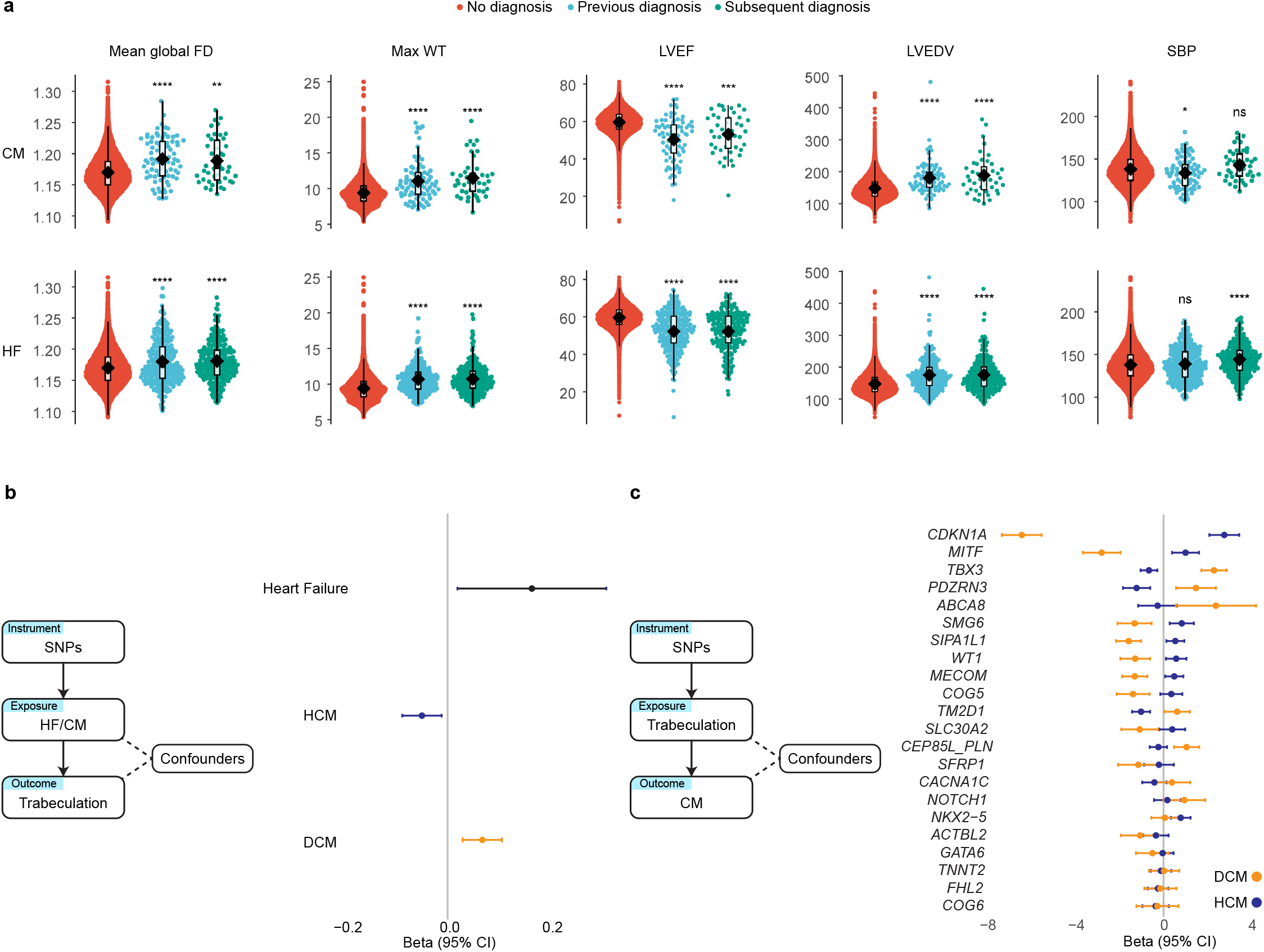
Relationship between trabecular complexity and cardiomyopathy. Increased FD may occur early in the natural history of cardiomyopathies (CM) and heart failure (HF). **a**, Imaging biomarkers (left ventricular end-diastolic volume (LVEDV), ejection fraction (LVEF), maximum wall thickness (maxWT)), systolic blood pressure (SBP), and mean global fractal dimension (FD)) for three groups of participants depending on cardiomyopathy (CM) or heart failure (HF) diagnosis status; as i) no diagnosis, ii) diagnosis recorded previous to imaging, or iii) diagnosis recorded after imaging. Analyses were performed on 38,245 participants of the UK Biobank. Student’s two-sided t-test was used to compare the group means to individuals with no diagnosis. The lower and upper hinges in the box plot correspond to the 25th and 75th percentiles (interquartile range (IQR)), respectively. The horizontal line in the box plot indicates the median. The lower and upper whiskers extend from the hinge to the smallest and largest values no further than 1.5× the IQR, respectively. Each dot is one individual. **b**, Mendelian randomization genetic determination model of CM and HF genetic instruments as exposures for trabeculation outcome. Common genetic loci influencing the variability of cardiac function in an additive fashion may be deferentially involved depending on the CM substrate. Common genetic variants associated with DCM increased trabeculation and HF risk. Common genetic variants associated with HCM had an inverse relationship with trabeculation. Two-sample Mendelian randomization was undertaken with genetically determined mean global FD (using summary statistics from the GWAS of 38,245 UK biobank participants) and genetically determined dilated cardiomyopathy (DCM), hypertrophic cardiomyopathy (HCM), and HF (from GWAS summary statistics of published data). Presented are the IVW estimates. **c**, Mendelian randomization genetic determination model of trabeculation genetic instruments as exposures for CM outcomes was not significant. The single SNP funnel plot for trabeculation genetic instruments as exposures for cardiomyopathies shows that trabeculation-associated variants have opposing directions of effect for DCM or HCM. The plot is ordered by the delta of the betas. Four variants were removed that were not present in the HCM summary statistics so GWAS betas could not be compared with DCM.

## Discussion

Myocardial trabeculae are highly conserved structures in vertebrates where their complex forms are driven by multiscale interactions during development^37^. Disrupting their development in animal models is frequently lethal, but their role in adults is not fully understood and may relate to maintaining efficient intraventricular flow patterns and force transmission in health and disease^5,12^. Here, we analyse myocardial trabecular traits in adult humans across the allele frequency spectrum discovering new loci regulating chamber maturation, complex biological patterning, and conduction traits. We found that the same trabeculation-associated common variants had opposing effects in cardiomyopathies suggesting that they cause distinct phenotypes in different diseases.

Several discovered loci are known to regulate trabecular development and cell fate determination. Common variants mapped to genes in *NRG1, NOTCH1*, and *NKX2-5*, which regulate trabecular morphogenesis and define the building plan for trabeculation through Neuregulin 1β/ErbB signaling and modifying cardiac jelly dynamics in the developing heart^2^. The zinc finger transcription factor *GATA6* is expressed early in the developing heart and is critical for activation of the cardiomyocyte gene expression program^38^. Of note, the *MIB1* gene is located upstream of *GATA6* and encodes an E3 ubiquitin ligase that promotes endocytosis of NOTCH ligands. Variants in this gene cause hypertrabeculation in an autosomal-dominant fashion and inactivation causes hypertrabeculation in mice^39^. The Hand proteins are also essential regulators of cardiac development and HAND2 is a crucial downstream transcriptional effector of endocardial Notch signaling during embryonic trabeculation^40^. The replicated common variant loci in *GOSR2* and *MTSS1* regulate cytoskeletal branching and may increase structural surface area in different organ systems^12,41,42^. Newly discovered loci in *CEP85L* and *PKD1* suggest there may also be a role for cilia and microtubule organisation in trabecular formation - potentially through endocardial mechanosensing and signaling^43,44^.

Trabeculation is a dynamic phenotype that responds to transient changes in loading conditions^6,7^, but the pathophysiological interactions between trabeculation and remodeling in cardiomyopathy are not known. We found that increased trabeculation can be evident at presentation in some patients with cardiomyopathy, suggesting that additional common or rare genetic modifiers may contribute to the phenotype^3^. We identified individually-rare protein-altering variants, including non-sarcomeric genes, that could be plausible candidates for such modifiers. We did not find that aggregating common variants, as an instrument for trabeculation, led to an association with outcomes when pruning cardiomyopathy-associated variants. However, this relationship may be better understood by the finding that many trabeculation-associated variants show opposing directions of effects depending on the underlying cardiomyopathy, suggesting they modify the phenotype differently in each disease. Variants in *CDKN1A*, a non-sarcomeric gene that regulates stress-induced remodelling^45^ and a regulator of cell-cycle progression, used as an instrument for trabeculation showed the strongest divergent effects on HCM and DCM as outcomes. There are genetic correlations between other LV traits and cardiomyopathies which also have opposing effects in HCM and DCM^46^. Together, these findings suggest that changes in trabeculation may occur early in the pathogenesis of heart disease, can be modified by genes regulating pathways outside the sarcomere, and causality depends on the underlying cardiomyopathic substrate.

Increased trabeculation is a characteristic of both HCM and DCM, but for some genes, the phenotypic associations may be predominantly associated with isolated hypertrabeculation phenotypes^3^. We identified several genes associated with trabeculation that also have evidence for what is termed as “left ventricular non-compaction cardiomyopathy” (LVNC) (*MYH7, MIB1 (GATA6), DES, MYBPC3, TTN, ACTN2, LMNA, PLN, TBX20, TBX5, DTNA, TNNT2*)^3,47,48^. The descriptive label for the phenotype is based on the proposed mechanism of arrested incorporation of trabeculae into the compacted wall during mammalian heart development^49^, although there is no direct evidence for such compaction in the developing human heart^50^. It has been previously reported that asymptomatic individuals with hypertrabeculation have a normal life expectancy, while those who are symptomatic with hypertrabeculation are at risk of adverse cardiac events and heart failure^9^. We found that both the lowest and highest extremes of trabeculation were associated with a greater risk of heart failure outcomes. A limitation of the current analysis is that it is at a single time point for each participant, so we do not know the temporal trajectory of trabecular adaptation and the relationship with remodelling.

In conclusion, genetic variants across the allele frequency spectrum are associated with trabecular traits in the mature human heart pointing to pathways that orchestrate early organ development, determine cell-fate commitment and regulate cytoskeletal complexity. Trabeculation-associated loci may also act as risk factors or disease modifiers in cardiomyopathy but have different effects depending on the genetic substrate. Myocardial trabeculae are an intricate and dynamic phenotype, and these findings reveal mechanisms of how these complex tissue forms emerge and their significance to adult health and disease.

## Data availability

All data arising from this analysis will be made available through the UK Biobank (https://biobank.ndph.ox.ac.uk/showcase/), GWAS catalog (https://www.ebi.ac.uk/gwas/), Github (https://github.com/ImperialCollegeLondon/trabecular_variants), and GWAS significant SNPs viewed on LocusZoom (https://my.locuszoom.org/). UK Biobank data is publicly available (https://www.ukbiobank.ac.uk/).

## CrediT statement

Conceptualization: K.A.M., D.P.O’R.; Methodology: K.A.M.; Formal Analysis: K.A.M.; Resources: D.P.O’R., J.S.W., F.S.N, T.R.L., W.B.; Data Curation: K.A.M., A.S., M.Q., S.L.Z, W.B., A.H.; Writing – Original Draft: K.A.M., D.P.O’R.; Writing – Review & Editing: (all authors); Visualization: K.A.M., D.P.O’R.; Funding Acquisition: K.A.M.

## Methods

### Study overview

The UK Biobank (UKB) study recruited 500,000 participants aged 40 to 69 years old from across the United Kingdom between 2006 and 2010 (National Research Ethics Service, 11/NW/0382)^17^. Genotyping array data and exome sequencing data are available for over 450,000 participants. This study was conducted under the terms of access of projects 40616 and 47602. All participants provided written informed consent. A sub-study recalled participants for cardiac magnetic resonance imaging (CMR)^18^, and volumetric traits measured using quality-controlled deep learning algorithms^19–21^. Imaging was made available in two releases which formed a discovery group of 38,245 and a validation group of 9,558 participants.

### Fractal analysis of trabecular morphology

Deep neural networks were used for short-axis cine end-diastolic left ventricular segmentation via a fully convolutional network to label pixels containing myocardium. Edge detection defined the trabecular boundary (representing the endocardial border, with the inclusion of trabeculation and papillary muscles) using the Sobel edge detection algorithm. Box counting across a range of sizes generated a log–log plot of box size and box count for which the negative gradient of a least-squares linear regression defined the fractal dimension (FD)^51^. Fractal analysis was automated using FracAnalyse software^11,13^ and adapted for the scale of the UK Biobank and derived segmentations (AutoFD^12^). The analysis was performed on each left ventricular slice.

A fractal dimension is a scale-invariant ratio providing an index of complexity. The raw fractal dimension data was interpolated using a Gaussian kernel local fit to a nine-slice template to allow for comparison across subjects. Summary statistics (minimum, mean, median, maximum) were calculated from the nine slices for four cardiac levels: global (1-9 slices), basal (1-3 slices), mid (4-6 slices), and apical (7-9 slices). The measures were adjusted for covariates: age at scan, age^2^, sex, age:sex interaction, imaging centre (batch effects), body surface area, systolic blood pressure, days per week of vigorous exercise, and 10 genetic principal components for ancestry, through multiple linear regression producing standardised residuals (mean=0, SD=1). See **Supplementary methods** for more information.

### Genetic association studies: rare, common, and cardiomyopathy-associated variants

Genotype calling was performed on the UK BiLEVE Axiom array and the UK Biobank Axiom array resulting in 805,426 markers in GRCh37 coordinates and quality control was undertaken^17^. The dataset was phased and 96M genotypes were imputed using the Haplotype Reference Consortium and UK10K haplotype resources^17^. Exome sequencing was undertaken on the Illumina NovaSeq 6000 platform. Reads were mapped to the hg38 reference genome and quality control was undertaken^52^.

Rare variant association studies (RVAS) were undertaken using Regenie software on the DNA Nexus Research Analysis Platform. The raw genotyping data for step 1 of Regenie was lifted from GrCh37 to GrCh38 using picard and bcftools. SNPs in autosomes with a minor allele frequency <0.01, missingness of >0.01, a minor allele count of <20, deviations from Hardy-Weinberg equilibrium (5x10^−15^), and individuals with greater than 10% missingness, were excluded from the analysis. Interchromosome, SNPs in linkage disequilibrium (indep-pairwise 1000 100 0.9), and areas of low complexity, were excluded for step 1. Exome sequencing data for step 2 was quality controlled for variants in the autosomes with missingness less than 10%, variants where less than 90% of all genotypes for that variant had a read depth less than 10, deviations from Hardy-Weinberg equilibrium (1x10^−15^), and individuals with more than 10% missingness. Step 2 of Regenie was run for the 25 trabeculation summary measures over different allele frequencies (singletons, 0.01, 0.001) for 6 overlapping, protein-altering variant, custom masks (LoF only; missense only (flagged by >1 of 5 deleterious software); missense only (all); missense only (flagged by all of 5 deleterious software); protein-altering variants (LoF and missense flagged by >1 of 5 deleterious software); protein-altering variants (all Lof and missense)), where the minimum minor allele count was at least 3. Bonferroni significance for 18,117 included genes was *P*<2.76x10^−6^ and the genes of the significant RVAS results in 29,480 participants were validated (*P*<0.05) in a holdout group of 7,293 participants. A full analysis was also completed by combining the discovery and validation cohorts.

For genome-wide association studies (GWAS), the imputed UK Biobank genotyping data was converted from bgen format to plink using plink2. Minor allele frequency of >0.001 in autosomes were included. Individuals with more than 5% missing genotypes and SNPs with more than 5% missingness were excluded. Participant sex discrepancies, heterozygosity, and relatedness were handled by keeping only European individuals and participants included in the UKBB principal components analysis^12,17^. SNPs deviating from Hardy-Weinberg equilibrium (1x10^−8^) and those with an imputation INFO score of <0.4 were excluded. Individuals with trabeculation data were extracted and GWAS was undertaken for 30,419 participants using GCTA software (version 64). A sparse genetic relationship matrix (GRM) was created and FastGWA was undertaken with a mixed linear model, adjusting for the genotyping array batch (and discovery or validation batch in the full analysis). 25 trabeculation summary traits were analysed and the significant GWAS results (*P*<5x10^−8^) were validated (*P*<0.05) in a holdout group of 7,593 participants. A full analysis was also completed by combining the discovery and validation cohorts (**Table S5, Table S9**). Genes of independent loci were prioritized through LocusZoom, eQTLs from GTEx v8, and transcriptome-wide association study (TWAS). TWAS with S-MultiXcan^53^ was completed using the GWAS summary statistics from GTEx V8 which identified 128 cardiac-specific (left ventricle and atrial appendage) genes and 336 genes from all tissues that significantly associated with trabeculation after correction for the number of tested genes (**Table S11-S12**)^26,27^. Phenotype associations through GWAS catalog, Phenoscanner, and PheWEB, were assessed (**Table S5, Table S9**), and resulting genes were analysed by gene ontology (GO) resource enrichment analysis using Panther (**Table S13**).

Cardiomyopathy-associated rare variants were identified similar to previously published^20,21^ for HCM and DCM. Individuals were classified as genotype negative (SARC-NEG) if they had no rare protein-altering genetic variation (minor allele frequency <0.001 in the UKB and the Genome Aggregation Database) in any genes that may cause or mimic HCM or DCM. These genes represented an inclusive list of genes with definitive or strong evidence of an association with CM, moderate evidence, and genes associated with syndromic phenotypes^25,54,55^. This SARC-NEG group was compared with individuals with disease-associated rare variants in genes with strong or definitive evidence for HCM (*MYBPC3, MYH7, MYL2, MYL3, TNNI3, TNNT2, TPM1, ACTC1*) and DCM (*BAG3, DES, DSP, FLNC, LMNA, MYH7, PLN, RBM20, SCN5A, TNNC1, TNNT2, TTN*). Analysis was restricted to robustly disease-associated variant classes for each gene and to variants sufficiently rare to cause penetrant disease (filtering allele frequency <0.00004 for HCM and 0.000084 for DCM^56^). Variants were classified as pathogenic/likely pathogenic (SARC-P/LP) if reported as P/LP for CM in ClinVar and confirmed by manual review. Further information on the variant curation pipeline can be found in the **Supplementary methods**.

### Statistical analysis

Phenome-wide association studies (PheWAS) were undertaken using the PheWAS R package with clinical outcomes and coded phenotypes converted to 1,840 categorical PheCodes. PheWAS was undertaken with the full cohort of 47,803 participants, all traits were adjusted using multiple linear regression for age at scan, age^2^, sex, age:sex interaction, imaging centre, body surface area, systolic blood pressure, vigorous exercise via questionnaire (days per week), and 10 genetic principal components of ancestry. P-values were deemed significant with Bonferroni adjustment for the number of PheCodes. Cumulative hazard curves were undertaken with the full cohort of 47,803 participants for mean global trabeculation and created using the first reported UK Biobank data (summarising the first date of report from all UK Biobank data (Hospital episode statistics, primary care, self-reported, etc.)) and the survival and survminer R packages from date of imaging to death or diagnosis date. Participants diagnosed with cardiomyopathy or heart failure before imaging were excluded. Hyper- and hypo-trabeculation were deemed > or < 1.5 standard deviations from the mean. Mendelian randomization was assessed using GWAS Summary statistics from published literature^26–28^ with mean global FD using the R packages TwoSampleMR and MVMR. Exposure variants were included if GWAS significant (*P*<5x10^−8^). The GWAS significant loci of mean global fractal dimension in the full cohort (n=38,012 European participants) were used to test causality for trabecular morphology in DCM, HCM, and HF, with and without adjustment for LVEDV. Tests of pleiotropy, steiger directionality, and heterogeneity were assessed (**Figures S19-S27, Table S6**). Heritability was estimated by creating a genetic relationship matrix in GCTA and using a restricted maximum likelihood analysis (REML) to estimate the variance explained by the SNPs that were used to estimate the GRM (**Table S3**). Genetic correlations were assessed using –reml-bivar in GCTA for two traits (**Table S2**). Polygenic risk scores for HCM and DCM were created previously from GWAS summary statistics and applied to the UK Biobank cohort^26,28,57^. Categorical variables were assessed using the Chi-Squared Test or Fisher’s Exact Test. Continuous variables were assessed using Student’s t-test. Please see **Supplementary methods** for more information.

