## Supplementary Material for "Genetic and phenotypic architecture of human myocardial trabeculation"

### Supplementary materials

#### Supplementary methods

##### Fractal analysis extra details

After analysis of AutoFD and FractAnalyse on the segmented UK Biobank cardiac MRI images[1,2], the data was interpolated. fracDecimateFD.R was used to interpolate the data to 9 slices (interpNoSlices=9) with a minimal slice cut-off of 6 (cut.off=6). summaryFD.R was used to create summary statistics from the interpolated results (sections="BMA", discard=FALSE) for basal, mid, and apical slices. The min, max, and medians were calculated using rowMins, rowMaxs, and rowMedians in R.

##### Adjustments and associations with covariates, ancestry, and curated clinical outcomes

The fractal dimensions for slices and summary measures were adjusted for Age at scan + (Age at scan)$^2$ + imaging centre (54-2.0) + reported sex + body surface area + automatic systolic blood pressure (4080-0.0) + Age:Sex (interaction term) + days/week of vigorous activity + 10 genetic principal components of ancestry (22009-0-(1-10)) (**Table S1**, **Figure S1**). Self-reported ancestry (21000-0-0) was used to assess any difference in trabecular morphology (TM) by ancestry. MET data was summarised in three variables: 22040.0.0 Summed MET minutes per week for all activity (**Figure S2**), 22035.0.0 Above moderate/vigorous recommendation, and 22039.0.0 MET minutes per week for vigorous activity. The curated clinical outcomes have been published previously[3,4]. The 2021 data release of refreshed Hospital Episode Statistics (HES) was used to identify the ICD codes and patients of the conditions. The UK Biobank provided the first occurrence of health outcomes defined by a 3-character ICD10 code using Hospital in-patient records, Death records, and Primary care records, which were used for cumulative hazard curves.

##### ECG diagnoses

UK Biobank ECGs from the first imaging visit (instance 2, n = 42,386) were labeled using a previously trained convolutional neural network[5] designed to identify six diagnoses from the ECG: sinus bradycardia, sinus tachycardia, left bundle branch block, right bundle branch block, 1st degree AV block, and atrial fibrillation. The ECGs were preprocessed with a bandpass filter 0.5 to 100hz, a notch filter at 60hz, and re-sampling to 400hz. Zero padding resulted in a signal with 4,096 samples for each lead for a 10s recording, which was used as input to the neural network model. The binary outputs (presence or absence of each diagnosis) were used for subsequent analyses.

##### HCM Variant curation

Variants 100 bp +/- the region of 29 HCM or LVH-associated genes were extracted from the whole exome sequencing data of 454,756 participants. MANE, protein-altering variants variants that had a MAF <0.1% in gnomAD and UK Biobank were identified. The UK Biobank exome data was annotated using Ensembl Variant Effect Predictor (VEP; version 105) with a plugin for gnomAD (version r2.1), LOFTEE, and SpliceAI. Intron variants that were pathogenic in ClinVar were manually curated for functional evidence of splicing. Splice region variants and other splice variants (excluding essential splice and splice donor 5th base), were included if they were predicted to cause splicing by SpliceAI (threshold >0.8). Step 1: 105,780 carriers of the 11,841 variants remaining include a list of individuals that are used to identify genotype-negative individuals in the UK Biobank. This includes 90,167 heterozygotes, 14,574 compound heterozygotes, and 1,390 homozygotes and 9 compound homozygotes (359 and 1 of which are also heterozygotes for other variants, respectively). There were 348,976 genotype negatives.

The variant list was then shortened to only include the 8 definitive evidence HCM genes (*MYBPC3, MYH7, TNNT2, TNNI3, TPM1, MYL2, MYL3, ACTC1*). LOFTEE was used to identify low-confidence pLoF variants and identify protein-altering variants (PAVs) that are mislabelled (e.g. NAGNAG sites). The variants were then filtered for disease-causing mechanisms; all MYBPC3 PAVs were kept, for the other 7 genes, only variants influencing gene product structure (indels, missense, start and stop lost) or gene product level but NMD escaping, were kept. Step 2: Carriers of the 2,708 variants remaining include a list of individuals that carry an HCM indeterminant variant. After exclusion for DCM variants (see Step3), 27,742 carriers were identified for the 2,690 variants, of which 26,734 were heterozygotes, 988 were compound heterozygotes, and 23 were homozygotes (3 of which were also heterozygotes for other variants).

The variant list was then shortened to only include variants that met a filtering allele frequencing (faf) of <0.00004 in gnomAD popmax faf and as a MAF in UK Biobank. Step 2b: Carriers of the 2,423 variants remaining include a list of individuals that carry an HCM indeterminant variant that is rare enough to have a substantial effect on the disease. After exclusion for DCM variants (see Step 3), 10,209 carriers were identified for the 2,405 variants, of which 10,031 were heterozygotes, 173 were compound heterozygotes, and 5 were homozygotes.

The variant list was then shortened to only include variants that would be called pathogenic/likely pathogenic if identified in a patient with HCM; using ClinVar, the variants were manually curated if they had any evidence of pathogenicity. 298 variants were assessed, of which 168 had strong evidence of pathogenicity in HCM, and 18 were identified with evidence of DCM. These 18 were excluded from Step 2 onwards. Carriers of the 168 variants remaining include a list of individuals that carry an HCM pathogenic/likely pathogenic variant. 929 carriers were identified for the 168 variants, of which 928 were heterozygotes, 1 was a compound heterozygote, and there were no homozygotes.

#### DCM Variant curation

Extraction was similar to the HCM variant curation pipeline but for 19 DCM-associated genes with definitive, strong, or moderate evidence for disease. Only TTNtvs in PSI>90% exons and variants with a splice prediction or curation of pathogenic (with assessment) were included. Step 1: 105,507 carriers of the 12,201 variants remaining include a list of individuals that are used to identify genotype-negative individuals in the UK Biobank. This includes 90,481 heterozygotes, 14,904 compound heterozygotes, 119 homozygotes and 3 compound homozygotes (46 and 1 of which are also heterozygous for other variants, respectively). There are 349,249 genotype negatives.

The variant list was then shortened to only include the 12 definitive or strong evidence DCM genes (*BAG3, DES, DSP, FLNC, LMNA, MYH7, PLN, RBM20, SCN5A, TNNC1, TNNT2, TTN*). LOFTEE was used to identify low-confidence pLoF variants and identify PAVs that are mislabelled (e.g. NAGNAG sites). The variants were filtered for disease-causing mechanisms; all variants in *BAG3, LMNA, PLN, SCN5A, RBM20, and DSP*, PAVs were kept; only variants influencing gene product structure (indels, missense, start and stop lost) or gene product level but NMD escaping, were kept for *MYH7, DES, TNNC1, and TNNT2*; only variants predicted to influencing gene product level (e.g. frameshift, splice, stop gained) were kept for *TTN and FLNC*. Step 2: Carriers of the 7,549 variants remaining include a list of individuals that carry a DCM indeterminant variant. After exclusion for HCM variants (see Step3), 62,292 carriers were identified for the 7,467 variants, of which 56,815 were heterozygotes, 5,404 were compound heterozygotes, 72 were homozygotes (19 of which were also heterozygous for other variants) and 1 compound homozygote.

The variant list was then shortened to only include variants that met a filtering allele frequency (FAF) of <0.000084 in gnomAD pop max faf and as a MAF in UK Biobank. Additionally, compound heterozygous carriers of common TTNtvs (same two TTN variants identified in >10 individuals) were removed from the analysis as these are likely rescued. Step 2b: Carriers of the 6,950 variants remaining include a list of individuals that carry a DCM indeterminant variant that is rare enough to have a substantial effect on the disease. After exclusion for HCM variants (see Step3), 26,181 carriers were identified for the 6,868 variants, of which 25,207 were heterozygotes, 957 were compound heterozygotes, and 17 were homozygotes (1 of which was also heterozygous for another variant).

The variant list was then shortened to only include variants that would be called pathogenic/likely pathogenic if identified in a patient with DCM; using ClinVar, the variants were manually curated if they had any evidence of pathogenicity. 1,351 variants were assessed, of which 1,018 had strong evidence of pathogenicity in DCM and 82 were identified with evidence of HCM. These 82 were excluded from Step 2 onwards. Carriers of the 1,018 variants remaining include a list of individuals that carry a DCM pathogenic/likely pathogenic variant. 2,195 carriers were identified for the 1,018 variants, of which 2,181 were heterozygotes, 14 were compound heterozygotes, and there were no homozygotes.

#### Details of physical activity

Physical activity increased fractal dimension (**Figure S2**). Summed metabolic equivalent (MET) minutes per week for all activity (data ID 22040.0.0, highest compared to lowest 10%, R=0.03; $\beta$=0.11, SE=0.01, *P*=2.157e-05), for vigorous activity (data ID 22039.0.0, R=0.04; $\beta$=0.13, SE=0.01, *P*=1.904e-09), and above moderate/vigorous recommendation (data ID 22035.0.0, $\beta$=0.08, SE=0.01, *P*=7.571e-12), as well as reported exercise via questionnaire (days per week (0 days versus 7 days); moderate exercise $\beta$=0.11, SE=0.02, *P*=6.677e-09; vigorous exercise $\beta$=0.19, SE=0.03, *P*=1.293e-07) and overall acceleration average via accelerometer data (R=0.06; $\beta$=0.24, SE=0.01, $P$=1.166e-12, (**Figure S2**)), associated with increased trabeculation, with the effect more substantial for activity deemed vigorous. The association with physical activity was in part due to LVEDV, which has been shown previously to predict exercise capacity[6]. Adjustment for LVEDV retained significance only for vigorous exercise (reported via questionnaire as days per week (0 days versus 7 days), $\beta$=0.11, SE=0.05, $P$=0.0269).

#### Details of curated cardiovascular traits

Curated cardiovascular trait association analysis showed that mean global fractal dimension was increased in participants diagnosed with cardiomyopathies (HCM ($\beta$=0.95, SE=0.17, $P$=8.573e-05, n=36 diagnosed), DCM ($\beta$=0.74, SE=0.16, $P$=0.01071, n=41), mixed cardiomyopathy ($\beta$=0.65, SE=0.09, $P$=1.448e-08, n=133), non-ischemic cardiomyopathy ($\beta$=0.22, SE=0.01, $P$=9.177e-12, n=1,572)), heart failure ($\beta$=0.24, SE=0.05, $P$=6.138e-04, n=409), coronary disease ($\beta$=0.08, SE=0.03, $P$=0.01151, n=1,728), fibrillation and flutter ($\beta$=0.08, SE=0.03, $P$=0.04195, n=1,730), and congenital heart disease (ICD10 Q20-Q28; $\beta$=0.37, SE=0.09, $P$=0.002172, n=137). The relationship between trabeculation and HCM was independent of LVEDV ($\beta$=0.87, SE=0.17, $P$=4.877e-04) and wall thickness ($\beta$=1.07, SE=0.17, $P$=1.952e-05). The relationship between trabeculation and DCM was not independent of LVEDV ($P$=0.6136). Adjustment for LVEDV identified a significant association with diabetes ($\beta$=0.08, SE=0.02, $P$=0.01194).

#### Details of relationship with risk markers

The relationship between mean global fractal dimension and risk markers is strengthened in participants diagnosed with DCM (**Figure** 28): trabeculation correlated with decreased measures of ejection fraction (LVEF, RVEF, LAEF, RAEF, R=-0.53--0.25) and increased strain and volumes (LVEDV, LVESV, R=0.63-0.67; global Err R=-0.54; global Ecc, global Ell, radial PDSR, R=0.26-0.47). Adjustment for LVEDV identified a negative relationship between trabeculation and cardiac output (LVCO; R=-0.52). The relationship between mean global fractal dimension and markers of cardiac structure and function in participants diagnosed with HCM identified a positive relationship with measures of wall thickness (R=0.33-0.40) and a negative relationship with volume in the right ventricle (R=-0.14--0.17), independent of LVEDV (**Figure S**29).

#### Full discussion of Mendelian randomization

To assess causality we used two-sample Mendelian randomization (MR) for association testing of genetically-predicted (or proxied) levels of an exposure with an outcome[7]. A genetic variant is considered an instrumental variable for a given exposure if it i) associated with the exposure, ii) not associated with the outcome due to confounding pathways, and iii) it does not affect the outcome except via the exposure[7]. Assessments of causality via MR for trabeculation are complex as trabeculation and cardiomyopathies are genetically and phenotypically overlapping (iii) and sometimes driven through LVEDV (ii). We do not believe that a variant identified in association with measures of trabeculation in the locus of cardiomyopathy-associated genes (e.g., *TTN*) causes cardiomyopathy or heart failure only through trabeculation (but rather an alteration in sarcomeric function, etc.). To overcome this, it has been suggested that one can prune potentially pleiotropic variants from an MR analysis but variant exclusion is difficult due the overlap of trabeculation-associated variants with those associated with inherited cardiac conditions. Selection is complicated by the availability, limited sample size, and diagnostic heterogeneity, of current case/control GWAS of inherited cardiac conditions; quantitative measures of trabeculation in nearly 50,000 participants are likely more statistically powered than case/control GWAS to date. Regardless, the genetically-predicted mean global fractal dimension was not a statistically significant exposure for cardiomyopathies or heart failure and efforts to prune cardiomyopathy-associated variants did not suggest causality. We tested a hypothesis that the observed hypertrabeculation for opposing HCM and DCM-associated variants may cancel out in combination at MR of trabeculation as exposure for cardiomyopathy outcomes. Many mean global fractal dimension genetic instruments were opposing for HCM and DCM but their removal remained non-significant.

We believe the converse may be true: genetic variants causing cardiomyopathy may alter trabeculation alongside the progression of cardiomyopathy. MR of cardiomyopathy as an exposure for trabeculation outcome was undertaken for HCM, DCM, and heart failure (HF; **Figure** 31, **Table S6**)[8–10]. The inferences of causality depended on LVEDV: adjustment of mean global fractal dimension for LVEDV at GWAS resulted in non-significant MR results using the resulting summary statistics (**Table S6**). Genetic proxied DCM and heart failure (with weaker significance) had a positive relationship with proxied trabeculation, although the Steiger test for directionality suggested it was the incorrect causal direction. The association with HF was driven through an intronic variant in the p53 cell division arrest protein target; p21 cyclin-dependent kinase inhibitor (*CDKN1A*; rs3176326). This SNP was also a MR genetic instrument for both DCM and HCM. The A allele decreases *CDKN1A* expression (eQTL on GTEx; NES=-0.12, P=0.000014), DCM, HF, and trabeculation (GWAS betas= -0.07, -0.07, and -0.05, respectively and regardless of LVEDV; P<$5x10^{-8}$) through increased cell division (and increased HCM risk; beta= 0.34, P<$5x10^{-8}$)[11].

Genetic proxied HCM had a negative relationship with proxied trabeculation suggesting that variants associated with HCM decrease trabeculation. The polygenic relationships between HCM, DCM, and HF, were assessed to help understand this finding: genetically-derived HCM and DCM have a significant, opposing relationship. This was previously identified; the same genetic pathways can lead to distinct disorders through opposing genetic effects[11]. The Steiger test for directionality suggested that genetically derived HCM is protective for DCM. Furthermore, genetic instruments for DCM were significant risk factors for HF but genetic instruments for HCM were protective risk factors for HF, likely driven by measures of volume.

As several associated variants have opposite associations with HCM and DCM, this finding suggests that shared biological processes contribute to trabeculation in both diseases, but through opposing mechanisms. The observed hypertrabeculation and increased left ventricular complexity progressing in parallel with ventricular volume in DCM and HF may be compensatory and adaptive in HCM compared to a less structurally complex ventricular wall.

### List of Figures

1 Association of covariates with mean global trabeculation. Covariate-adjusted mean global trabeculation measured by fractal dimension analysis was compared to age at scan, age$^2$, systolic blood pressure (SBP), body surface area (BSA), sex (0=female, 1=male), and imaging batch (three imaging centres). . . . . . . . . . . . . 9

2 Association of alcohol intake and average acceleration with mean global trabeculation. Covariate-adjusted mean global trabeculation measured by fractal dimension analysis was compared to **A)** alcohol intake and the lowest 10% of alcohol drinkers had significantly decreased trabeculation. The comparison with **B)** showed that trabeculation had a positive relationship with acceleration measures via accelerometer. . . . . . . . . . 10

3 Adjustment for LVEDV makes little difference to the meta Manhattan plot of the minimum p-value for all SNPs across all TM measures. The Manhattan plots depict the p-value of association (x-axis, -log10(p-value) and the SNPs across 22 chromosomes (y-axis) from a GWAS of 38,245 participants of the UK Biobank population. 10

4 Manhattan and quantile-quantile plots of the genome-wide association analyses of trabeculation traits. The Manhattan plots depict the p-value of association (x-axis, -log10(p-value) and the SNPs across 22 chromosomes (y-axis). The quantile-quantile (QQ) plots depict the expected versus observed p-values for each GWAS. Please see the inflation factor estimates (lambda) in the supplementary tables. . . . . . . . . . . . . . . . . . . . . . 11

5 Manhattan and quantile-quantile plots of the genome-wide association analyses of trabeculation traits. The Manhattan plots depict the p-value of association (x-axis, -log10(p-value) and the SNPs across 22 chromosomes (y-axis). The quantile-quantile (QQ) plots depict the expected versus observed p-values for each GWAS. Please see the inflation factor estimates (lambda) in the supplementary tables. . . . . . . . . . . . . . . . . . . . . . 12

6 Manhattan and quantile-quantile plots of the genome-wide association analyses of trabeculation traits. The Manhattan plots depict the p-value of association (x-axis, -log10(p-value) and the SNPs across 22 chromosomes (y-axis). The quantile-quantile (QQ) plots depict the expected versus observed p-values for each GWAS. Please see the inflation factor estimates (lambda) in the supplementary tables. . . . . . . . . . . . . . . . . . . . . . 13

7 Manhattan and quantile-quantile plots of the genome-wide association analyses of trabeculation traits. The Manhattan plots depict the p-value of association (x-axis, -log10(p-value) and the SNPs across 22 chromosomes (y-axis). The quantile-quantile (QQ) plots depict the expected versus observed p-values for each GWAS. Please see the inflation factor estimates (lambda) in the supplementary tables. . . . . . . . . . . . . . . . . . . . . . 14

8 Manhattan and quantile-quantile plots of the genome-wide association analyses of trabeculation traits. The Manhattan plots depict the p-value of association (x-axis, -log10(p-value) and the SNPs across 22 chromosomes (y-axis). The quantile-quantile (QQ) plots depict the expected versus observed p-values for each GWAS. Please see the inflation factor estimates (lambda) in the supplementary tables. . . . . . . . . . . . . . . . . . . . . . 15

9 Manhattan and quantile-quantile plots of the genome-wide association analyses of trabeculation traits. The Manhattan plots depict the p-value of association (x-axis, -log10(p-value) and the SNPs across 22 chromosomes (y-axis). The quantile-quantile (QQ) plots depict the expected versus observed p-values for each GWAS. Please see the inflation factor estimates (lambda) in the supplementary tables. . . . . . . . . . . . . . . . . . . . . . 16

10 Manhattan and quantile-quantile plots of the genome-wide association analyses of trabeculation traits. The Manhattan plots depict the p-value of association (x-axis, -log10(p-value) and the SNPs across 22 chromosomes (y-axis). The quantile-quantile (QQ) plots depict the expected versus observed p-values for each GWAS. Please see the inflation factor estimates (lambda) in the supplementary tables. . . . . . . . . . . . . . . . . . . . . . 17

11 Manhattan and quantile-quantile plots of the genome-wide association analyses of trabeculation traits. The Manhattan plots depict the p-value of association (x-axis, -log10(p-value) and the SNPs across 22 chromosomes (y-axis). The quantile-quantile (QQ) plots depict the expected versus observed p-values for each GWAS. Please see the inflation factor estimates (lambda) in the supplementary tables. . . . . . . . . . . . . . . . . . . . . . 18

12 Manhattan and quantile-quantile plots of the genome-wide association analyses of trabeculation traits. The Manhattan plots depict the p-value of association (x-axis, -log10(p-value) and the SNPs across 22 chromosomes (y-axis). The quantile-quantile (QQ) plots depict the expected versus observed p-values for each GWAS. Please see the inflation factor estimates (lambda) in the supplementary tables. . . . . . . . . . . . . . . . . . . . . . 19

13 Manhattan and quantile-quantile plots of the genome-wide association analyses of trabeculation traits. The Manhattan plots depict the p-value of association (x-axis, -log10(p-value) and the SNPs across 22 chromosomes (y-axis). The quantile-quantile (QQ) plots depict the expected versus observed p-values for each GWAS. Please see the inflation factor estimates (lambda) in the supplementary tables. . . . . . . . . . . . . . . . . . . . . . 20

14 Manhattan and quantile-quantile plots of the genome-wide association analyses of trabeculation traits. The Manhattan plots depict the p-value of association (x-axis, -log10(p-value) and the SNPs across 22 chromosomes (y-axis). The quantile-quantile (QQ) plots depict the expected versus observed p-values for each GWAS. Please see the inflation factor estimates (lambda) in the supplementary tables. . . . . . . . . . . . . . . . . . . . . . 21

15 Manhattan and quantile-quantile plots of the genome-wide association analyses of trabeculation traits. The Manhattan plots depict the p-value of association (x-axis, -log10(p-value) and the SNPs across 22 chromosomes (y-axis). The quantile-quantile (QQ) plots depict the expected versus observed p-values for each GWAS. Please see the inflation factor estimates (lambda) in the supplementary tables. . . . . . . . . . . . . . . . . . . . . 22
16 Manhattan and quantile-quantile plots of the genome-wide association analyses of trabeculation traits. The Manhattan plots depict the p-value of association (x-axis, -log10(p-value) and the SNPs across 22 chromosomes (y-axis). The quantile-quantile (QQ) plots depict the expected versus observed p-values for each GWAS. Please see the inflation factor estimates (lambda) in the supplementary tables. . . . . . . . . . . . . . . . . . . . . 23
17 Increase in mean global trabeculation with variant pathogenicity. These independent groups of individuals for **A)** HCM and **B)** DCM, were separated by carrier status. For HCM-associated variants, there were 28,006 genotype-negative participants (Negative), 6,040 participants with variants in syndromic genes (Positive1), 1,330 participants with common variants (AF<0.01 & >0.00004) in definitive-evidence genes (Positive2A), 680 participants with rare indeterminant (VUS) variants in definitive-evidence genes (Positive2B - significant), 72 carriers of P/LP variants (Positive3 - significant). For DCM-associated variants, there were 28,109 genotype-negative participants (Negative), 3,379 participants with variants in syndromic genes (Positive1), 2,710 participants with common variants (AF<0.01 & >0.000084) in definitive-evidence genes (Positive2A), 1,837 participants with rare indeterminant variants in definitive-evidence genes (Positive2B), 162 carriers of P/LP variants (Positive 3 - significant). Means were compared against the genotype-negative group. . . . . . . . . 24
18 PheWAS for summary trabeculation measures when adjusted for LVEDV. . . . . . . . . . . . . . . . . . . . . 25
19 Mendelian randomization analysis of HF as exposure on mean global FD. The plots show summary information on the analyses, performed as per the TwoSampleMR R package. . . . . . . . . . . . . . . . . . . . . . . . . 26
20 Mendelian randomization analysis of DCM as exposure on trabeculation outcome. The plots show summary information on the analyses, performed as per the TwoSampleMR R package. . . . . . . . . . . . . . . . . . 27
21 Mendelian randomization analysis of DCM as exposure on trabeculation outcome, pruning for an NFIA variant. The plots show summary information on the analyses, performed as per the TwoSampleMR R package. . . . 28
22 Mendelian randomization analysis of for HCM as exposure on trabeculation outcome. The plots show summary information on the analyses, performed as per the TwoSampleMR R package. . . . . . . . . . . . . . . . . . 29
23 Mendelian randomization analysis of for HCM as exposure on HF outcome. The plots show summary information on the analyses, performed as per the TwoSampleMR R package. . . . . . . . . . . . . . . . . . . . . . 30
24 Mendelian randomization analysis of DCM as exposure on HF outcome. The plots show summary information on the analyses, performed as per the TwoSampleMR R package. . . . . . . . . . . . . . . . . . . . . . . . . 31
25 Mendelian randomization analysis of DCM and HCM as exposures on HF outcome. . . . . . . . . . . . . . . 32
26 Mendelian randomization analysis of HCM as exposure on DCM outcome. The plots show summary information on the analyses, performed as per the TwoSampleMR R package. . . . . . . . . . . . . . . . . . . . . . . . . 33
27 Mendelian randomization analysis of DCM as exposure on HCM outcome. The plots show summary information on the analyses, performed as per the TwoSampleMR R package. . . . . . . . . . . . . . . . . . . . . . . . . 34
28 Association of trabecular morphology with ancestry and cardiac MRI-derived summary measures. The analyses were completed on 38,245 participants of the UK Biobank population. a) Compared to British ancestry which dominates the UK Biobank, the mean global fractal dimension was increased for participants of African ancestry. Indian, Chinese, Bangladeshi, and "Any other white background" self-reported ancestry had the lowest significant FD. Student's two-sided t-test was used to compare means. b) The table quantifies the relationship (correlation coefficient, R) between mean global fractal dimension and 2D summary imaging cardiac measures separately for participants with no diagnosis (n=31,067), hypertrophic cardiomyopathy (HCM, n=31), dilated cardiomyopathy (DCM, n=29), and heart failure (n=332). . . . . . . . . . . . . . . . . 35
29 The relationship of mean global FD adjusted for LVEDV with summary CMR measures in participants with cardiomyopathy or heart failure. . . . . . . . . . . . . . . . . . . . . . . . . . . . . . . . . . . . . . . . . 36
30 Outcome associations. The analyses were completed on 47,803 participants of the UK Biobank population and assessed from years since imaging for a diagnosis of a cardiac condition or all-cause death, with anyone diagnosed before imaging excluded. Trabeculation was separated into three groups: hyper-, median, and hypotrabeculation where <1.5 SD is hypotrabeculation and >1.5 SD is hypertrabeculation. a) The hazard ratio for heart failure, for mitral valve disorders, and bundle branch block, showed increased risk with hypertrabeculation. b) The hazard ratio with the adjustment for LVEDV (and removal of the association of trabeculation with DCM) showed that hypotrabeculation was associated with the risk of non-DCM heart failure. c) The hazard ratio for assessments with participants diagnosed with heart failure only showed that heart failure risk increased with hypotrabeculation. d) The mean coxfit linear predictors for heart failure were plotted for trabeculation by decile. Of those with heart failure or death, participants with hypotrabeculation were diagnosed or died nearly a year later on average. Confidence intervals and log-rank p-values are depicted. CI, concordance index. . . . . . . . . . . . . . . . . . . . . . . . . . . . . . . . . . . . . . . . . . . . . . . 37

31 Selected outcome associations. **a**, The analyses were completed on 47,803 participants of the UK Biobank population and assessed from years since imaging for a diagnosis of a cardiac condition or all-cause death, with anyone diagnosed before imaging excluded. Trabeculation was separated into three groups: hyper-, median, and hypotrabeculation where <-1.5 SD is hypotrabeculation and >1.5 SD is hypertrabeculation, where hypotrabeculation and hypertrabeculation were compared to the median trabeculation (dashed line). The hazard ratio for heart failure, mitral valve disorders, and bundle branch block, showed increased risk with hypertrabeculation. All cause death was also assessed but was not significant. The hazard ratio with the adjustment for LVEDV (and removal of the association of trabeculation with DCM) showed that hypotrabeculation was associated with risk of non-DCM heart failure. The hazard ratio for assessments with participants diagnosed with heart failure only showed that heart failure risk increased with hypotrabeculation. Point estimates with 95% confidence intervals (CI) are shown. *, P<0.05; **, P<0.01; ***, P<0.001; ****, P<0.0001. **b**, Carriers of cardiomyopathy-associated pathogenic or likely pathogenic variants (G+) with or without a diagnosis of disease (phenotype; P+/-) had increased mean trabeculation compared to genotype negatives. There were 23,936 genotype-negative participants, 222 participants with a P/LP variant without a diagnosis of cardiomyopathy, 63 participants with disease and no variant in definitive-evidence genes, and 12 participants with a variant and diagnosis of disease. Student's two-sided t-test was used to compare the group means. The lower and upper hinges in the box plot correspond to the 25th and 75th percentiles (interquartile range (IQR)), respectively. The horizontal line in the box plot indicates the median. The lower and upper whiskers extend from the hinge to the smallest and largest values no further than 1.5× the IQR, respectively. Each dot is one individual. P-G+, phenotype negative, genotype positive; P+G-, phenotype positive, genotype negative; P+G+, phenotype positive, genotype positive. . . . . . . 38
32 PheWAS for summary trabeculation without aggregation. . . . . . . 39

### List of Tables

1 Model information from the multiple linear regression of covariates when adjusting mean global trabeculation. PC, genetic principal component. . . . 40

2 Genetic covariance ($cov_g$) and correlation ($r_g$) with mean global trabeculation. Mean global trabeculation had a heritability estimate of 43%. The estimates are shown with and without adjustment for covariates. The analyses were completed on 38,245 participants of the UK Biobank population. . . . 40

3 Summary information on the two groups of imaged UK Biobank participants and heritability estimates for trabeculation measures. Slice, imaging slice; Min, minimum; Med, median; FD, fractal dimension measure of trabeculation; SD, standard deviation; SE, standard error. . . . 41

4 Inflation factor from genome-wide association studies. Slice, image slices. . . . 42

5 **See excel file for extended Table S5.** The table depicts the independent significant GWAS loci for trabecular measures as observed from LocusZoom. name, random ID of independent locus; nocmhf?, whether the locus was also observed when participants in the UK Biobank with cardiomyopathy or heart failure were removed; nocm?, whether the locus was also observed when participants in the UK Biobank with cardiomyopathy were removed; Region, position of locus in GrCh38; Gene1-3; nearest genes from LocusZoom. . . . 42

6 **See excel file for extended Table S6.** Summary of Mendelian randomisation results. The table depicts the results for trabeculation as outcome and exposure with dilated cardiomyopathy (DCM), hypertrophic cardiomyopathy (HCM), and heart failure (HF). Five methods were used to assess for causality, with the number of SNPs included in the analysis (nsnp), beta effect size (b), standard error (se), and p-value (pval) shown. . . 43

7 **See excel file for extended Table S7.** The table depicts the validated rare variant association studies results for trabecular measures. CHROM, chromosome; GENPOS, gene position; ALLELE1, masks; A1FREQ, allele frequency; N, sample size of discovery cohort; TEST, RVAS method analysed by Regenie software; SE, standard error; LOG10P, p-value of association. . . . 43

8 **See excel file for extended Table S8.** The table depicts the phenome-wide association studies results for imaging and trabecular measures. SE, standard error; OR, odds ratio; p, p-value of association; n_total phenotype, number of participants with imaging measure. . . . 43

9 **See excel file for extended Table S9.** The table depicts the novel, independent, significant GWAS loci for trabecular measures for both validation and discovery datasets as observed from LocusZoom. name, random ID of independent locus; Region, position of locus in GrCh38; Gene1-3; nearest genes from LocusZoom. . . 43

10 **See excel file for extended Table S10.** The table depicts the novel, rare variant association studies results for trabecular measures using both discovery and validation cohorts. CHROM, chromosome; GENPOS, gene position; ALLELE1, masks; A1FREQ, allele frequency; N, sample size of discovery cohort; TEST, RVAS method analysed by Regenie software; SE, standard error; LOG10P, p-value of association. . . . 43

11 **See excel file for extended Table S11.** Significant transcriptome-wide association study results for trabecular measures using both discovery and validation cohorts for cardiac tissues only (left ventricle and atrial appendage). . . . 43

12 **See excel file for extended Table S12.** Significant transcriptome-wide association study results for trabecular measures using both discovery and validation cohorts. . . . 43

13 **See excel file for extended Table S13.** Significant Gene Ontology (GO) Resource Enrichment analysis results using Panther on all genes in Tables S5 and S9. A PANTHER overrepresentation test (Released 20231017) was performed on the GO Ontology human database (doi:10.5281/zenodo.10536401 Released 2024-01-17). Significance was assessed via fisher exact test with an FDR correction. The GO gene lists assessed are shown in bold. . . . 43

#### Supplementary figures

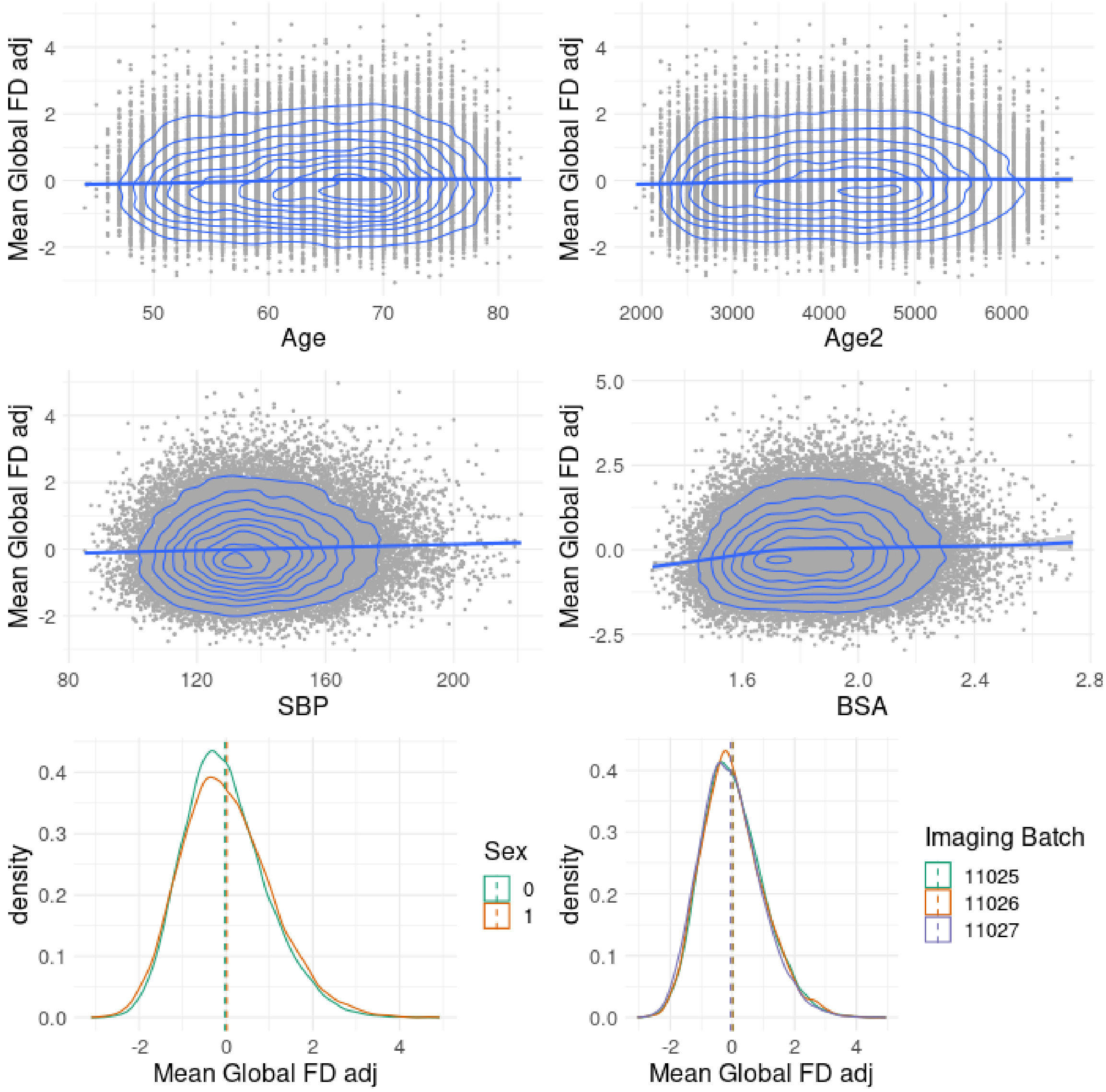

**Figure S 1.** Association of covariates with mean global trabeculation. Covariate-adjusted mean global trabeculation measured by fractal dimension analysis was compared to age at scan, age$^2$, systolic blood pressure (SBP), body surface area (BSA), sex (0=female, 1=male), and imaging batch (three imaging centres).

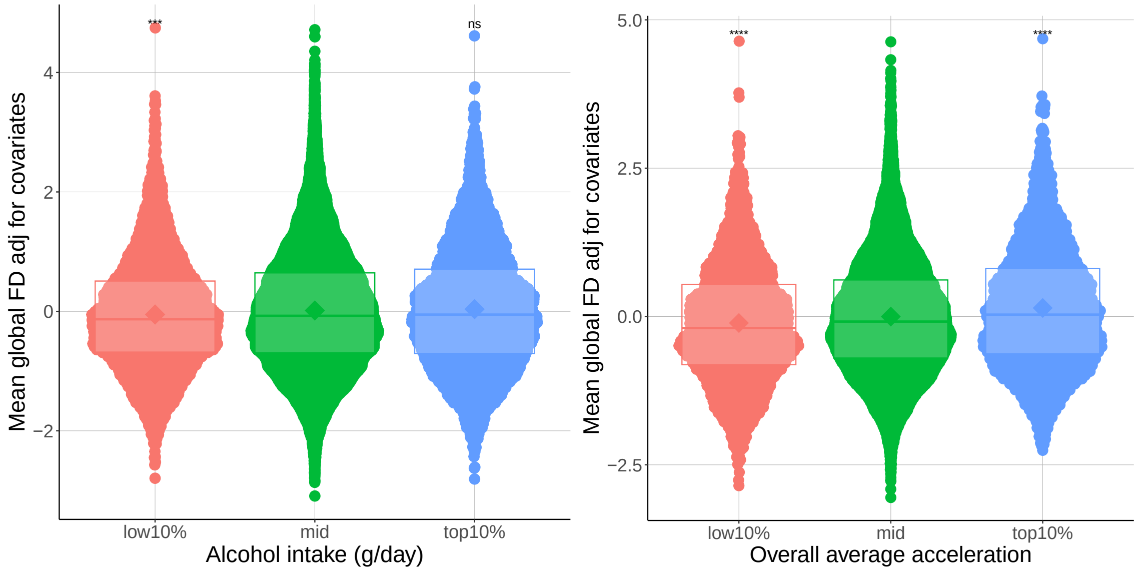

**(a)** Lowest alcohol drinkers have decreased cardiac trabeculation.

**(b)** Cardiac trabeculation has a positive relationship with average acceleration.

**Figure S 2.** Association of alcohol intake and average acceleration with mean global trabeculation. Covariate-adjusted mean global trabeculation measured by fractal dimension analysis was compared to **A)** alcohol intake and the lowest 10% of alcohol drinkers had significantly decreased trabeculation. The comparison with **B)** showed that trabeculation had a positive relationship with acceleration measures via accelerometer.

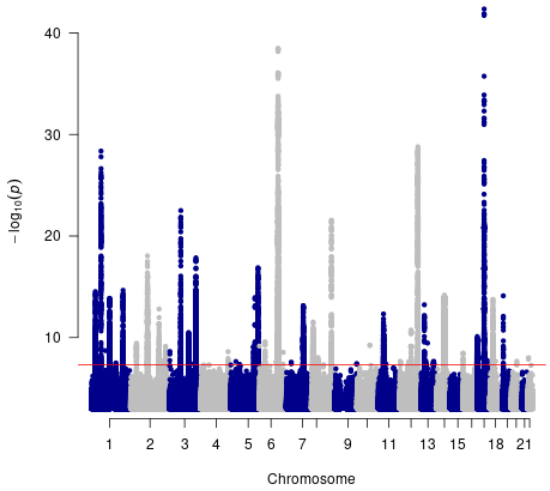

**Figure S 3.** Adjustment for LVEDV makes little difference to the meta Manhattan plot of the minimum p-value for all SNPs across all TM measures. The Manhattan plots depict the p-value of association (x-axis, -log10(p-value) and the SNPs across 22 chromosomes (y-axis) from a GWAS of 38,245 participants of the UK Biobank population.

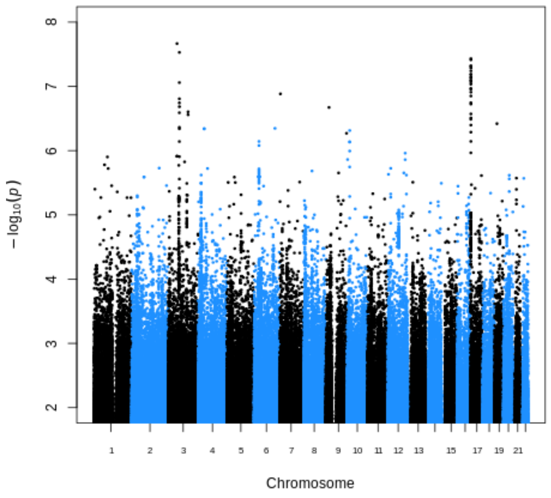

**(a)** Slice 1 Manhattan plot

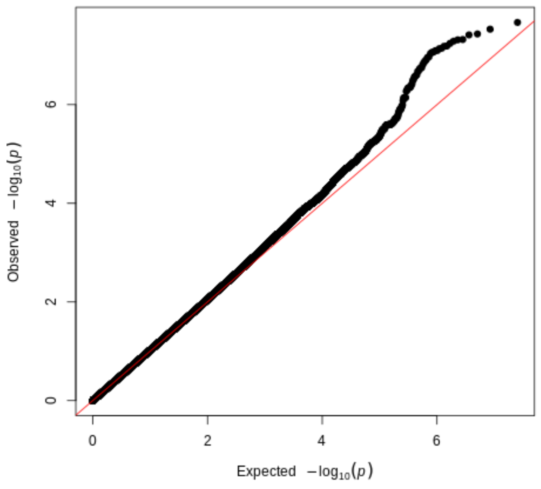

**(b)** Slice 1 quantile-quantile plot

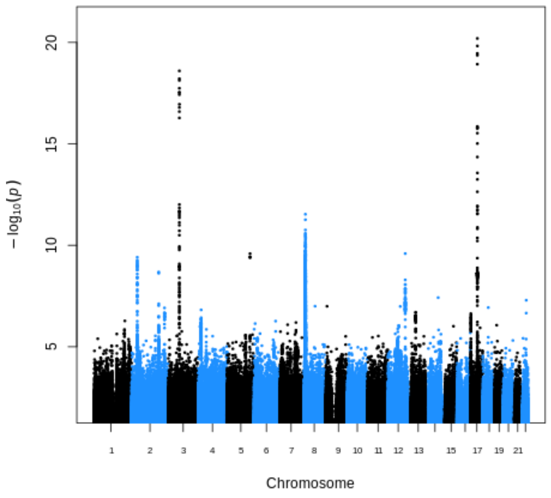

**(c)** Slice 2 Manhattan plot

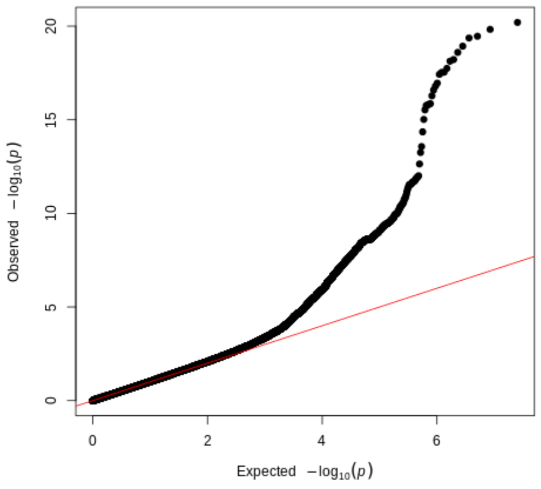

**(d)** Slice 2 quantile-quantile plot

**Figure S 4.** Manhattan and quantile-quantile plots of the genome-wide association analyses of trabeculation traits. The Manhattan plots depict the p-value of association (x-axis, -log10(p-value) and the SNPs across 22 chromosomes (y-axis). The quantile-quantile (QQ) plots depict the expected versus observed p-values for each GWAS. Please see the inflation factor estimates (lambda) in the supplementary tables.

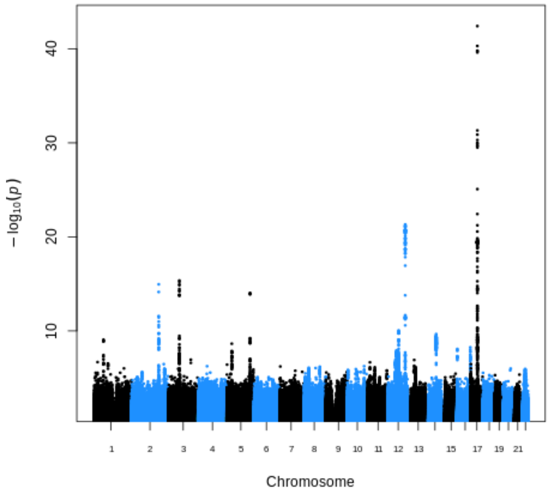

**(a)** Slice 3 Manhattan plot

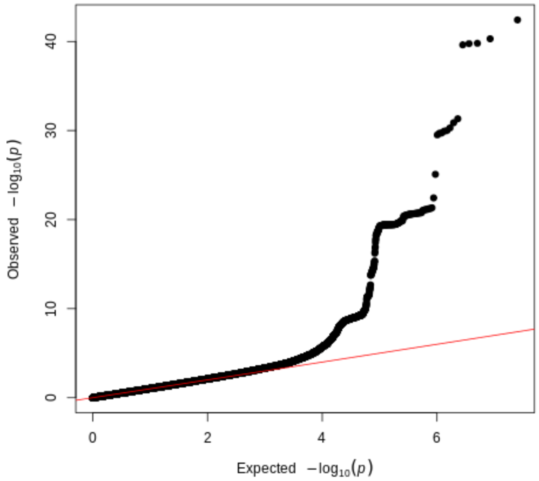

**(b)** Slice 3 quantile-quantile plot

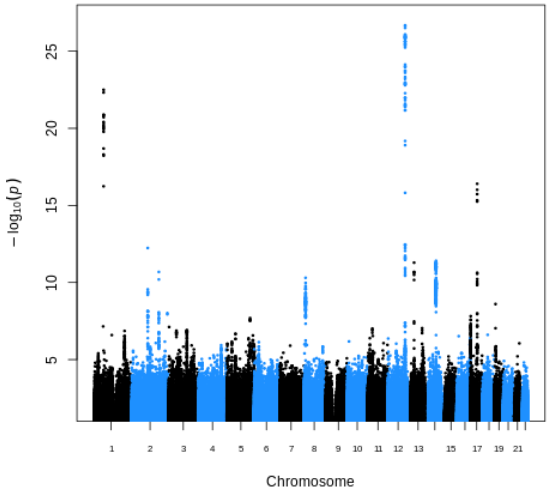

**(c)** Slice 4 Manhattan plot

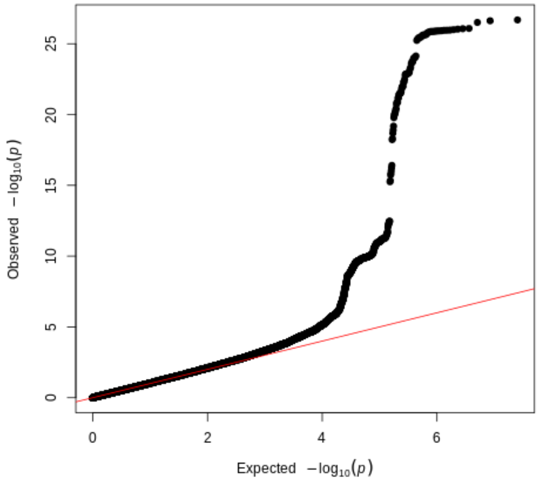

**(d)** Slice 4 quantile-quantile plot

**Figure S 5.** Manhattan and quantile-quantile plots of the genome-wide association analyses of trabeculation traits. The Manhattan plots depict the p-value of association (x-axis, -log10(p-value) and the SNPs across 22 chromosomes (y-axis). The quantile-quantile (QQ) plots depict the expected versus observed p-values for each GWAS. Please see the inflation factor estimates (lambda) in the supplementary tables.

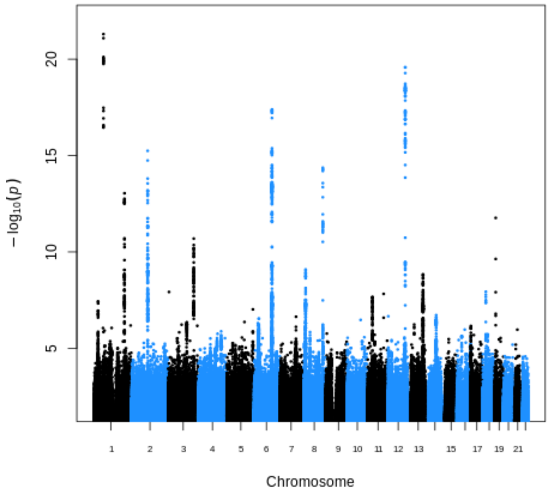

**(a)** Slice 5 Manhattan plot

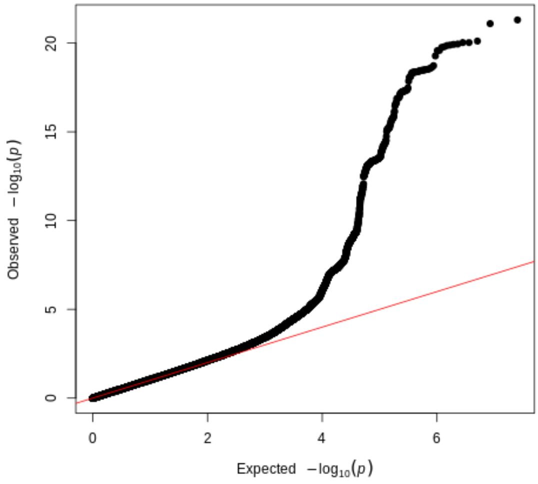

**(b)** Slice 5 quantile-quantile plot

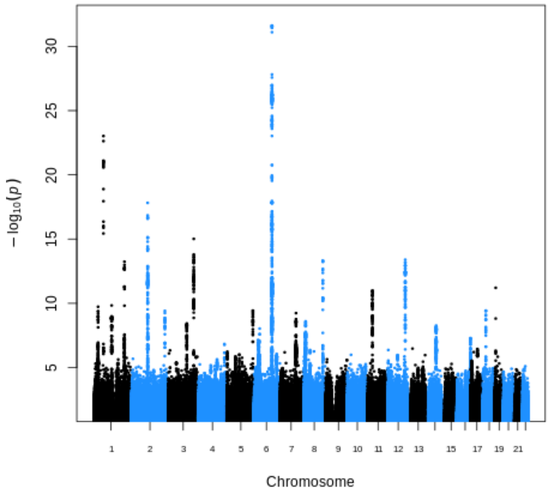

**(c)** Slice 6 Manhattan plot

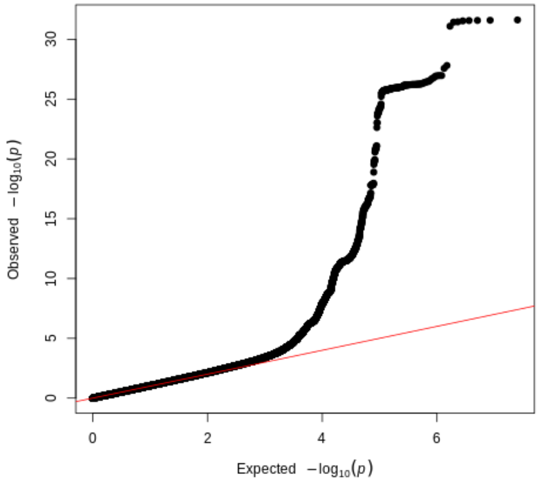

**(d)** Slice 6 quantile-quantile plot

**Figure S 6.** Manhattan and quantile-quantile plots of the genome-wide association analyses of trabeculation traits. The Manhattan plots depict the p-value of association (x-axis, -log10(p-value) and the SNPs across 22 chromosomes (y-axis). The quantile-quantile (QQ) plots depict the expected versus observed p-values for each GWAS. Please see the inflation factor estimates (lambda) in the supplementary tables.

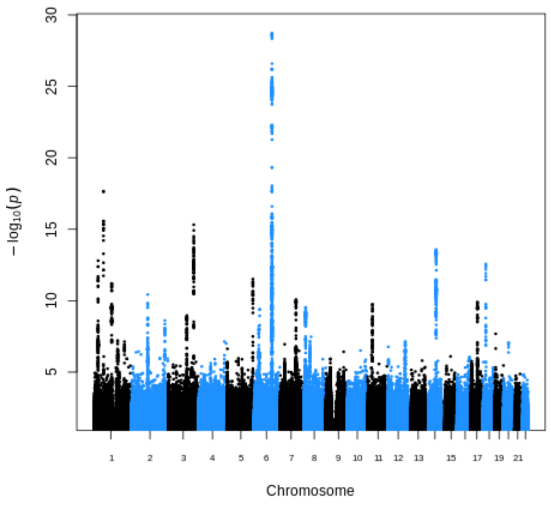

**(a)** Slice 7 Manhattan plot

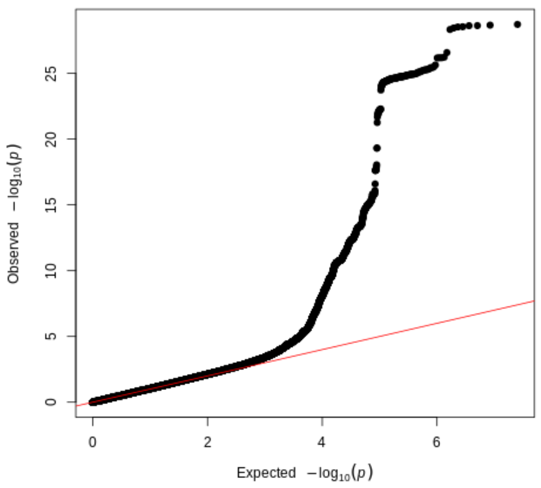

**(b)** Slice 7 quantile-quantile plot

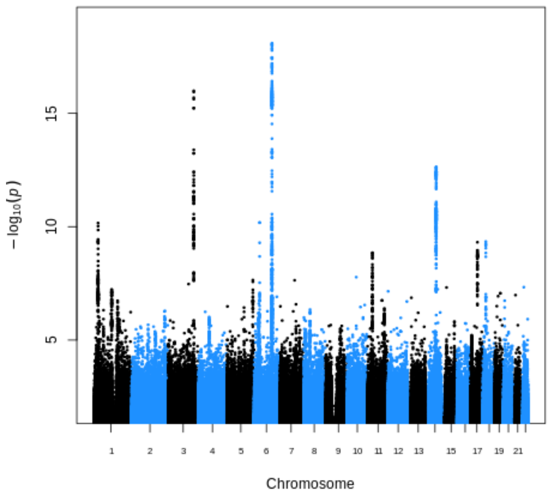

**(c)** Slice 8 Manhattan plot

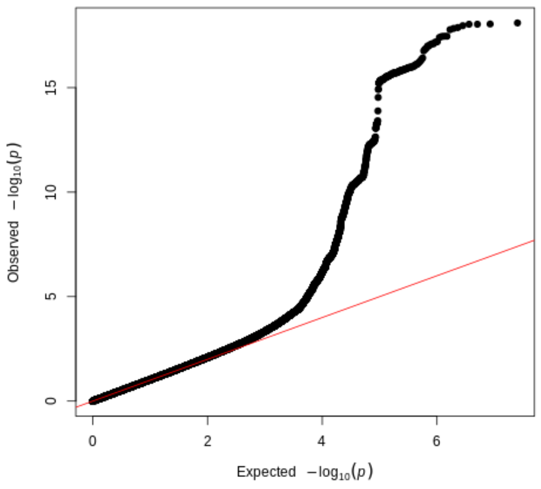

**(d)** Slice 8 quantile-quantile plot

**Figure S 7.** Manhattan and quantile-quantile plots of the genome-wide association analyses of trabeculation traits. The Manhattan plots depict the p-value of association (x-axis, -log10(p-value) and the SNPs across 22 chromosomes (y-axis). The quantile-quantile (QQ) plots depict the expected versus observed p-values for each GWAS. Please see the inflation factor estimates (lambda) in the supplementary tables.

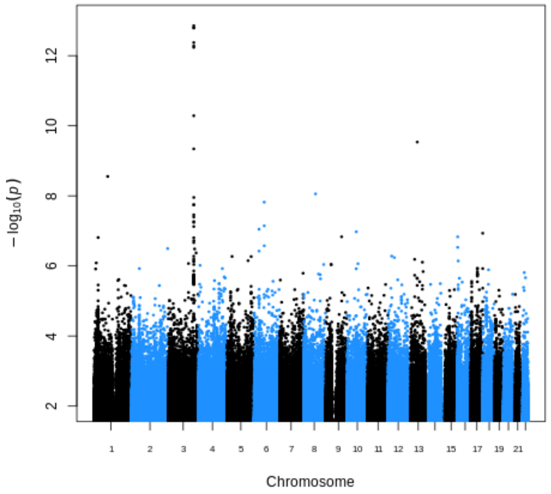

**(a)** Slice 9 Manhattan plot

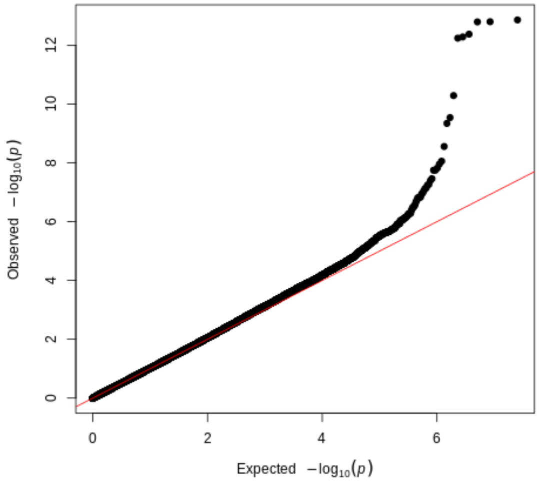

**(b)** Slice 9 quantile-quantile plot

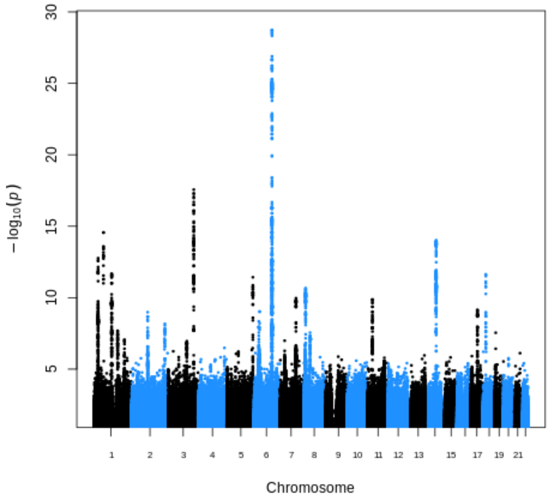

**(c)** Max apical Manhattan plot

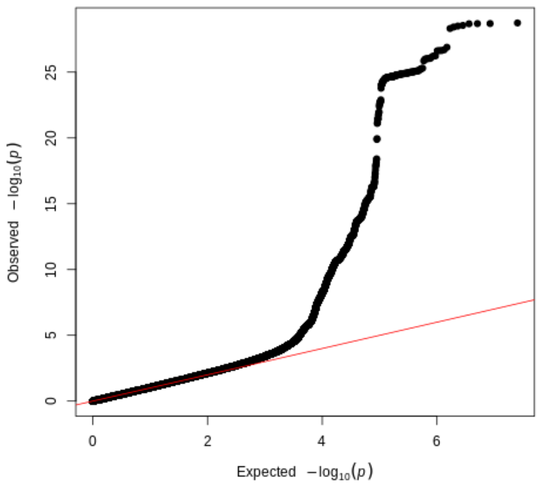

**(d)** Max apical quantile-quantile plot

**Figure S 8.** Manhattan and quantile-quantile plots of the genome-wide association analyses of trabeculation traits. The Manhattan plots depict the p-value of association (x-axis, -log10(p-value) and the SNPs across 22 chromosomes (y-axis). The quantile-quantile (QQ) plots depict the expected versus observed p-values for each GWAS. Please see the inflation factor estimates (lambda) in the supplementary tables.

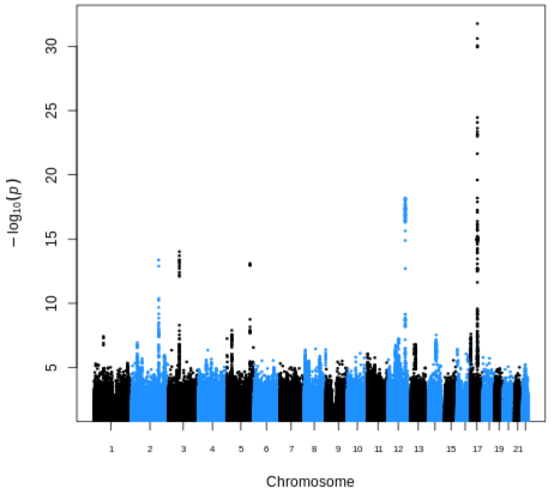

**(a)** Max basal Manhattan plot

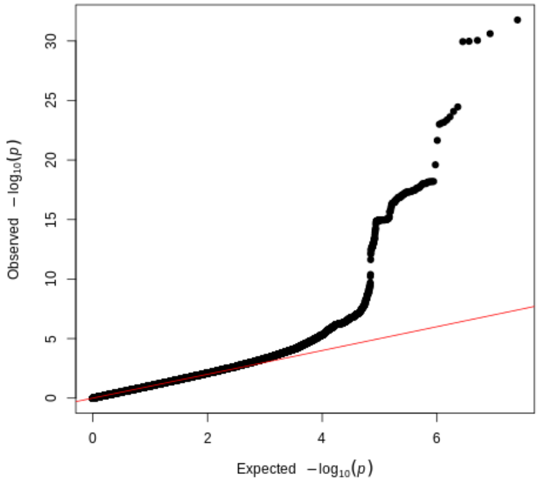

**(b)** Max basal quantile-quantile plot

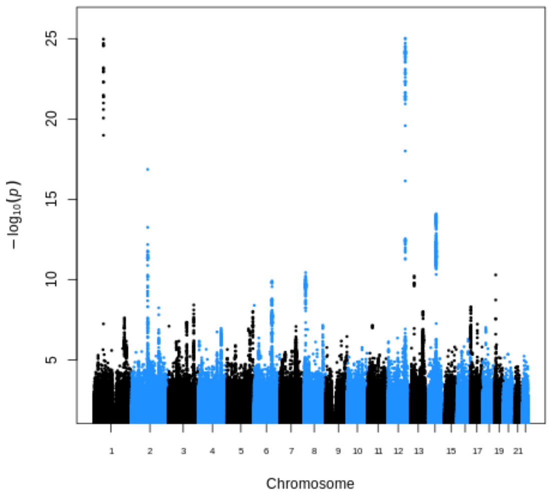

**(c)** Max global Manhattan plot

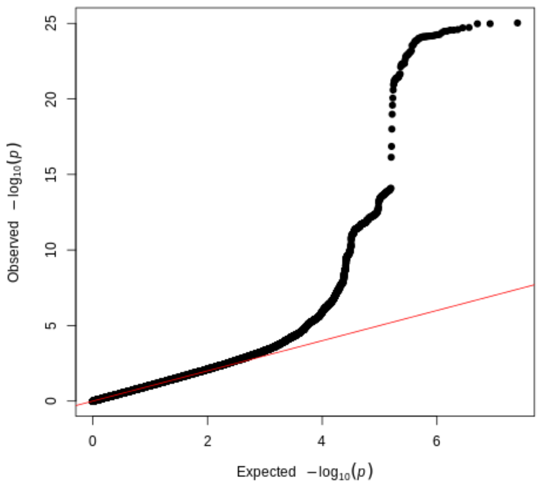

**(d)** Max global quantile-quantile plot

**Figure S 9.** Manhattan and quantile-quantile plots of the genome-wide association analyses of trabeculation traits. The Manhattan plots depict the p-value of association (x-axis, -log10(p-value) and the SNPs across 22 chromosomes (y-axis). The quantile-quantile (QQ) plots depict the expected versus observed p-values for each GWAS. Please see the inflation factor estimates (lambda) in the supplementary tables.

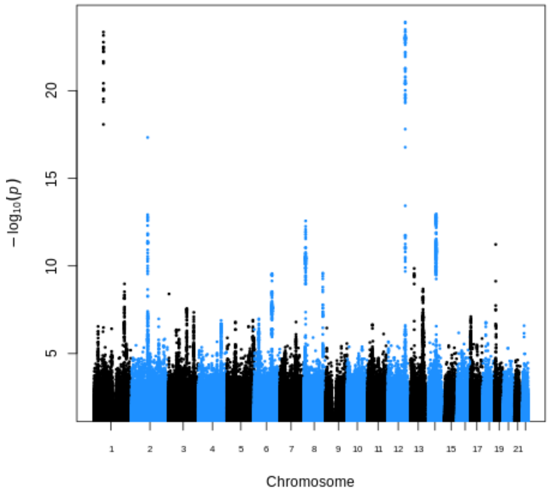

**(a)** Max mid Manhattan plot

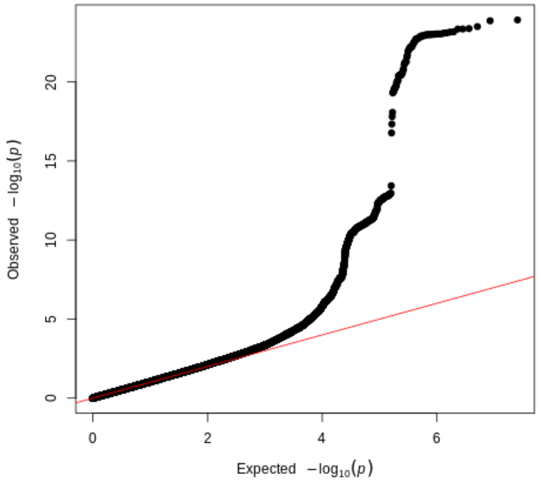

**(b)** Max mid quantile-quantile plot

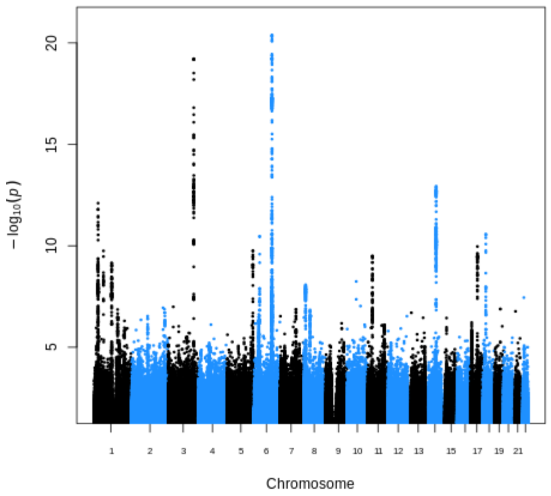

**(c)** Mean apical Manhattan plot

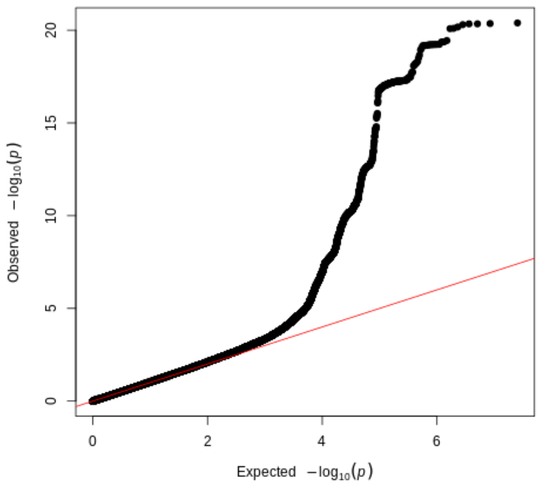

**(d)** Mean apical quantile-quantile plot

**Figure S 10.** Manhattan and quantile-quantile plots of the genome-wide association analyses of trabeculation traits. The Manhattan plots depict the p-value of association (x-axis, -log10(p-value) and the SNPs across 22 chromosomes (y-axis). The quantile-quantile (QQ) plots depict the expected versus observed p-values for each GWAS. Please see the inflation factor estimates (lambda) in the supplementary tables.

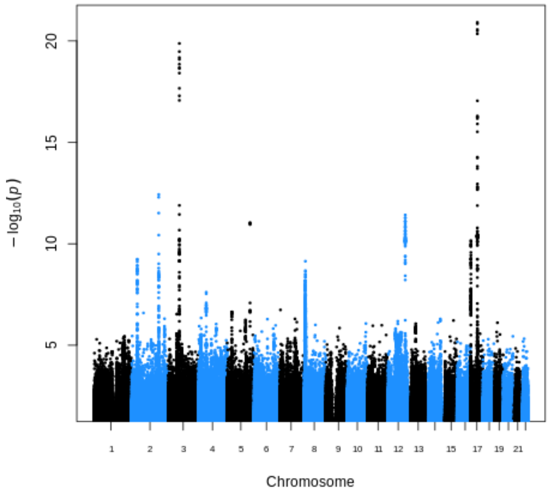

**(a)** Mean basal Manhattan plot

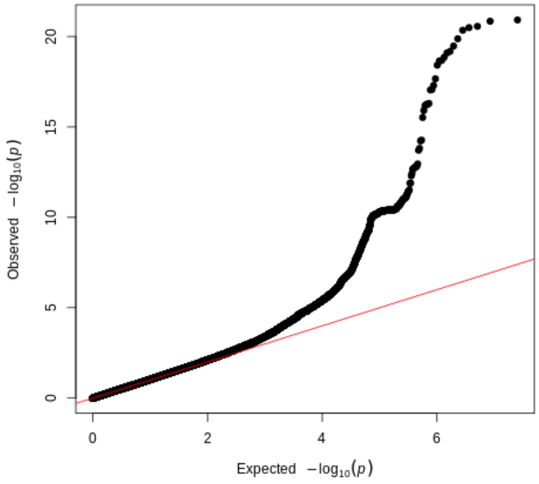

**(b)** Mean basal quantile-quantile plot

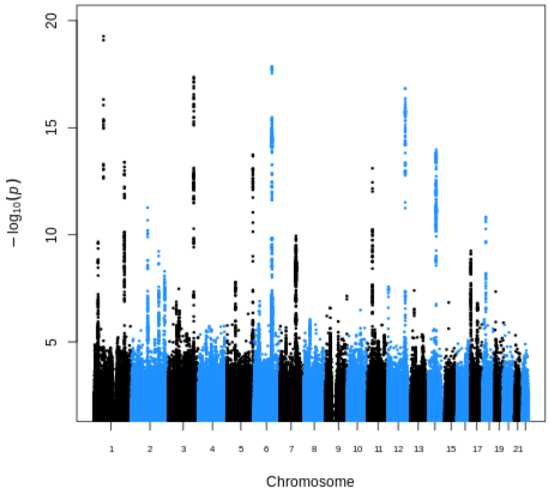

**(c)** Mean global Manhattan plot

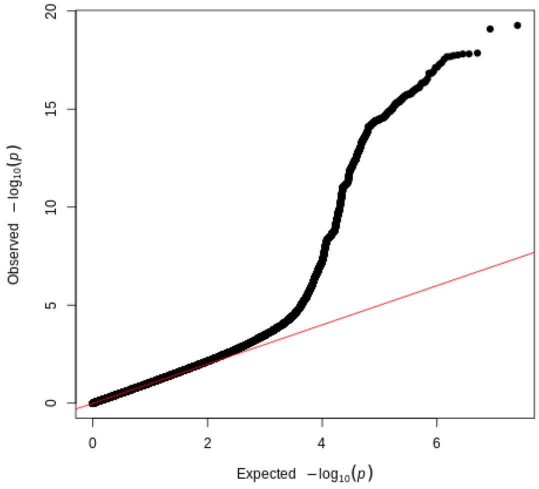

**(d)** Mean global quantile-quantile plot

**Figure S 11.** Manhattan and quantile-quantile plots of the genome-wide association analyses of trabeculation traits. The Manhattan plots depict the p-value of association (x-axis, -log10(p-value) and the SNPs across 22 chromosomes (y-axis). The quantile-quantile (QQ) plots depict the expected versus observed p-values for each GWAS. Please see the inflation factor estimates (lambda) in the supplementary tables.

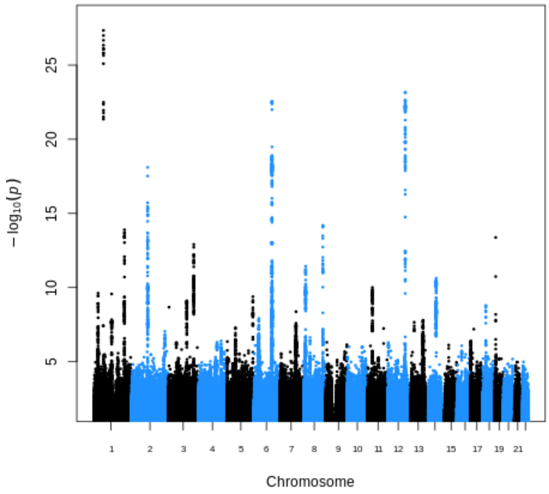

(a) Mean mid Manhattan plot

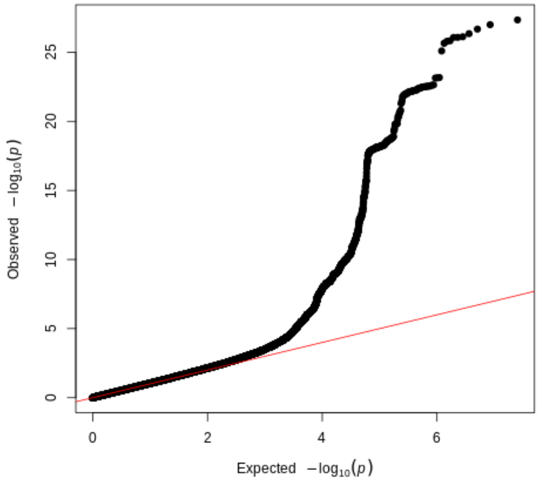

(b) Mean mid quantile-quantile plot

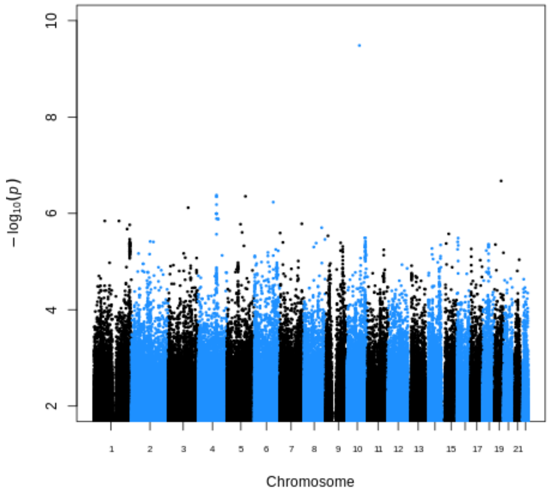

(c) Median apical Manhattan plot

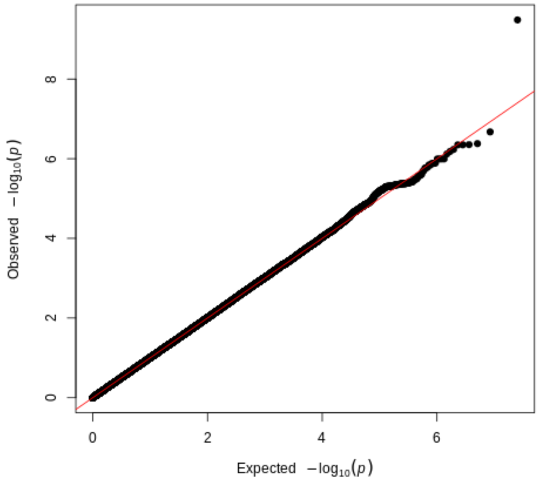

(d) Median apical quantile-quantile plot

**Figure S 12.** Manhattan and quantile-quantile plots of the genome-wide association analyses of trabeculation traits. The Manhattan plots depict the p-value of association (x-axis, -log10(p-value) and the SNPs across 22 chromosomes (y-axis). The quantile-quantile (QQ) plots depict the expected versus observed p-values for each GWAS. Please see the inflation factor estimates (lambda) in the supplementary tables.

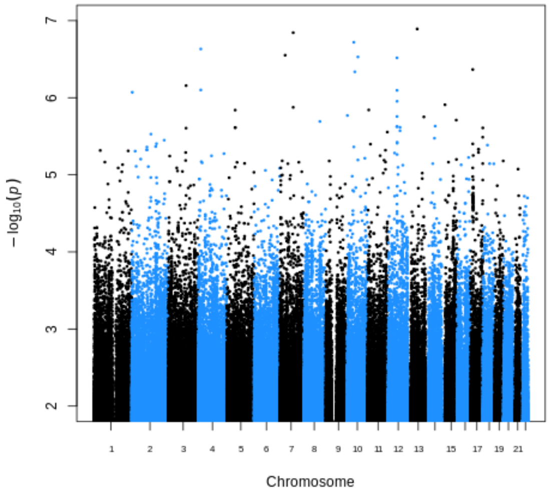

**(a)** Median basal Manhattan plot

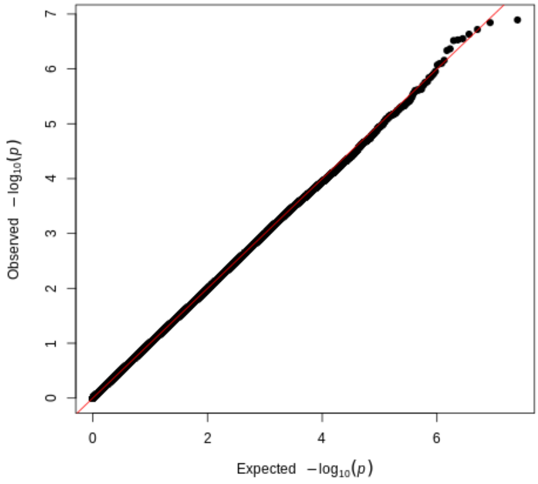

**(b)** Median basal quantile-quantile plot

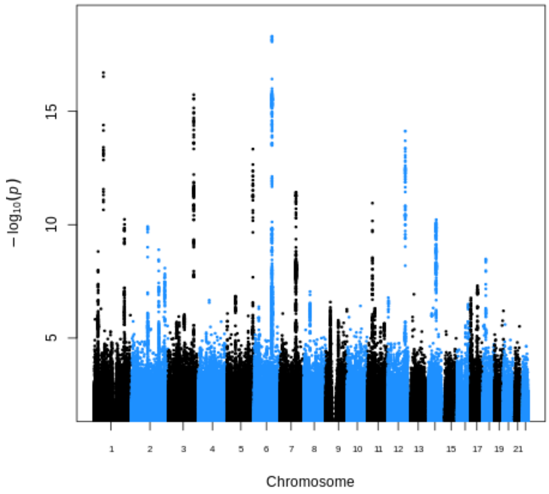

**(c)** Median global Manhattan plot

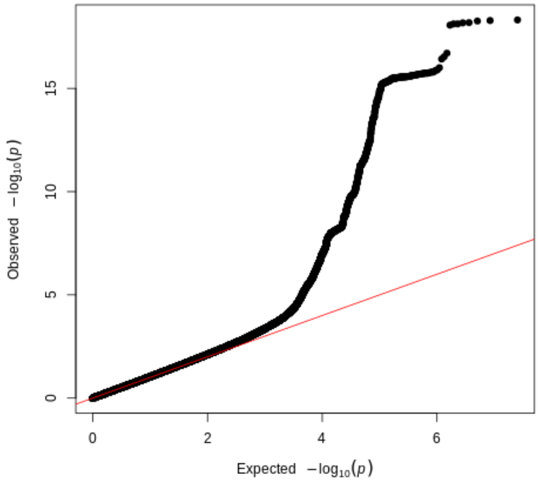

**(d)** Median global quantile-quantile plot

**Figure S 13.** Manhattan and quantile-quantile plots of the genome-wide association analyses of trabeculation traits. The Manhattan plots depict the p-value of association (x-axis, -log10(p-value) and the SNPs across 22 chromosomes (y-axis). The quantile-quantile (QQ) plots depict the expected versus observed p-values for each GWAS. Please see the inflation factor estimates (lambda) in the supplementary tables.

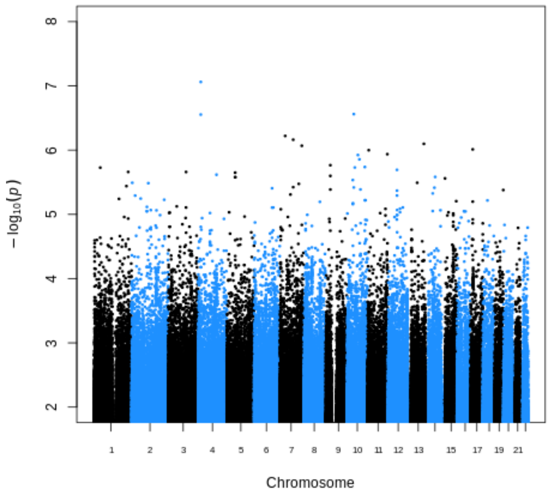

**(a)** Median mid Manhattan plot

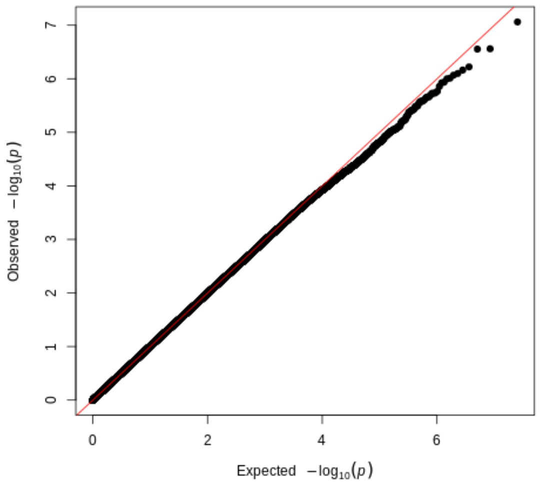

**(b)** Median mid quantile-quantile plot

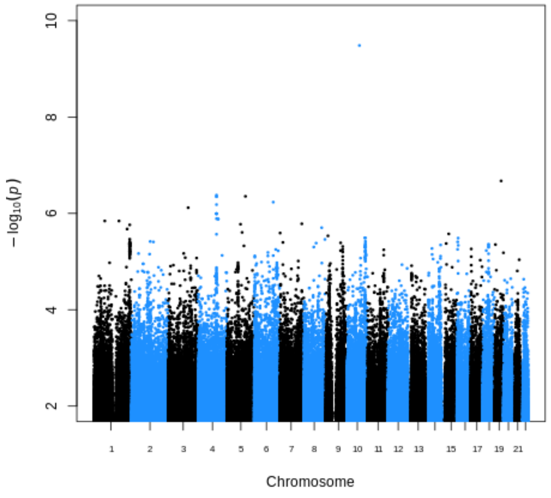

**(c)** Minimum apical Manhattan plot

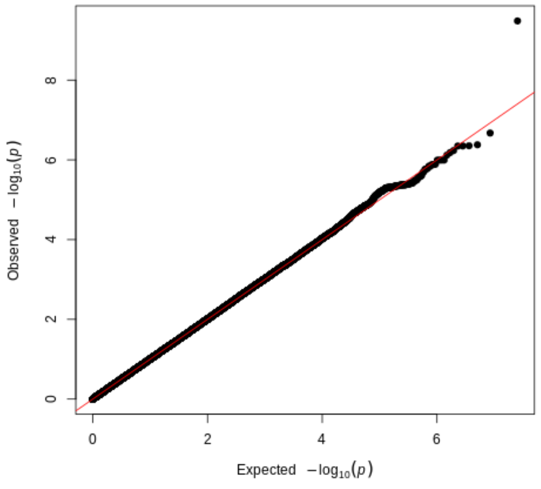

**(d)** Minimum apical quantile-quantile plot

**Figure S 14.** Manhattan and quantile-quantile plots of the genome-wide association analyses of trabeculation traits. The Manhattan plots depict the p-value of association (x-axis, -log10(p-value) and the SNPs across 22 chromosomes (y-axis). The quantile-quantile (QQ) plots depict the expected versus observed p-values for each GWAS. Please see the inflation factor estimates (lambda) in the supplementary tables.

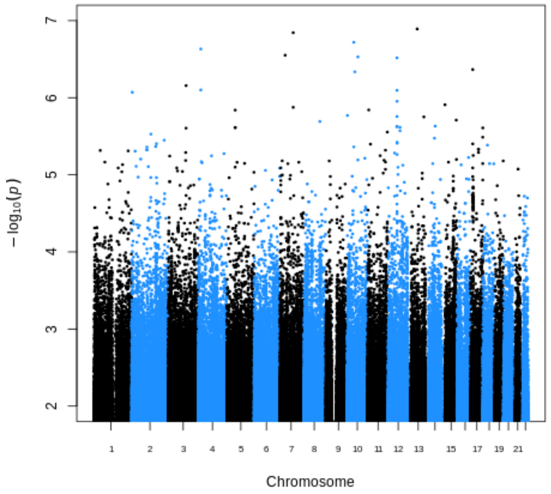

**(a)** Minimum basal Manhattan plot

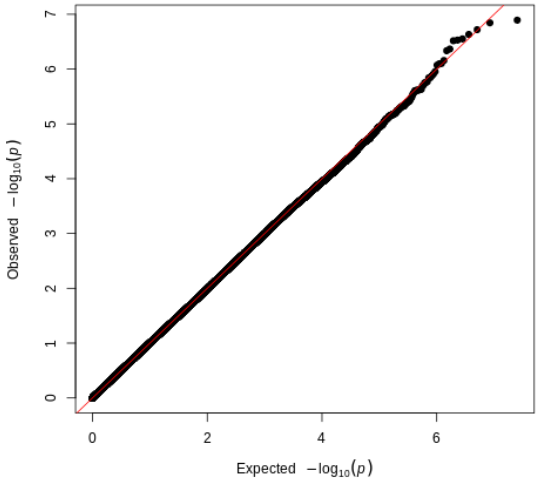

**(b)** Minimum basal quantile-quantile plot

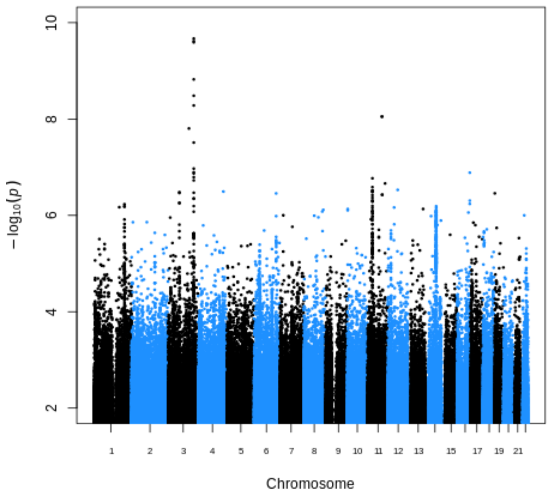

**(c)** Minimum global Manhattan plot

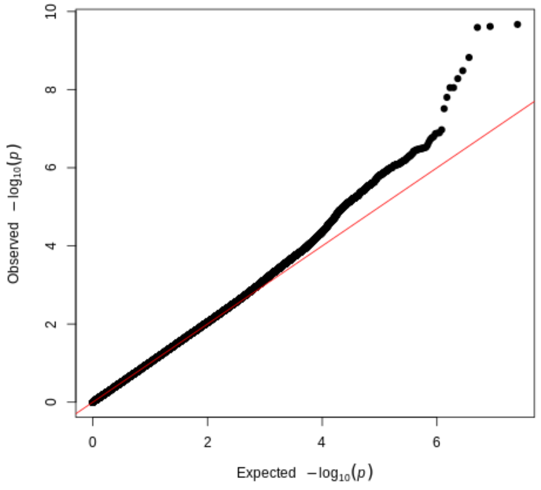

**(d)** Minimum global quantile-quantile plot

**Figure S 15.** Manhattan and quantile-quantile plots of the genome-wide association analyses of trabeculation traits. The Manhattan plots depict the p-value of association (x-axis, -log10(p-value) and the SNPs across 22 chromosomes (y-axis). The quantile-quantile (QQ) plots depict the expected versus observed p-values for each GWAS. Please see the inflation factor estimates (lambda) in the supplementary tables.

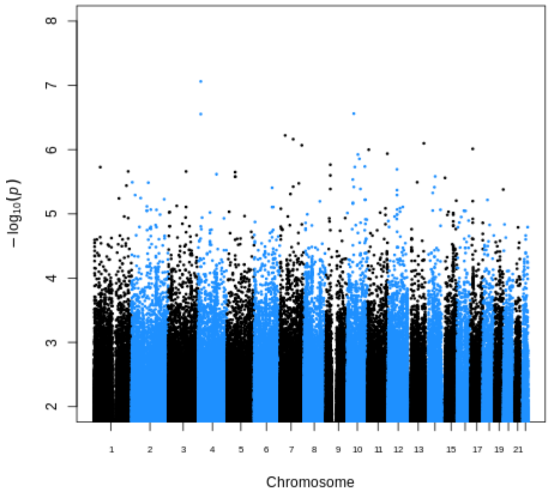

(a) Minimum mid Manhattan plot

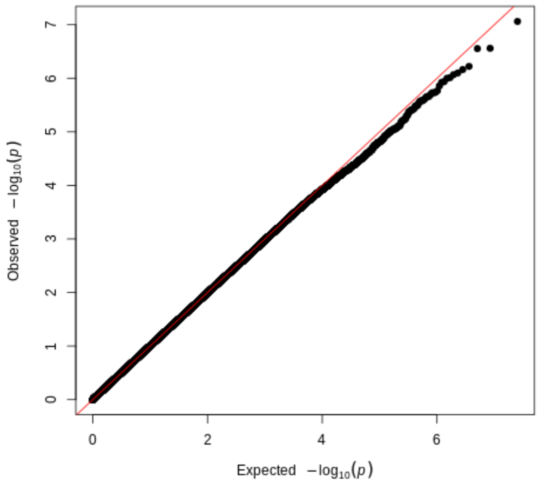

(b) Minimum mid quantile-quantile plot

**Figure S 16.** Manhattan and quantile-quantile plots of the genome-wide association analyses of trabeculation traits. The Manhattan plots depict the p-value of association (x-axis, -log10(p-value) and the SNPs across 22 chromosomes (y-axis). The quantile-quantile (QQ) plots depict the expected versus observed p-values for each GWAS. Please see the inflation factor estimates (lambda) in the supplementary tables.

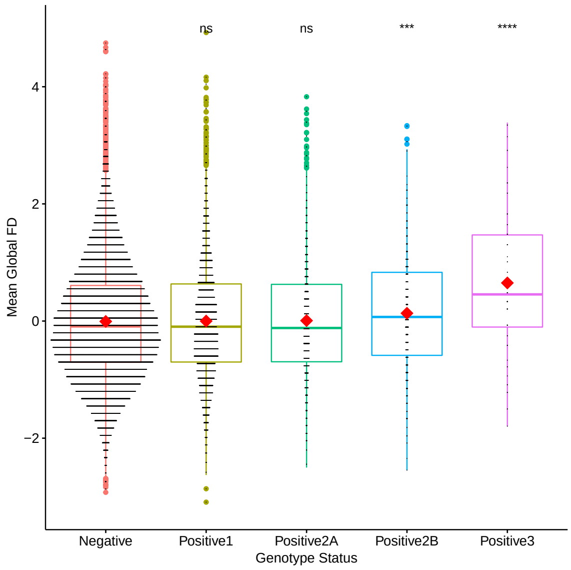

**(a)** Increase in mean global trabeculation with HCM variant pathogenicity.

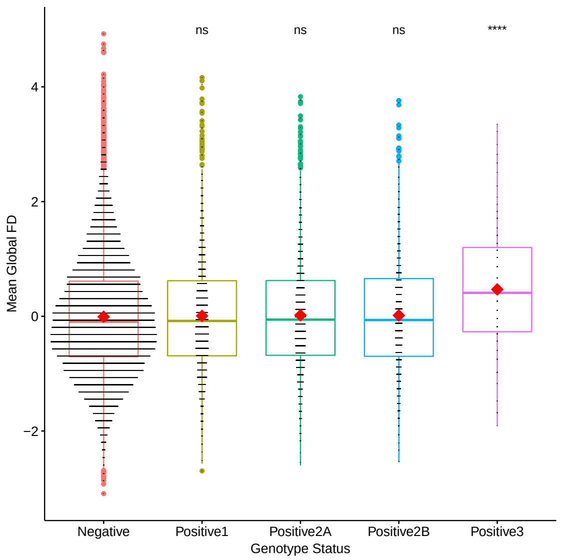

**(b)** Increase in mean global trabeculation with DCM variant pathogenicity.

**Figure S 17.** Increase in mean global trabeculation with variant pathogenicity. These independent groups of individuals for **A)** HCM and **B)** DCM, were separated by carrier status. For HCM-associated variants, there were 28,006 genotype-negative participants (Negative), 6,040 participants with variants in syndromic genes (Positive1), 1,330 participants with common variants (AF<0.01 & >0.00004) in definitive-evidence genes (Positive2A), 680 participants with rare indeterminant (VUS) variants in definitive-evidence genes (Positive2B - significant), 72 carriers of P/LP variants (Positive3 - significant). For DCM-associated variants, there were 28,109 genotype-negative participants (Negative), 3,379 participants with variants in syndromic genes (Positive1), 2,710 participants with common variants (AF<0.01 & >0.000084) in definitive-evidence genes (Positive2A), 1,837 participants with rare indeterminant variants in definitive-evidence genes (Positive2B), 162 carriers of P/LP variants (Positive 3 - significant). Means were compared against the genotype-negative group.

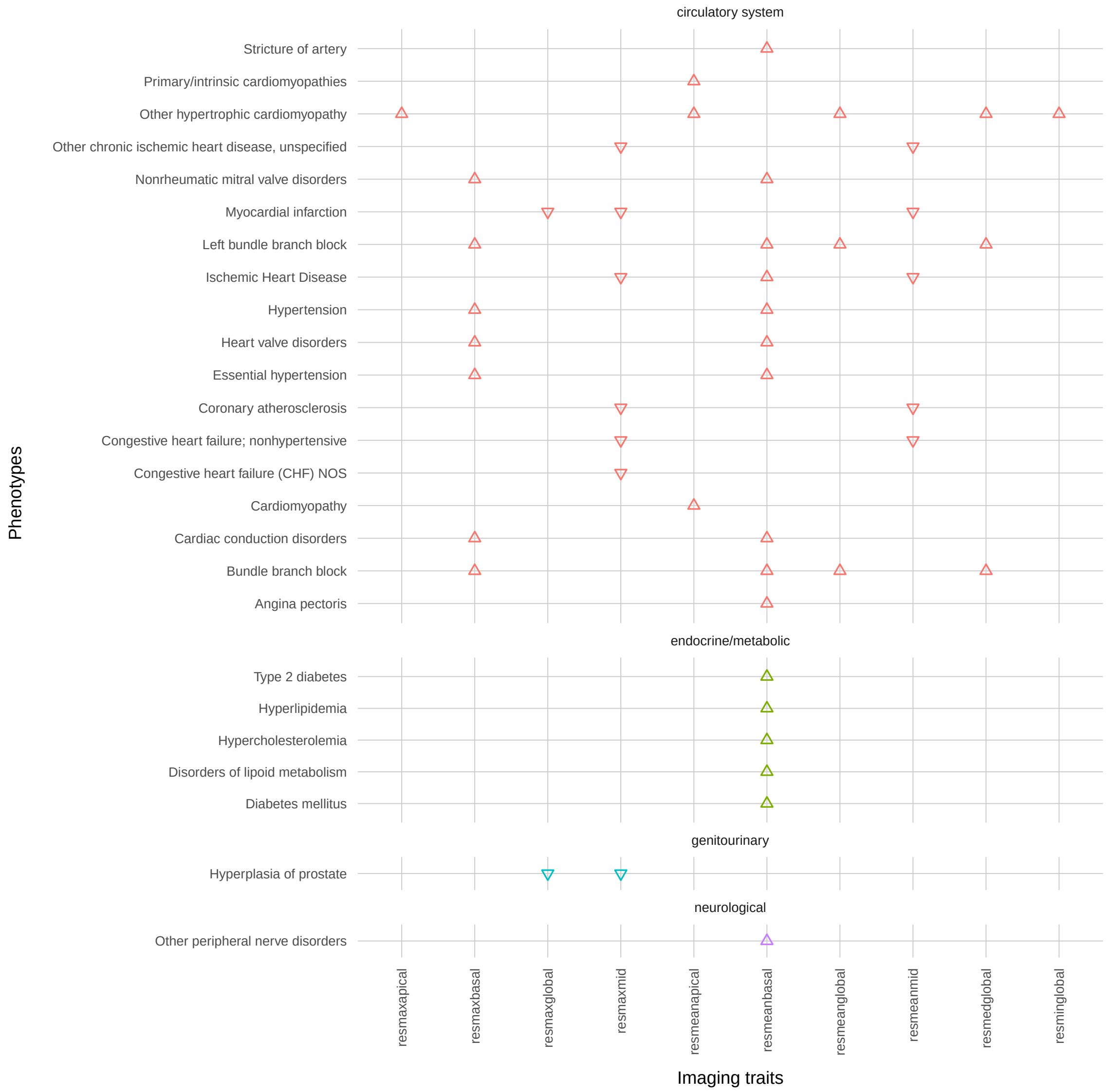

**Figure S 18.** PheWAS for summary trabeculation measures when adjusted for LVEDV.

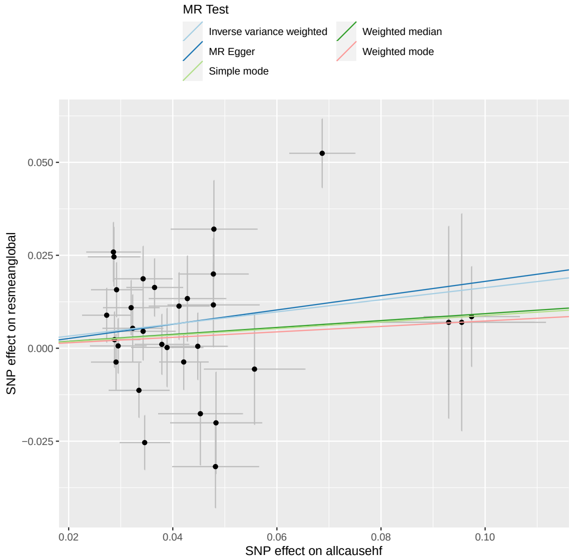

**(a)** Mendelian randomization scatter plot for HF as exposure on mean global FD.

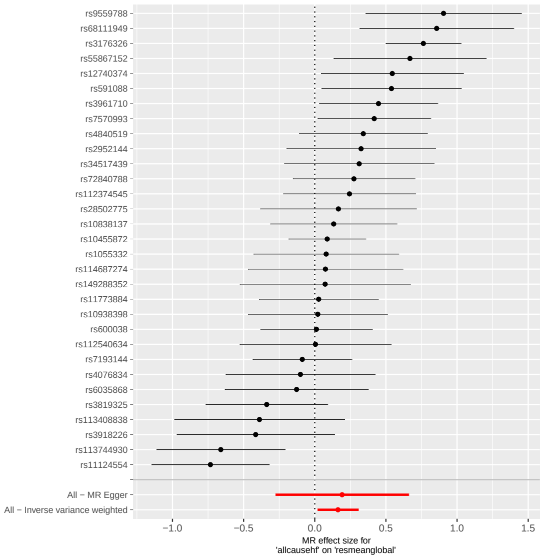

**(b)** Mendelian randomization single SNP funnel plot for HF as exposure on mean global FD.

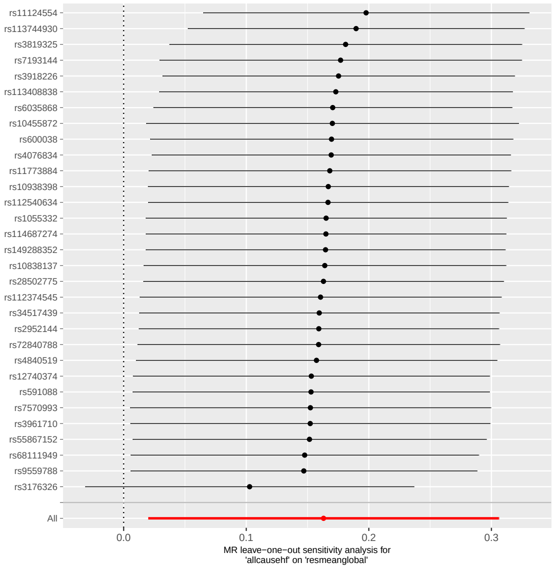

**(c)** Mendelian randomization single SNP forest plot for HF as exposure on mean global FD.

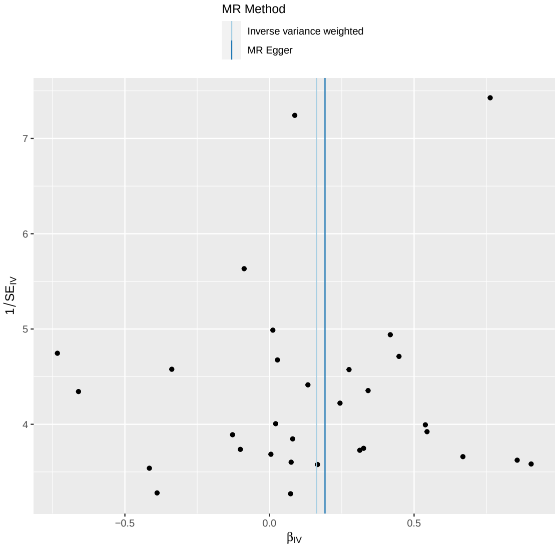

**(d)** Mendelian randomization leave one out plot for HF as exposure on mean global FD.

**Figure S 19.** Mendelian randomization analysis of HF as exposure on mean global FD. The plots show summary information on the analyses, performed as per the TwoSampleMR R package.

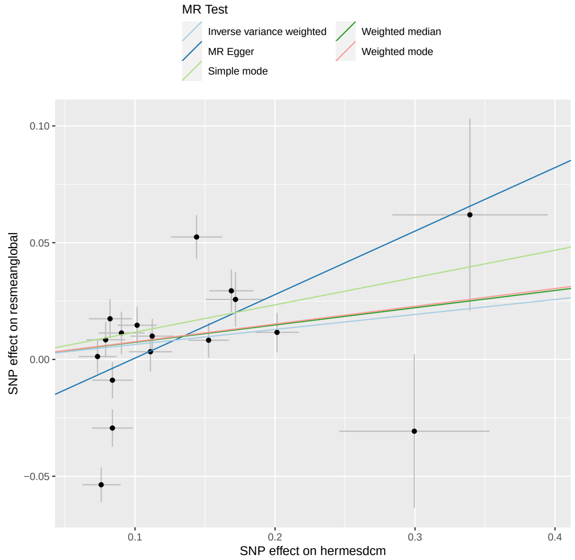

**(a)** Mendelian randomization scatter plot for DCM as exposure on trabeculation.

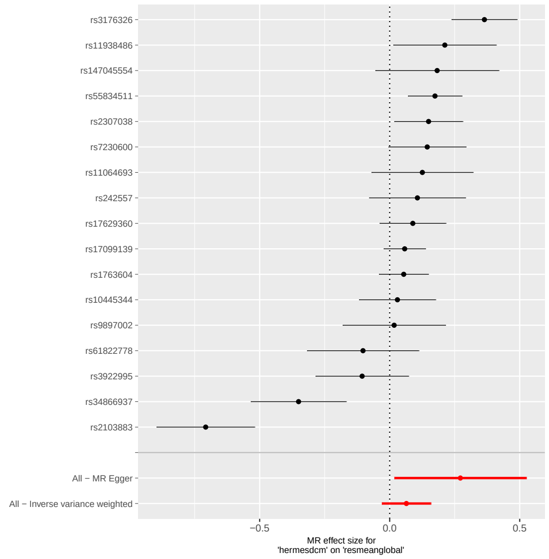

**(b)** Mendelian randomization single SNP funnel plot for DCM as exposure on trabeculation.

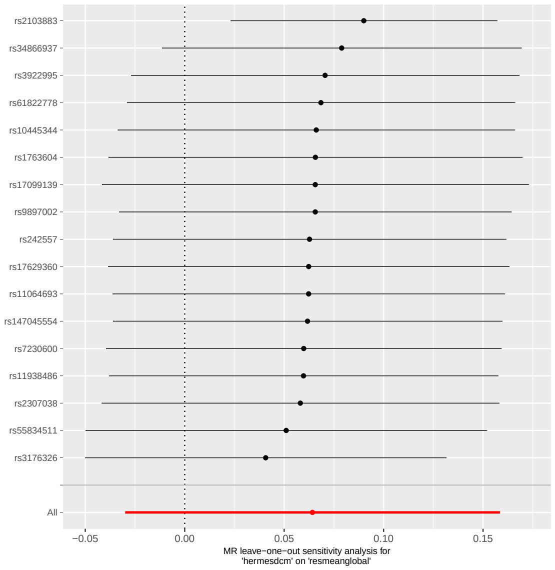

**(c)** Mendelian randomization single SNP forest plot for DCM as exposure on trabeculation.

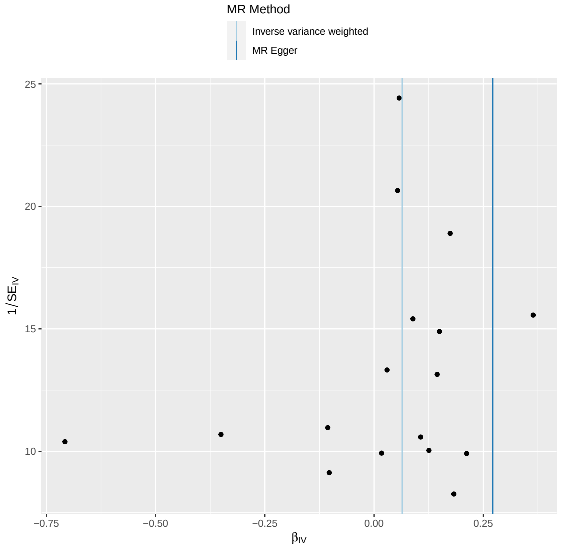

**(d)** Mendelian randomization leave one out plot for DCM as exposure on trabeculation.

**Figure S 20.** Mendelian randomization analysis of DCM as exposure on trabeculation outcome. The plots show summary information on the analyses, performed as per the TwoSampleMR R package.

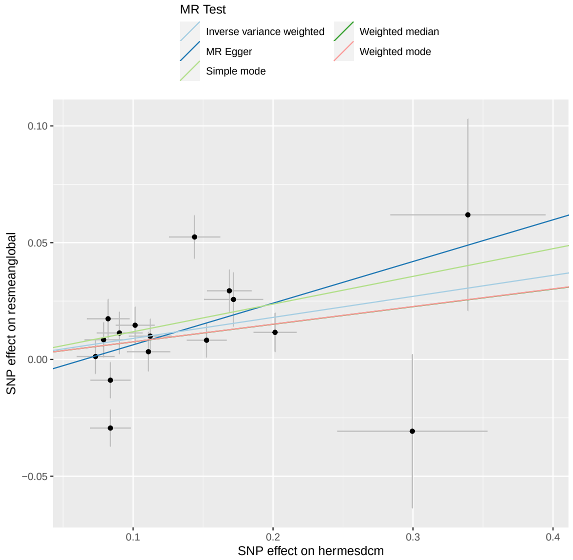

**(a)** Mendelian randomization scatter plot for DCM as exposure on trabeculation.

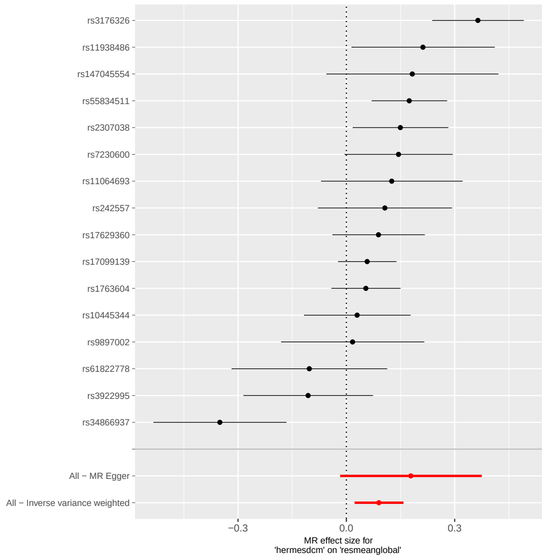

**(b)** Mendelian randomization single SNP funnel plot for DCM as exposure on trabeculation.

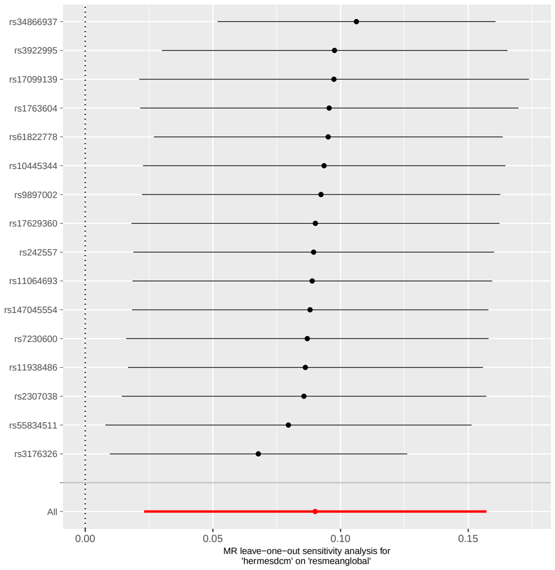

**(c)** Mendelian randomization single SNP forest plot for DCM as exposure on trabeculation.

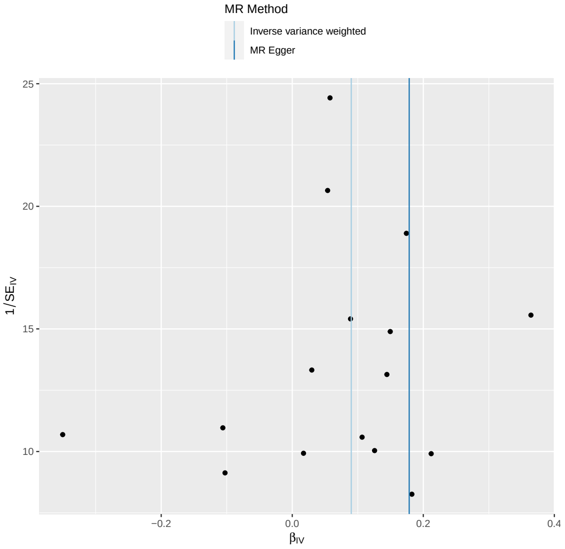

**(d)** Mendelian randomization leave one out plot for DCM as exposure on trabeculation.

**Figure S 21.** Mendelian randomization analysis of DCM as exposure on trabeculation outcome, pruning for an NFIA variant. The plots show summary information on the analyses, performed as per the TwoSampleMR R package.

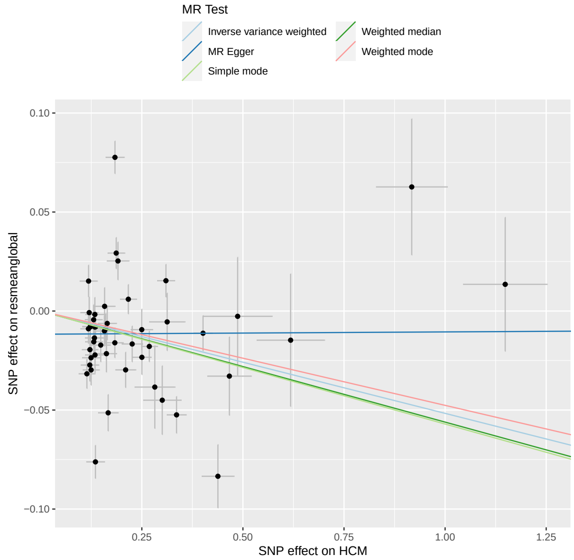

**(a)** Mendelian randomization scatter plot for HCM as exposure on trabeculation.

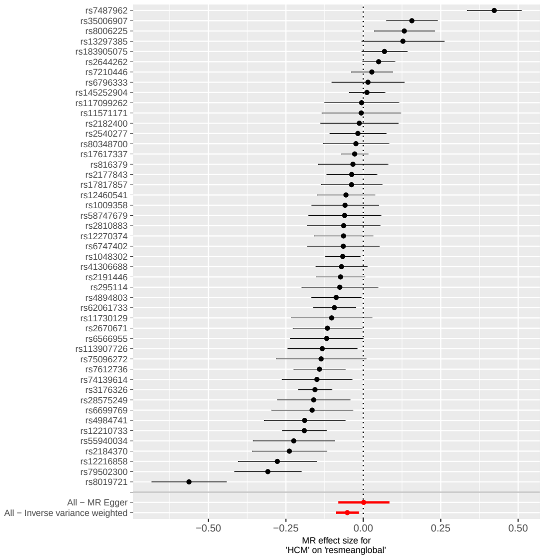

**(b)** Mendelian randomization single SNP funnel plot for HCM as exposure on trabeculation.

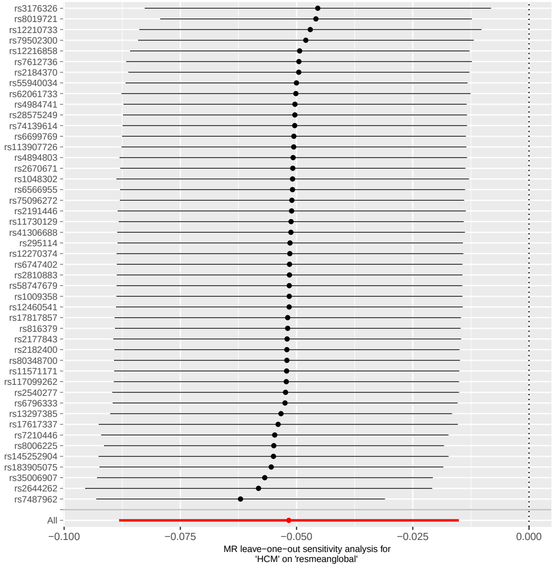

**(c)** Mendelian randomization single SNP forest plot for HCM as exposure on trabeculation.

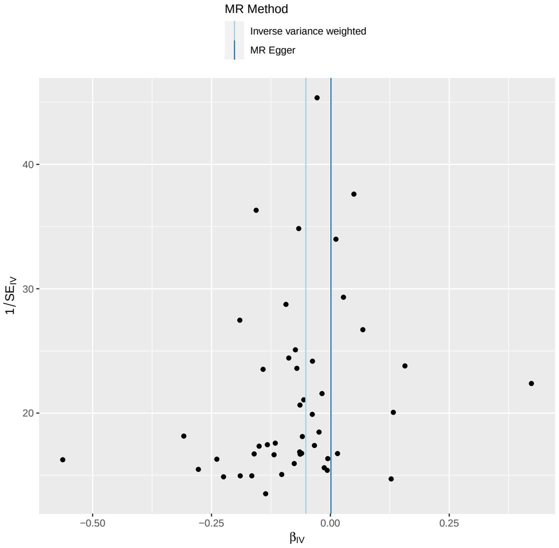

**(d)** Mendelian randomization leave one out plot for HCM as exposure on trabeculation.

**Figure S 22.** Mendelian randomization analysis of for HCM as exposure on trabeculation outcome. The plots show summary information on the analyses, performed as per the TwoSampleMR R package.

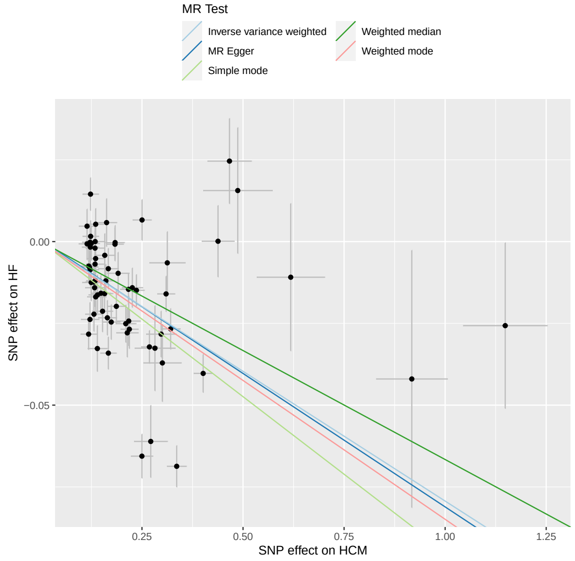

**(a)** Mendelian randomization scatter plot for HCM as exposure on HF.

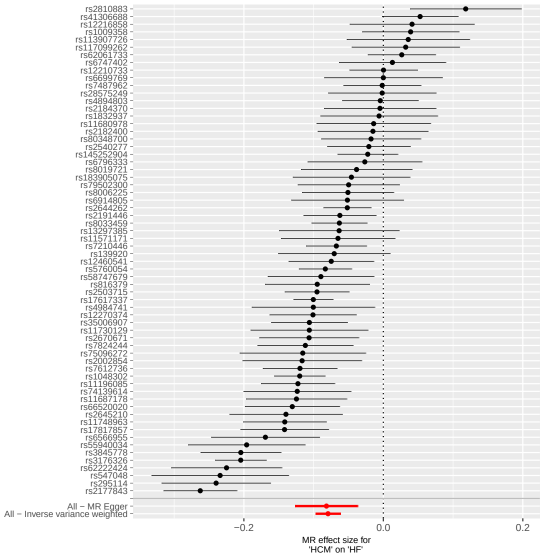

**(b)** Mendelian randomization single SNP funnel plot for HCM as exposure on HF.

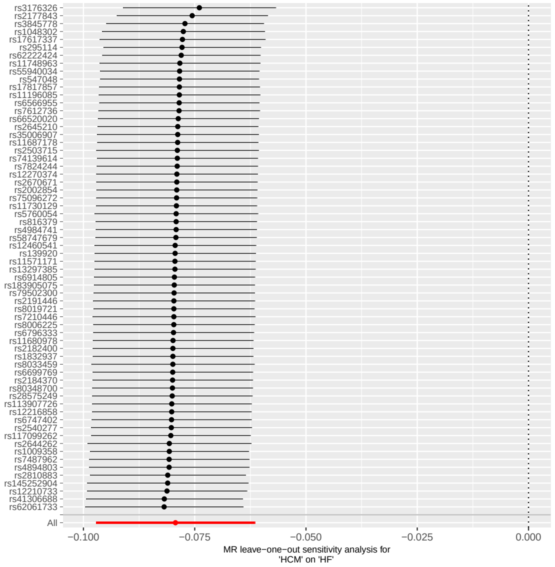

**(c)** Mendelian randomization single SNP forest plot for HCM as exposure on HF.

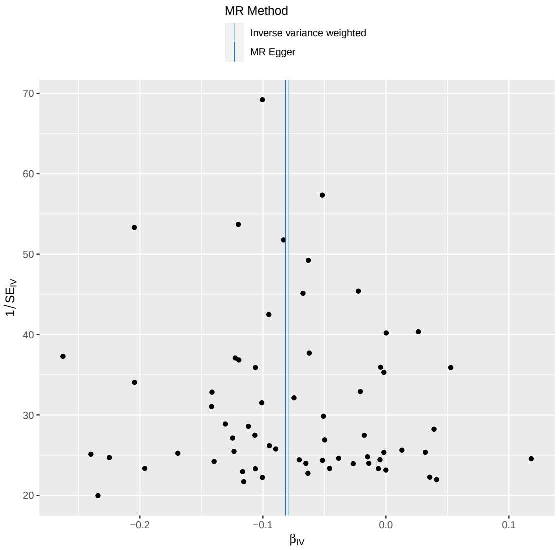

**(d)** Mendelian randomization leave one out plot for HCM as exposure on HF.

**Figure S 23.** Mendelian randomization analysis of for HCM as exposure on HF outcome. The plots show summary information on the analyses, performed as per the TwoSampleMR R package.

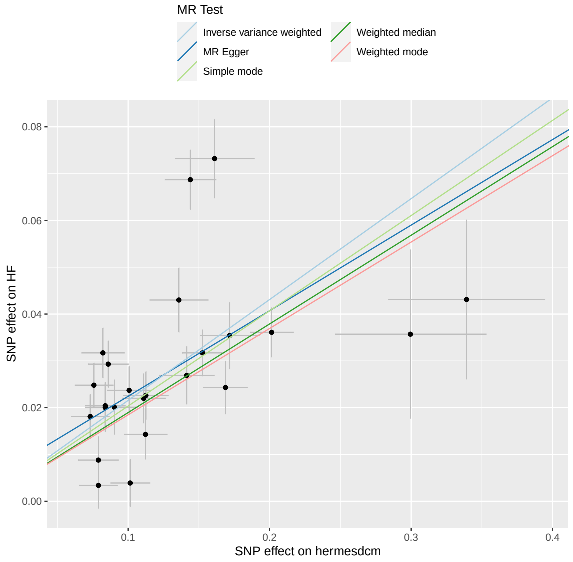

**(a)** Mendelian randomization scatter plot for DCM as exposure on HF.

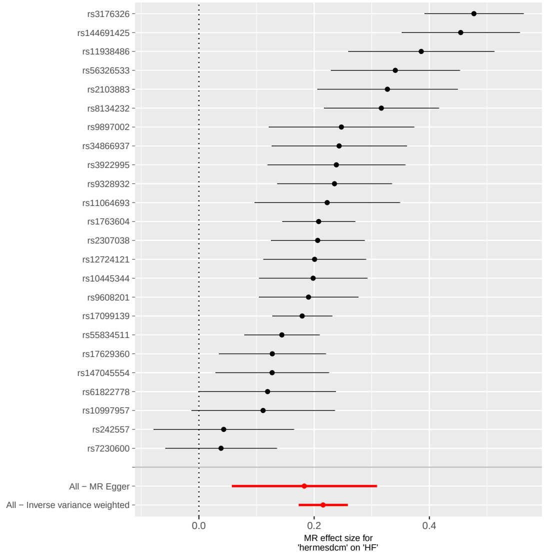

**(b)** Mendelian randomization single SNP funnel plot for DCM as exposure on HF.

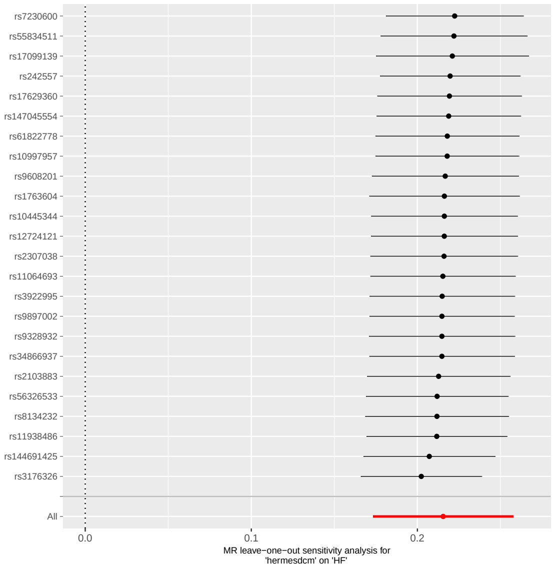

**(c)** Mendelian randomization single SNP forest plot for DCM as exposure on HF.

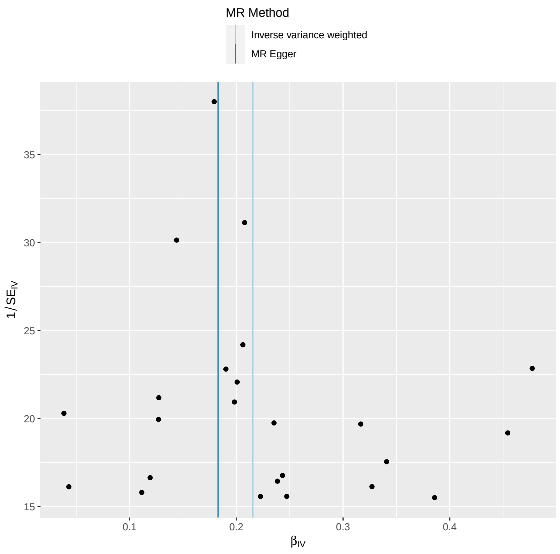

**(d)** Mendelian randomization leave one out plot for DCM as exposure on HF.

**Figure S 24.** Mendelian randomization analysis of DCM as exposure on HF outcome. The plots show summary information on the analyses, performed as per the TwoSampleMR R package.

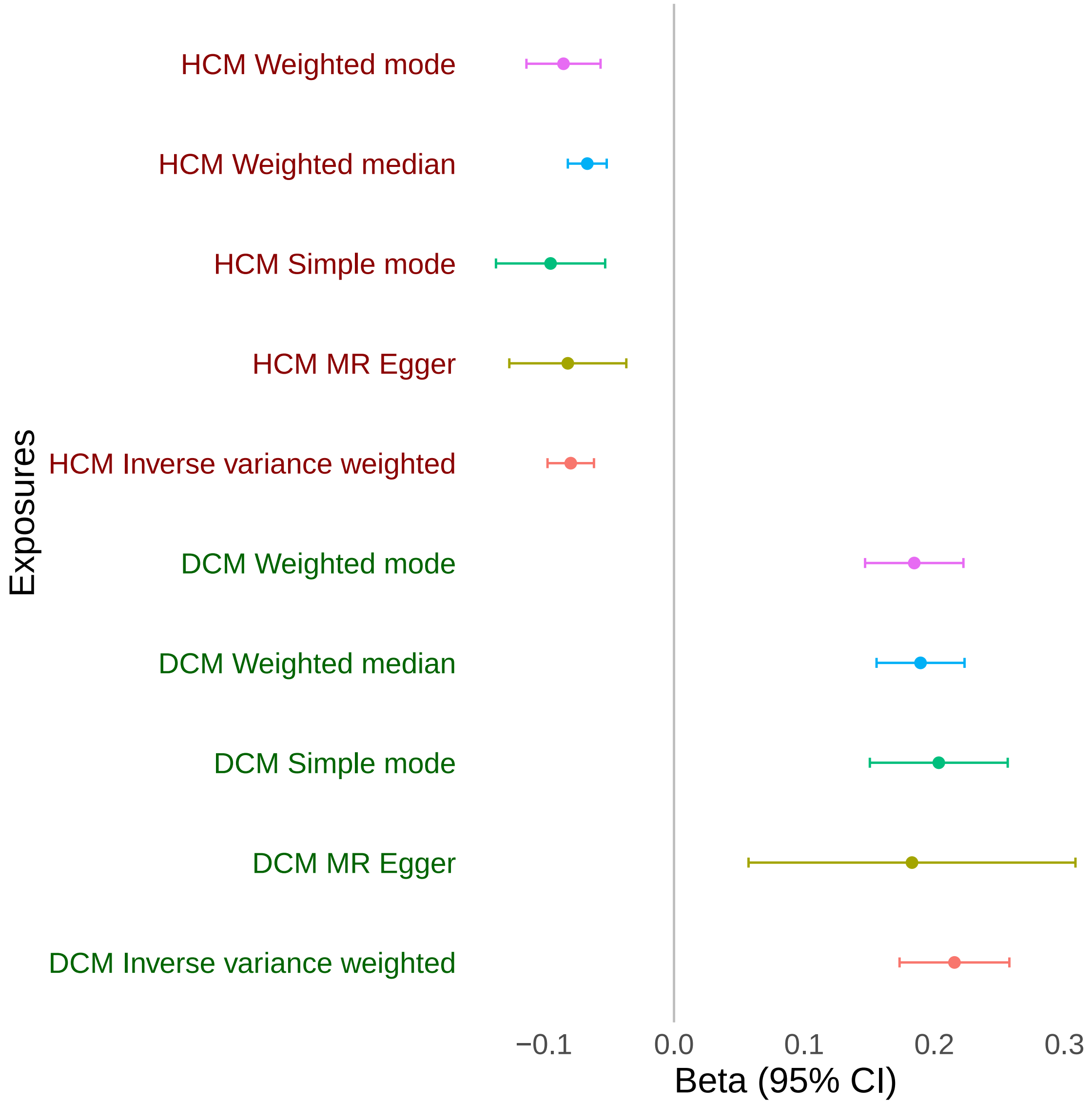

**Figure S 25.** Mendelian randomization analysis of DCM and HCM as exposures on HF outcome.

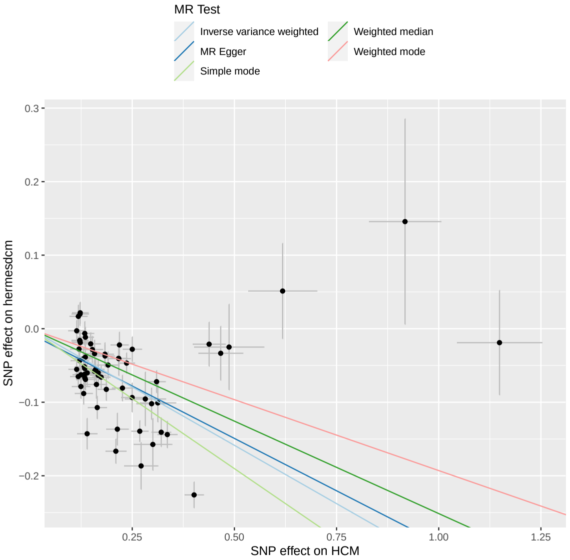

**(a)** Mendelian randomization scatter plot for HCM as exposure on DCM.

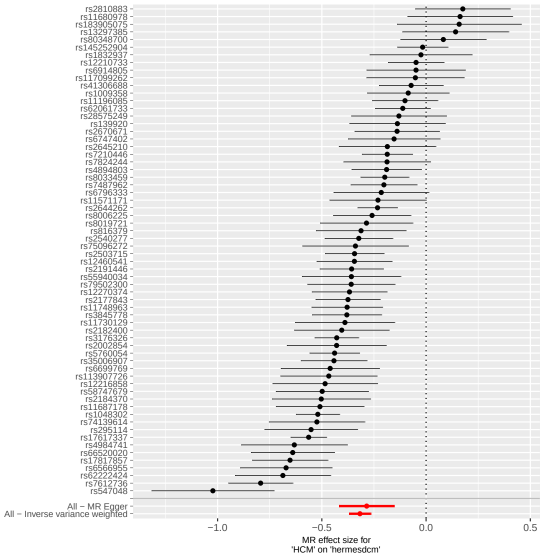

**(b)** Mendelian randomization single SNP funnel plot for HCM as exposure on DCM.

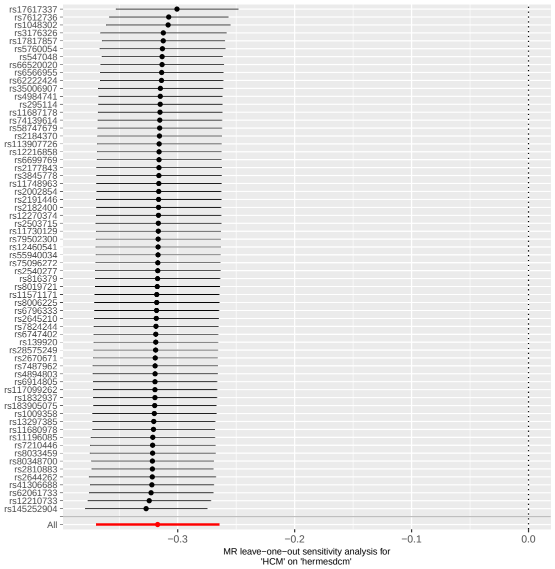

**(c)** Mendelian randomization single SNP forest plot for HCM as exposure on DCM.

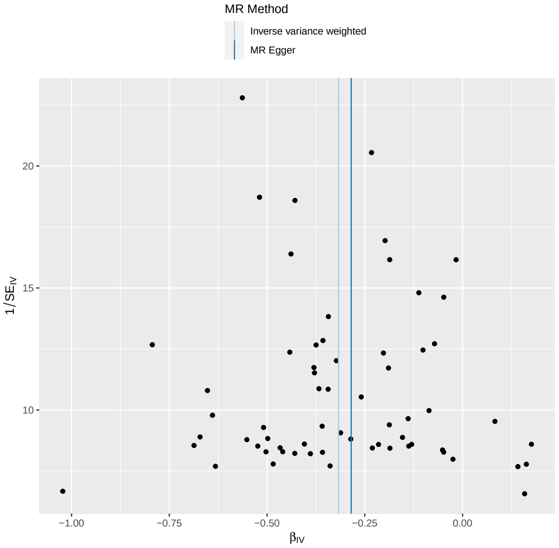

**(d)** Mendelian randomization leave one out plot for HCM as exposure on DCM.

**Figure S 26.** Mendelian randomization analysis of HCM as exposure on DCM outcome. The plots show summary information on the analyses, performed as per the TwoSampleMR R package.

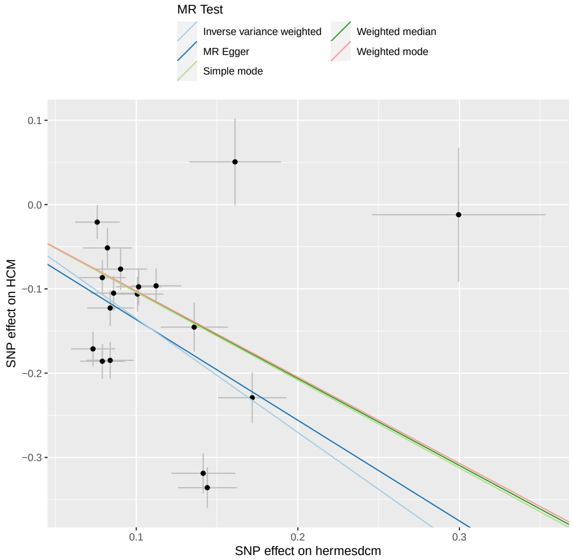

**(a)** Mendelian randomization scatter plot for DCM as exposure on HCM.

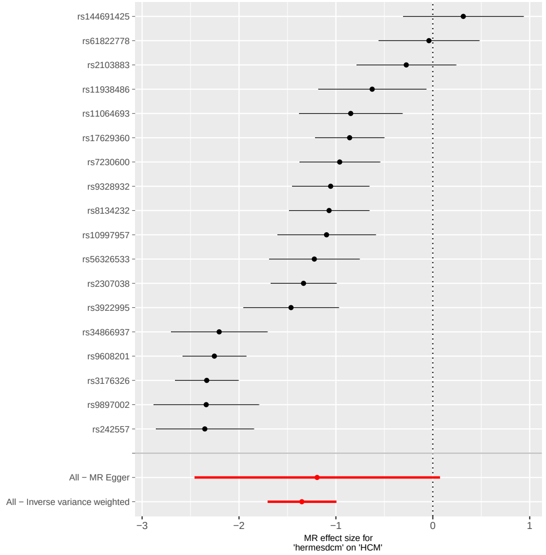

**(b)** Mendelian randomization single SNP funnel plot for DCM as exposure on HCM.

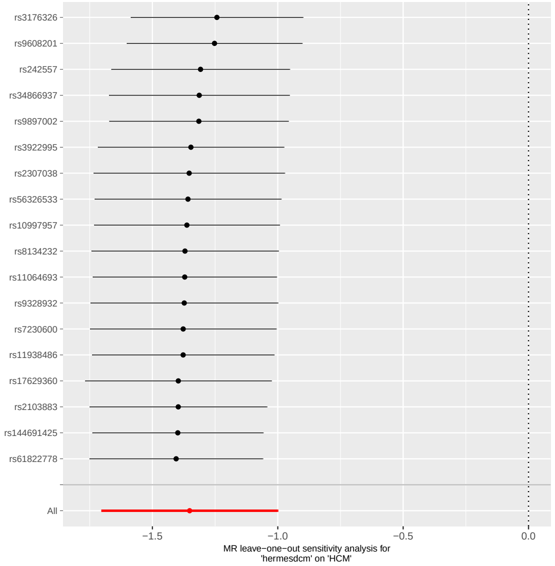

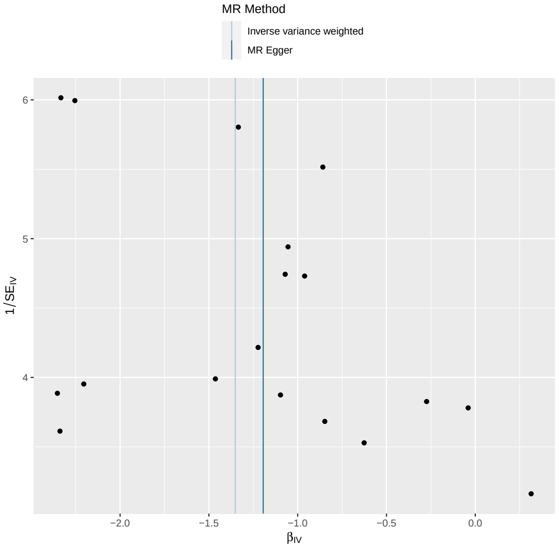

**(c)** Mendelian randomization single SNP forest plot for DCM as exposure on HCM. **(d)** Mendelian randomization leave one out plot for DCM as exposure on HCM.

**Figure S 27.** Mendelian randomization analysis of DCM as exposure on HCM outcome. The plots show summary information on the analyses, performed as per the TwoSampleMR R package.

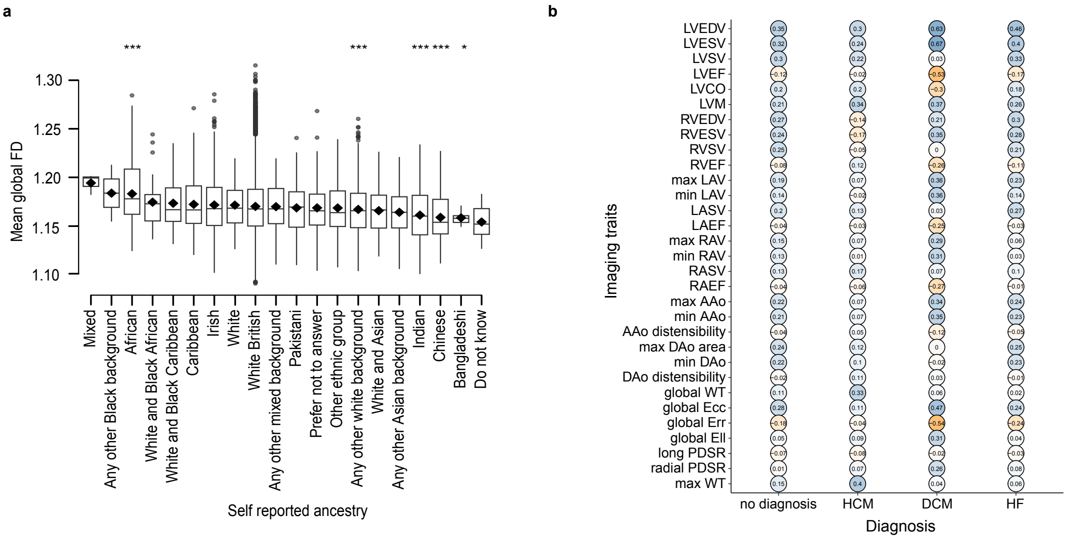

**Figure S 28.** Association of trabecular morphology with ancestry and cardiac MRI-derived summary measures. The analyses were completed on 38,245 participants of the UK Biobank population. a) Compared to British ancestry which dominates the UK Biobank, the mean global fractal dimension was increased for participants of African ancestry. Indian, Chinese, Bangladeshi, and "Any other white background" self-reported ancestry had the lowest significant FD. Student's two-sided t-test was used to compare means. b) The table quantifies the relationship (correlation coefficient, R) between mean global fractal dimension and 2D summary imaging cardiac measures separately for participants with no diagnosis (n=31,067), hypertrophic cardiomyopathy (HCM, n=31), dilated cardiomyopathy (DCM, n=29), and heart failure (n=332).

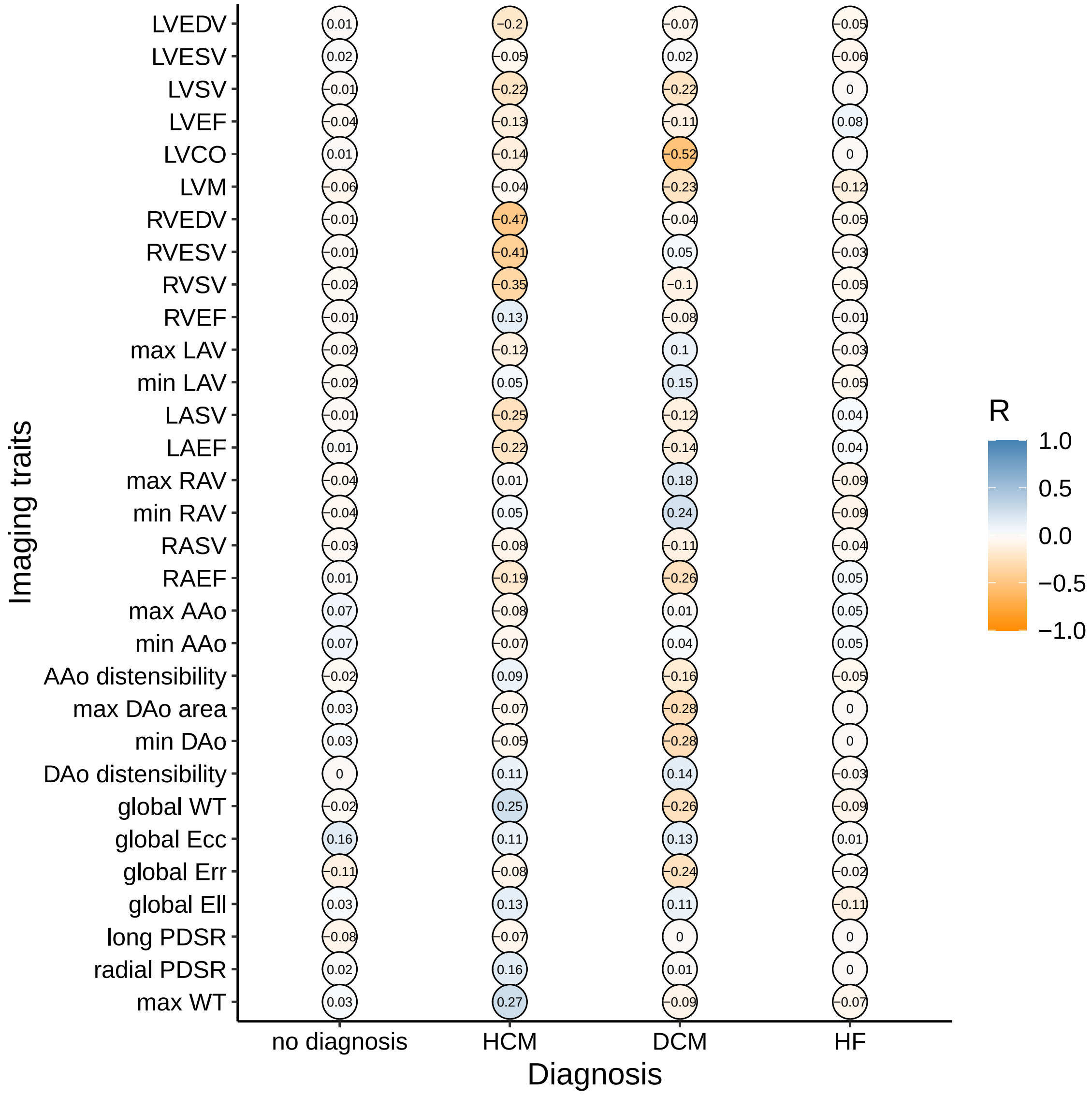

**Figure S 29.** The relationship of mean global FD adjusted for LVEDV with summary CMR measures in participants with cardiomyopathy or heart failure.

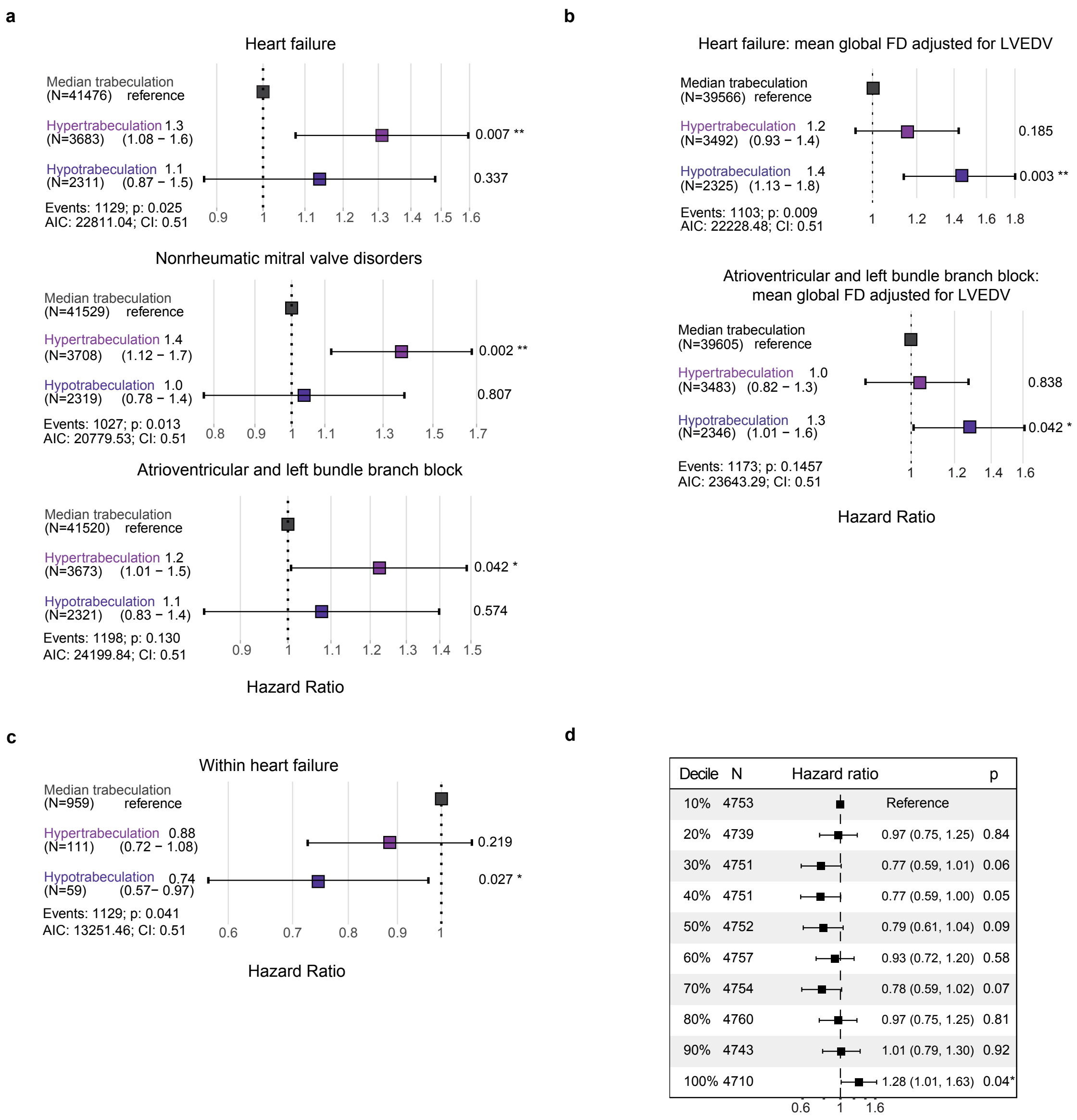

**Figure S 30.** Outcome associations. The analyses were completed on 47,803 participants of the UK Biobank population and assessed from years since imaging for a diagnosis of a cardiac condition or all-cause death, with anyone diagnosed before imaging excluded. Trabeculation was separated into three groups: hyper-, median, and hypotrabeculation where <1.5 SD is hypotrabeculation and >1.5 SD is hypertrabeculation. a) The hazard ratio for heart failure, for mitral valve disorders, and bundle branch block, showed increased risk with hypertrabeculation. b) The hazard ratio with the adjustment for LVEDV (and removal of the association of trabeculation with DCM) showed that hypotrabeculation was associated with the risk of non-DCM heart failure. c) The hazard ratio for assessments with participants diagnosed with heart failure only showed that heart failure risk increased with hypotrabeculation. d) The mean coxfit linear predictors for heart failure were plotted for trabeculation by decile. Of those with heart failure or death, participants with hypotrabeculation were diagnosed or died nearly a year later on average. Confidence intervals and log-rank p-values are depicted. CI, concordance index.

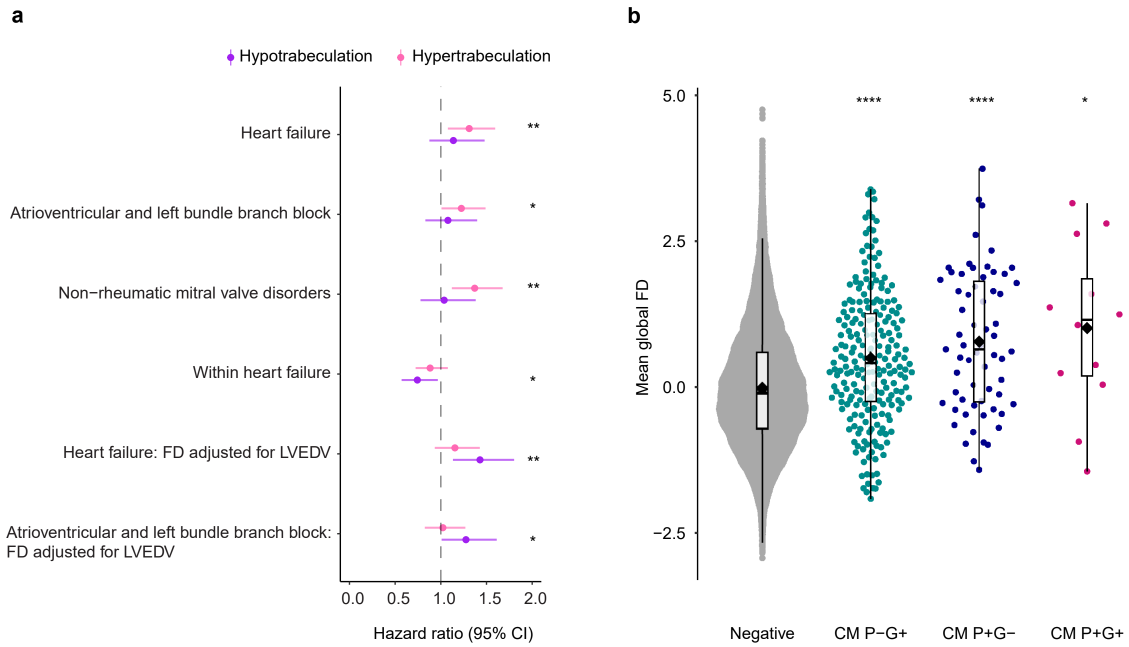

**Figure S 31.** Selected outcome associations. **a**, The analyses were completed on 47,803 participants of the UK Biobank population and assessed from years since imaging for a diagnosis of a cardiac condition or all-cause death, with anyone diagnosed before imaging excluded. Trabeculation was separated into three groups: hyper-, median, and hypotrabeculation where <-1.5 SD is hypotrabeculation and >1.5 SD is hypertrabeculation, where hypotrabeculation and hypertrabeculation were compared to the median trabeculation (dashed line). The hazard ratio for heart failure, mitral valve disorders, and bundle branch block, showed increased risk with hypertrabeculation. All cause death was also assessed but was not significant. The hazard ratio with the adjustment for LVEDV (and removal of the association of trabeculation with DCM) showed that hypotrabeculation was associated with risk of non-DCM heart failure. The hazard ratio for assessments with participants diagnosed with heart failure only showed that heart failure risk increased with hypotrabeculation. Point estimates with 95% confidence intervals (CI) are shown. *, P<0.05; **, P<0.01; ***, P<0.001; ****, P<0.0001. **b**, Carriers of cardiomyopathy-associated pathogenic or likely pathogenic variants (G+) with or without a diagnosis of disease (phenotype; P+/-) had increased mean trabeculation compared to genotype negatives. There were 23,936 genotype-negative participants, 222 participants with a P/LP variant without a diagnosis of cardiomyopathy, 63 participants with disease and no variant in definitive-evidence genes, and 12 participants with a variant and diagnosis of disease. Student's two-sided t-test was used to compare the group means. The lower and upper hinges in the box plot correspond to the 25th and 75th percentiles (interquartile range (IQR)), respectively. The horizontal line in the box plot indicates the median. The lower and upper whiskers extend from the hinge to the smallest and largest values no further than 1.5× the IQR, respectively. Each dot is one individual. P-G+, phenotype negative, genotype positive; P+G-, phenotype positive, genotype negative; P+G+, phenotype positive, genotype positive.

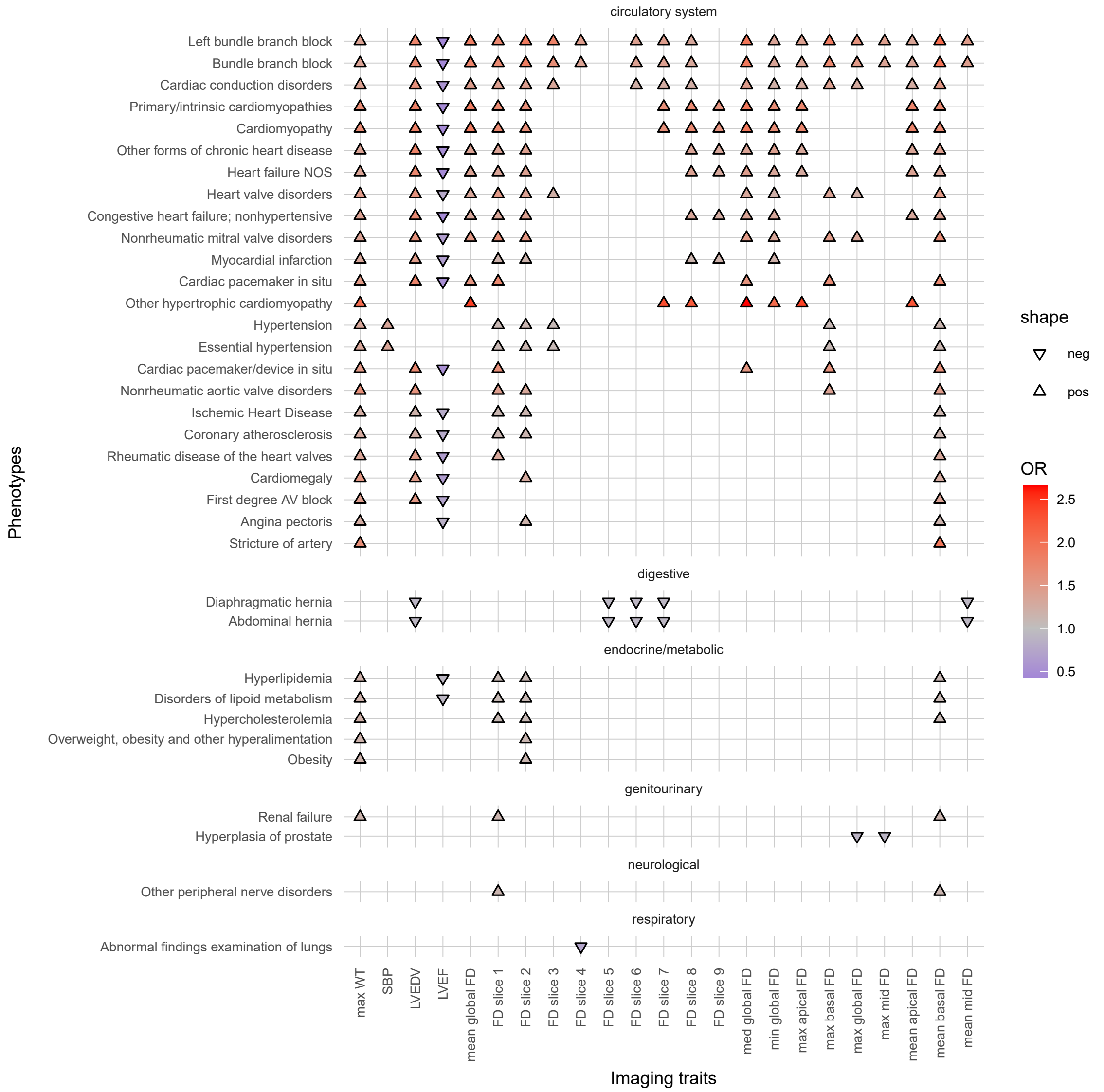

**Figure S 32.** PheWAS for summary trabeculation without aggregation.

#### Supplementary tables

Residuals:

| Min | 1Q | Median | 3Q | Max |
|---|---|---|---|---|
| -0.085824 | -0.019476 | -0.002443 | 0.017250 | 0.136712 |

| Coefficients: | Estimate | Std. Error | t value | Pr(>\|t\|) |
|---|---|---|---|---|
| (Intercept) | 1.490e+01 | 1.910e+00 | 7.798 | 6.44e-15 *** |
| Age at scan | 9.063e-04 | 2.953e-04 | 3.069 | 0.002150 ** |
| $Age^2$ | -5.730e-06 | 2.343e-06 | -2.446 | 0.014448 * |
| Imaging centre | -1.252e-03 | 1.732e-04 | -7.228 | 4.98e-13 *** |
| Sex | 4.002e-03 | 2.447e-03 | 1.635 | 0.101990 |
| Vigorous activity | 7.753e-04 | 7.661e-05 | 10.120 | <2e-16 *** |
| PC1 | 1.191e-05 | 4.222e-06 | 2.821 | 0.004792 ** |
| PC2 | 3.885e-05 | 6.654e-06 | 5.839 | 5.30e-09 *** |
| PC3 | -3.408e-05 | 1.201e-05 | -2.836 | 0.004567 ** |
| PC4 | 1.668e-05 | 1.570e-05 | 1.063 | 0.287978 |
| PC5 | 2.447e-05 | 1.951e-05 | 1.254 | 0.209840 |
| PC6 | -3.126e-05 | 3.604e-05 | -0.867 | 0.385739 |
| PC7 | -2.239e-05 | 3.367e-05 | -0.665 | 0.506029 |
| PC8 | -1.207e-04 | 3.384e-05 | -3.568 | 0.000361 *** |
| PC9 | -2.198e-05 | 3.775e-05 | -0.582 | 0.560404 |
| PC10 | 8.451e-05 | 4.391e-05 | 1.925 | 0.054288 . |
| BSA | 1.886e-02 | 9.122e-04 | 20.679 | <2e-16 *** |
| SBP | 7.318e-05 | 8.359e-06 | 8.755 | <2e-16 *** |
| Age:Sex | -3.029e-05 | 3.792e-05 | -0.799 | 0.424467 |

Signif. codes: 0 '***' 0.001 '**' 0.01 '*' 0.05 '.' 0.1 ' ' 1
Residual standard error: 0.02776 on 38226 degrees of freedom
Multiple R-squared: 0.04108, Adjusted R-squared: 0.04063
F-statistic: 90.99 on 18 and 38226 DF, p-value: <2.2e-16

**Table S 1.** Model information from the multiple linear regression of covariates when adjusting mean global trabeculation. PC, genetic principal component.

| Imaging trait | $h^2_{snp}$ | $cov_g$ | $r_g$ |
|---|---|---|---|
| LVEDV | 0.30 - 0.39 | 0.15 - 0.16 | 0.40 - 0.43 |
| SBP | 0.26 - 0.27 | -0.01 - 0.00 | -0.05 - 0.00 |
| MaxWT | 0.21 - 0.32 | -0.04 - 0.00 | -0.10 - -0.05 |
| LVEF | 0.26 - 0.27 | -0.01 - -0.04 | -0.11 - -0.14 |

**Table S 2.** Genetic covariance ($cov_g$) and correlation ($r_g$) with mean global trabeculation. Mean global trabeculation had a heritability estimate of 43%. The estimates are shown with and without adjustment for covariates. The analyses were completed on 38,245 participants of the UK Biobank population.

| | n=38245 | | n=9558 | | | |
|---|---|---|---|---|---|---|
| **Traits** | **Mean** | **SD** | **Mean** | **SD** | **$h^2$** | **SE** |
| Age (recruitment) | 54.90 | 7.47 | 55.87 | 7.93 | | |
| Age (imaging) | 63.63 | 7.57 | 67.73 | 7.86 | | |
| Female | 52% | | 51% | | | |
| White British | 87% | | 85% | | | |
| Slice 1 | 1.13 | 0.06 | 1.13 | 0.06 | 0.15 | 0.02 |
| Slice 2 | 1.14 | 0.05 | 1.14 | 0.05 | 0.20 | 0.02 |
| Slice 3 | 1.21 | 0.04 | 1.21 | 0.04 | 0.29 | 0.02 |
| Slice 4 | 1.25 | 0.03 | 1.26 | 0.03 | 0.28 | 0.02 |
| Slice 5 | 1.24 | 0.04 | 1.24 | 0.04 | 0.36 | 0.02 |
| Slice 6 | 1.20 | 0.05 | 1.20 | 0.05 | 0.39 | 0.02 |
| Slice 7 | 1.15 | 0.05 | 1.15 | 0.05 | 0.37 | 0.02 |
| Slice 8 | 1.11 | 0.05 | 1.11 | 0.05 | 0.28 | 0.02 |
| Slice 9 | 1.10 | 0.05 | 1.10 | 0.05 | 0.14 | 0.02 |
| Min Global FD | 1.08 | 0.03 | 1.08 | 0.03 | 0.16 | 0.02 |
| Mean Global FD | 1.17 | 0.03 | 1.17 | 0.03 | 0.43 | 0.02 |
| Med Global FD | 1.17 | 0.04 | 1.17 | 0.04 | 0.35 | 0.02 |
| Max Global FD | 1.26 | 0.03 | 1.26 | 0.03 | 0.34 | 0.02 |
| Min Basal FD | 1.12 | 0.04 | 1.10 | 0.02 | 0.02 | 0.02 |
| Mean Basal FD | 1.16 | 0.04 | 1.16 | 0.04 | 0.26 | 0.02 |
| Med Basal FD | 1.15 | 0.05 | 1.10 | 0.02 | 0.02 | 0.02 |
| Max Basal FD | 1.22 | 0.04 | 1.22 | 0.04 | 0.25 | 0.02 |
| Min Mid FD | 1.20 | 0.04 | 1.16 | 0.03 | 0.02 | 0.02 |
| Mean Mid FD | 1.23 | 0.03 | 1.23 | 0.03 | 0.42 | 0.02 |
| Med Mid FD | 1.24 | 0.04 | 1.16 | 0.03 | 0.02 | 0.02 |
| Max Mid FD | 1.26 | 0.03 | 1.26 | 0.03 | 0.34 | 0.02 |
| Min Apical FD | 1.09 | 0.04 | 1.13 | 0.05 | 0.03 | 0.02 |
| Mean Apical FD | 1.12 | 0.04 | 1.12 | 0.04 | 0.32 | 0.02 |
| Med Apical FD | 1.11 | 0.05 | 1.13 | 0.05 | 0.03 | 0.02 |
| Max Apical FD | 1.16 | 0.05 | 1.16 | 0.05 | 0.36 | 0.02 |

**Table S 3.** Summary information on the two groups of imaged UK Biobank participants and heritability estimates for trabeculation measures. Slice, imaging slice; Min, minimum; Med, median; FD, fractal dimension measure of trabeculation; SD, standard deviation; SE, standard error.

| Trait | Inflation factor (lambda) |
|---|---|
| Slice 1 | 1.02 |
| Slice 2 | 1.02 |
| Slice 3 | 1.03 |
| Slice 4 | 1.03 |
| Slice 5 | 1.04 |
| Slice 6 | 1.04 |
| Slice 7 | 1.04 |
| Slice 8 | 1.03 |
| Slice 9 | 1.02 |
| Max Apical | 1.04 |
| Max Basal | 1.03 |
| Max Global | 1.04 |
| Max Mid | 1.04 |
| Mean Apical | 1.03 |
| Mean Basal | 1.03 |
| Mean Global | 1.05 |
| Mean Mid | 1.05 |
| Med Apical | 1.01 |
| Med Basal | 1.01 |
| Med Global | 1.04 |
| Med Mid | 1.01 |
| Min Apical | 1.01 |
| Min Basal | 1.01 |
| Min Global | 1.02 |
| Min Mid | 1.01 |

**Table S 4.** Inflation factor from genome-wide association studies. Slice, image slices.

**Table S 5. See excel file for extended Table S5.** The table depicts the independent significant GWAS loci for trabecular measures as observed from LocusZoom. name, random ID of independent locus; nocmhf?, whether the locus was also observed when participants in the UK Biobank with cardiomyopathy or heart failure were removed; nocm?, whether the locus was also observed when participants in the UK Biobank with cardiomyopathy were removed; Region, position of locus in GrCh38; Gene1-3; nearest genes from LocusZoom.

**Table S 6. See excel file for extended Table S6.** Summary of Mendelian randomisation results. The table depicts the results for trabeculation as outcome and exposure with dilated cardiomyopathy (DCM), hypertrophic cardiomyopathy (HCM), and heart failure (HF). Five methods were used to assess for causality, with the number of SNPs included in the analysis (nsnp), beta effect size (b), standard error (se), and p-value (pval) shown.

**Table S 7. See excel file for extended Table S7.** The table depicts the validated rare variant association studies results for trabecular measures. CHROM, chromosome; GENPOS, gene position; ALLELE1, masks; A1FREQ, allele frequency; N, sample size of discovery cohort; TEST, RVAS method analysed by Regenie software; SE, standard error; LOG10P, p-value of association.

**Table S 8. See excel file for extended Table S8.** The table depicts the phenome-wide association studies results for imaging and trabecular measures. SE, standard error; OR, odds ratio; p, p-value of association; n_total phenotype, number of participants with imaging measure.

**Table S 9. See excel file for extended Table S9.** The table depicts the novel, independent, significant GWAS loci for trabecular measures for both validation and discovery datasets as observed from LocusZoom. name, random ID of independent locus; Region, position of locus in GrCh38; Gene1-3; nearest genes from LocusZoom.

**Table S 10. See excel file for extended Table S10.** The table depicts the novel, rare variant association studies results for trabecular measures using both discovery and validation cohorts. CHROM, chromosome; GENPOS, gene position; ALLELE1, masks; A1FREQ, allele frequency; N, sample size of discovery cohort; TEST, RVAS method analysed by Regenie software; SE, standard error; LOG10P, p-value of association.

**Table S 11. See excel file for extended Table S11.** Significant transcriptome-wide association study results for trabecular measures using both discovery and validation cohorts for cardiac tissues only (left ventricle and atrial appendage).

**Table S 12. See excel file for extended Table S12.** Significant transcriptome-wide association study results for trabecular measures using both discovery and validation cohorts.

**Table S 13. See excel file for extended Table S13.** Significant Gene Ontology (GO) Resource Enrichment analysis results using Panther on all genes in Tables S5 and S9. A PANTHER overrepresentation test (Released 20231017) was performed on the GO Ontology human database (doi:10.5281/zenodo.10536401 Released 2024-01-17). Significance was assessed via fisher exact test with an FDR correction. The GO gene lists assessed are shown in bold.
